# Water, Sanitation, and Women’s Empowerment: A systematic review and qualitative metasynthesis

**DOI:** 10.1101/2021.10.26.21265535

**Authors:** Bethany A. Caruso, Amelia Conrad, Madeleine Patrick, Ajilé Owens, Kari Kviten, Olivia Zarella, Hannah Rogers, Sheela S. Sinharoy

## Abstract

**Background:** Water and sanitation programs historically have focused on women’s instrumental value in improving effectiveness and impact of programs, though focus is shifting to consider how programming and conditions may contribute to women’s empowerment an gender equality. To date no systematic review has comprehensively assessed and synthesized evidence on water and sanitation and women and girls’ empowerment. The primary aims of this review were to: a) identify empirical water and sanitation research that engaged empowerment and/or empowerment-related domains from a pre-specified conceptual model; b) tabulate and report how empowerment-related terminology was used, where and when research was conducted, what methods were leveraged, and if water and/or sanitation was the primary focus; c) synthesize findings by empowerment domain and water and/or sanitation focus.

**Methods and Findings:** The conceptual model of women’s and girls’ empowerment developed by van Eerdewijk et.al (2017) informed our search strategy and analysis. The model presents three interrelated domains (agency, resources, institutional structures) and 13 sub-domains of empowerment. We searched MEDLINE, EMBASE, CABI Global Health, PsycINFO, CINAHL and AGRICOLA for any peer-reviewed sources presenting research related to water and/or sanitation and either empowerment and/or related terms from the conceptual model (4 May 2020). Systematic and ancestry and decendency searching identified 12,616 publications, of which 257 were included following screening, representing 1,600,348 participants. We assessed all studies using the Mixed-Method Appraisal Tool (MMAT). We followed the ‘best-fit framework synthesis’ approach for analysis, using the domains and sub-domains of the conceptual model as codes to assess all included sources. During coding, we inductively identified two additional sub-domains relevant to water and sanitation: privacy and freedom of movement. Thematic analysis guided synthesis of coded text by domain and sub-domain. The majority of research took place in Asia (46%; 117) or Africa (40%; 102), engaged adults (69%; 177), and were published since 2010; (82%; 211). A greater proportion of studies focused on water (45%; 115) than sanitation (22%; 57) or both (33%; 85). Over half of articles use the term empowerment yet only 7% (17) provided a clear definition or conceptualization. Agency was the least commonly engaged domain (47%; 122) while the Resources domain was dominant (94%; 241). Measures for assessing empowerment and related domains is limited. This review was limited by only including sources in English and only includes menstruation-focused research in the context of water and sanitation.

**Conclusions:** Water and sanitation research specifically engaging women’s and girls’ empowerment in a well-defined or conceptualized manner is limited. A substantial body of research examining domains and sub-domains of empowerment exists, as does research that illuminates myriad negative impacts of water and sanitation conditions and circumstances women’s and girl’s well-being. Available research should be used to develop and evaluate programs focused on improving the life outcomes of women and girls, which has only been minimally conducted to date. A more comprehensive ‘transformative WASH’ that includes gender-transformative approaches to challenge and reduce systemic constraints on women’s and girls’ resources and agency is not only warranted but long overdue.

## Introduction

Water, sanitation, and hygiene (WASH) access, behaviors, experiences, and physical and social environments have been shown to influence multiple outcomes, including diarrheal disease, soil transmitted helminth and protozoa infection, active trachoma and schistosomiasis, pneumonia, anaemia, mental health and general well-being, economic productivity, school absence, and child growth and cognitive development.^1–14^ This demonstrated importance of WASH underlies Sustainable Development Goal (SDG) 6, which aims to “Ensure availability and sustainable management of water and sanitation for all.”^15^ Still, water and sanitation access remain out of reach for large proportions of the global population: 29% of the global population lacks access to water that is available when needed and free from chemical and fecal contamination, and 55% lacks access to household sanitation facilities that safely manage excreta.^16^ Furthermore, while SDG Target 6.2 emphasizes “paying special attention to the needs of women and girls,” who are recognized as WASH duty-bearers globally,^17–20^ data often fail to reflect the gender-specific benefits and harms of WASH conditions, behaviors, and interventions. Despite recognition of WASH as on the pathway to gender equality,^21^ a full understanding of the gendered effects of WASH remains limited, prompting calls for improved gender measurement, data, and learning.^19, 22–25^

Critical discourse on gender and WASH is evolving. While historically WASH programs focused on women’s instrumental value in improving effectiveness and impact of programs, focus has been shifting to consider how WASH programming may contribute to women’s empowerment^26–29^. Recent reviews examining WASH and gender further demonstrate this shift in focus.^30–32^ In their scoping review, Dery et al. (2020) explored how empowerment was used in WASH; five interrelated dimensions of empowerment were identified among the 13 included articles: access to information, participation, capacity building, leadership and accountability, and decision-making.^30^ MacArthur and colleagues (2020) conducted a critical review of WASH-gender literature from 2008-2018 to understand how WASH studies engaged gender equality. Their distant-reading analysis of the 155 included articles, which focused only on assessment of titles and abstracts, revealed that few engaged with gender transformational-aspects of gender equality.^31^

To date there have been no rigorous systematic reviews to assess and synthesize evidence on WASH and women and girls’ empowerment. The primary aims of this literature review were to: a) identify empirical water and sanitation research that engaged empowerment and/or empowerment-related domains; b) tabulate and report how empowerment-related terms were used, where and when research was conducted, what methods were leveraged, and if water and/or sanitation was the primary focus; c) synthesize findings by empowerment domain and water and/or sanitation focus.

## Methods

We report our review using Preferred Reporting Items for Systematic Reviews and Meta-Analyses (PRISMA) criteria (see Supplemental Table 1).

### Search Strategy

Our search strategy aimed to identify studies that engaged with and reported on water, sanitation, and empowerment, including associated domains and sub-domains of empowerment. The conceptual model of empowerment outlined by Van Eerdewijk et al. (2017),^33^ which extended work by Naila Kabeer,^34^ the World Bank,^35^ and CARE,^36^ guided our search and subsequent analyses. Table 1 provides definitions of empowerment, and the three domains (agency, resources, institutional structures) and 13 subdomains included in the model.

**Table 1.** Definitions of empowerment and related domains and sub-domains from Van Eerdewijk et al. (2017)

| Term | Definition |
| --- | --- |
| **Empowerment** | The expansion of choice and strengthening of voice through the transformation of power relations so women and girls have more control over their lives and futures. |
| **Disempowerment** | Unequal distribution of resources and women and girls' lack of control over their bodies and low self-esteem, combined with biased laws and policies and discriminatory gender norms and practices. |
| **1. Agency** | Women and girls pursuing goals, expressing voice and influencing and making decisions free from violence and retribution |
| **a. Decision-Making** | Women and girls influencing and making decisions, and establishing and acting on goals |
| **b. Leadership** | Women and girls leading, inspiring social change and effectively participating in governance |
| **c. Collective Action** | Women and girls gaining solidarity and taking action collectively on their interests to enhance their position and expand the realm of what is possible |
| **2. Resources** | The tangible and intangible capital and sources of power that women and girls have, own or use, individually or collectively, in the exercise of agency |
| **a. Bodily Integrity** | Women and girls' security and control over their bodies, and physical and mental well-being |
| **i. Safety & Security<sup>1</sup>** | Women and girls' freedom from acts or threats of violence, coercion or force |
| **ii. Health** | Women and girls' complete physical, mental and social well-being and not merely the absence of disease or infirmity |
| **b. Critical Consciousness** | Women and girls identifying and questioning how inequalities in power operate in their lives, and asserting and affirming their sense of self and their entitlement ('power-within') |
| **c. Assets** | Women and girls' control over tangible or intangible economic, social or productive resources that include (1) financial and productive assets, (2) knowledge and skills, (3) time and (4) social capital |
| **i. Financial & Productive Assets** | Women and girls' control over economic resources such as income, credit or savings, as well as long-term stocks of value like land, equipment, housing or livestock that can be owned, controlled or used by a person |
| **ii. Knowledge & Skills** | Women and girls' knowledge and skills (including life skills), and their abilities to apply knowledge to situations, obtained through high-quality formal or informal education, training or information |
| **iii. Social Capital** | Women and girls' relations and social networks that provide tangible and intangible value and support |
| **iv. Time** | Women and girls' control over their time and labor, which is key to time poverty and work burden |
| **3. Institutional Structures** | The social arrangements of formal and informal rules and practices that enable and constrain the agency of women and girls, and govern the distribution of resources |
| **a. Formal Laws & Policies** | Formally recognized rules of conduct or procedures established by nation states, international treaties and conventions, or local governance authorities that govern the rights and entitlements of women and girls |
| **b. Norms** | Collectively held expectations and beliefs of how women, men, girls, and boys should behave and interact in specific social settings and during different stages of their lives |
| **c. Relations** | The interactions and relations with key actors that women and girls experience in their daily lives |
| 1. In our analysis, we use the definition of sexual violence used by Van Eerdewijk et al. (2017) which comes from Krug et al. (2002): "any sexual act, attempt to obtain a sexual act, unwanted sexual comments or advances, or acts to traffic, or otherwise directed, against a person's sexuality using coercion, by any person regardless of their relationship to the victim, in any setting, including but not limited to home and work" (p. 149).<sup>37</sup> Forms of sexual violence included in this definition include: rape within marriage or relationships; rape by strangers; systematic rape during armed conflict; unwanted sexual advances or sexual harassment; sexual abuse of mentally or physically disabled people; sexual abuse of children; forced marriage or cohabitation including child marriage; denial of the right to use contraception or to adopt other measures to protect against sexually transmitted diseases; forced abortion; female genital mutilation and obligatory inspections for virginity; forced prostitution and trafficking of people for the purpose of sexual exploitation. |  |

We completed our search on 4 May 2020 for articles published in English in peer-reviewed sources on any date in the following databases: MEDLINE (PubMed), EMBASE, CABI Global Health, PsycINFO (EBSCOhost), CINAHL (EBSCOhost) and AGRICOLA (EBSCOhost) (Supplemental Table 2 for terms). One co-author (AC) identified additional articles by reviewing the reference lists of each included article (ancestry search) and by using Google Scholar to identify articles that cited each included article (descendancy search). Finally, additional articles identified by BC and SS not captured in the search were included.

### Study Eligibility

Any peer-reviewed article presenting primary or secondary research related to water and/or sanitation and either empowerment and/or one of the domains of empowerment from the conceptual model was eligible for inclusion. We included all countries, settings, human populations, and study designs; we excluded articles not in English.

To determine inclusion, one team member independently reviewed all titles and abstracts from the database search. Three other team members then split all titles and abstracts to complete a second review. When a consensus decision could not be made from the title and abstract, the full article was reviewed by two reviewers. When reviewers disagreed on eligibility, six members of the broader study team met to reach consensus on inclusion or exclusion.

### Analysis

One team member (MP) extracted and collated study design, setting, population, and relevant water and sanitation information from each study.

To assess if papers engaged ‘empowerment’ and like terms, we conducted a word search on all papers for ‘empow,’ which enabled identification of ‘empowerment’ and similar terms (e.g. empower, disempowered). We then classified how papers engaged empowerment-related terms using a four-tier classification tool we created; see Figure 1 for tier classifications.

**Figure 1:**
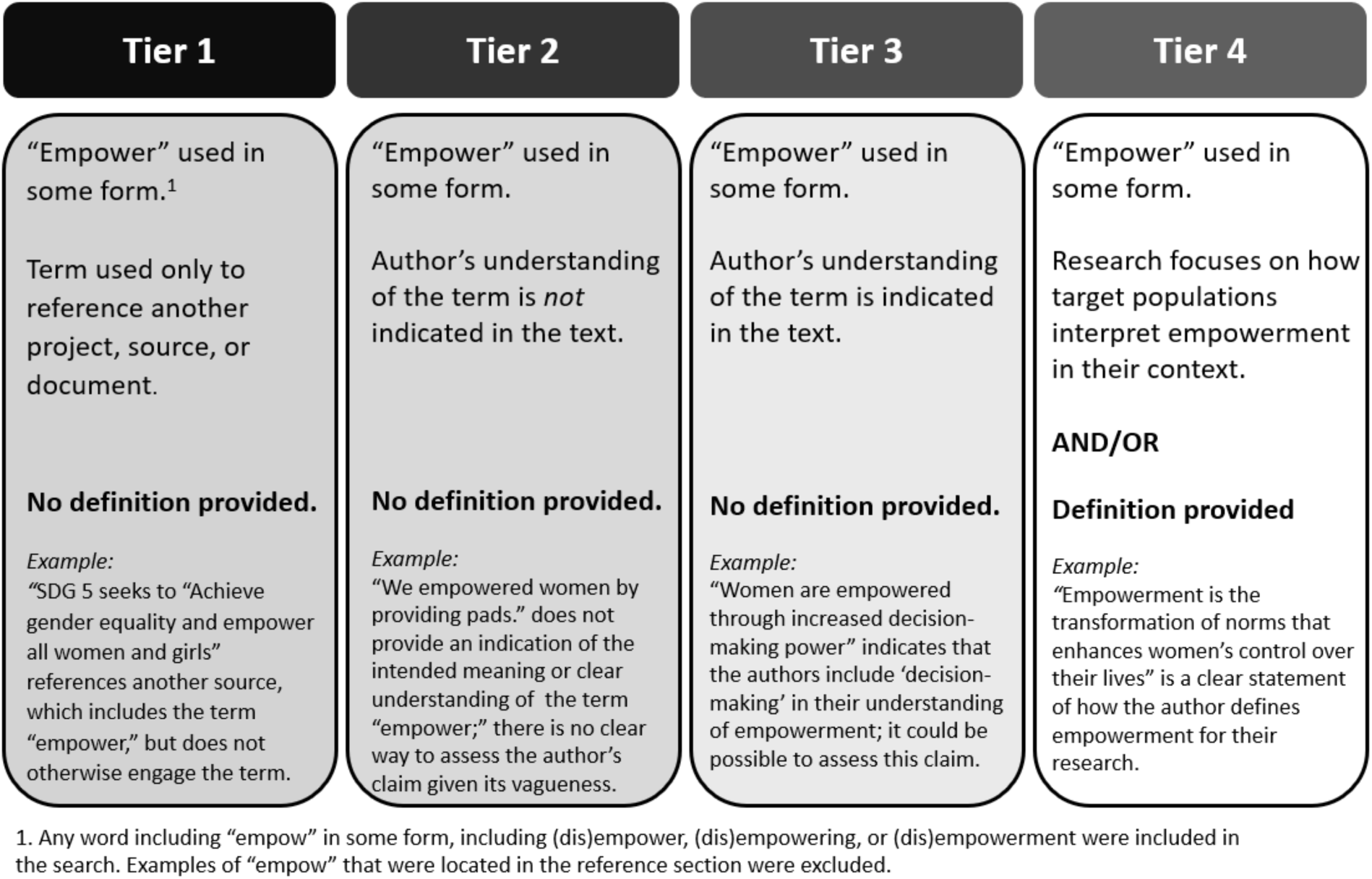
Four Tier Classification Schema for Article Engagement of ‘Empowerment’.

To classify and synthesize evidence on water, sanitation, and empowerment, we used the ‘best-fit framework synthesis’ approach.^38^ Using this method, themes are identified to use as codes *a priori* from pre-existing, guiding frameworks or models. This synthesis approach allows for the guiding framework or conceptual model to be modified as themes emerge inductively from the data.

We created and defined codes based on the empowerment subdomains in the conceptual model by Van Eerdewijk et al.^33^ (Table 1), including two additional empowerment-related subdomains identified iteratively through analysis, thus expanding the existing model: *Privacy* and *Freedom of Movement* (Figure 2). Other codes included water, sanitation, and menstruation to identify the broad topics engaged.

**Figure 2:**
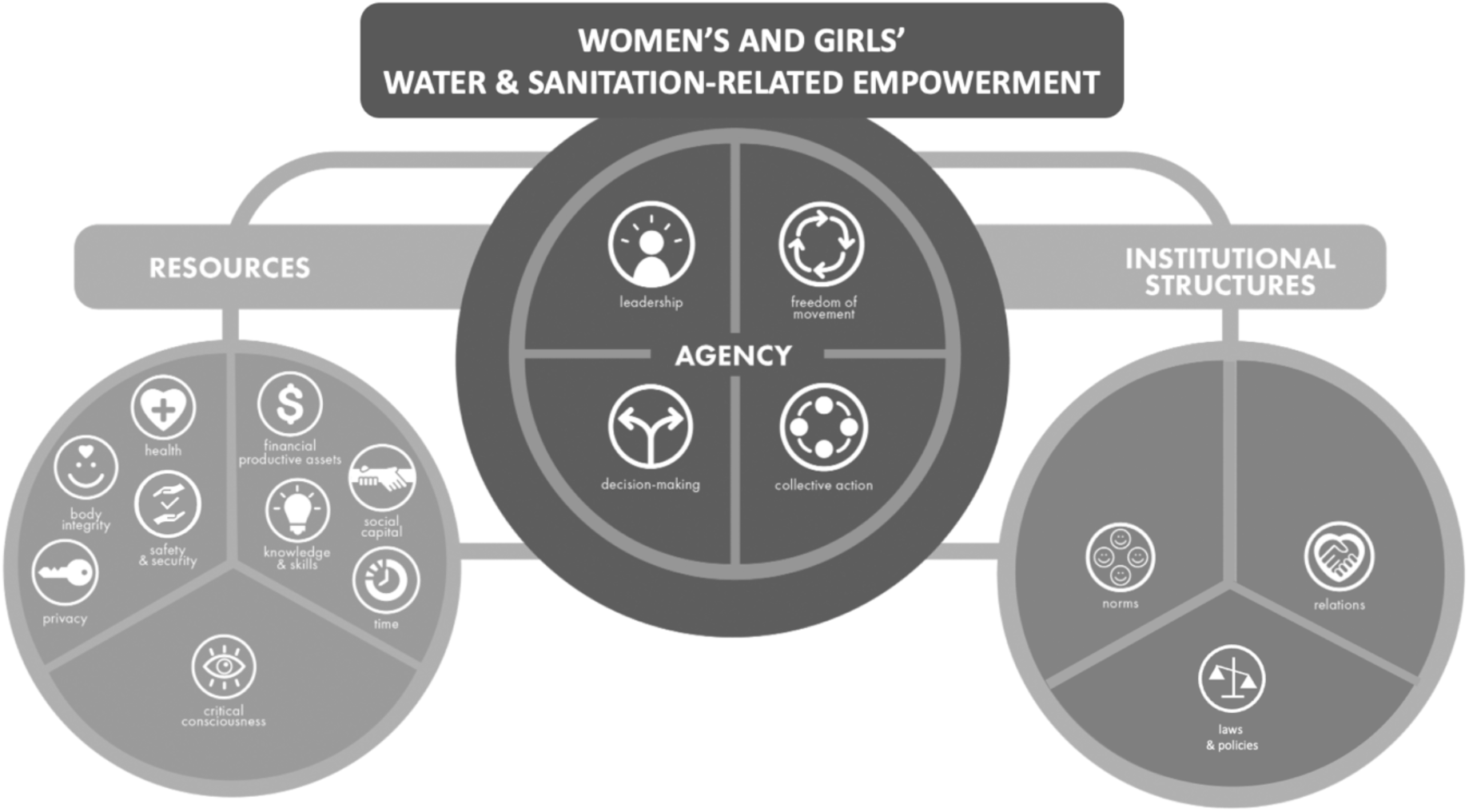
Domains and Sub-Domains of Women’s and Girls’ Water and Sanitation-Related Empowerment (Adapted from Van Eerdewijk et al. (2017)

All eligible papers were imported into MaxQDA (version 12)^39^ qualitative analysis software. Line-by-line coding of the results sections of eligible papers was carried out independently by two team members, who first coded two papers independently and compared to assess inter-coder agreement.

Using thematic analysis, two team members generated descriptive themes for each empowerment code based on the relevant coded texts. All coded segments containing both an empowerment-related code and a sanitation/water code (e.g. ‘safety’ and ‘sanitation’) were exported into an Excel file and analyzed further. Each subdomain of empowerment (e.g. ‘safety,’ ‘privacy’) was analyzed separately. Themes were refined into a smaller set of descriptive themes that outlined how experiences with water and sanitation related to the various subdomains of empowerment.

Descriptive themes were then used to describe how each domain/subdomain of empowerment has been researched in the literature.

### Study Quality Appraisal

We assessed all studies using the Mixed-Method Appraisal Tool (MMAT) 2018 developed by Pluye et al.^40^ and updated in 2018^41^ for the appraisal of qualitative, quantitative, and mixed-methods studies. One team member performed quality appraisals of all studies and a second team member performed a quality appraisal agreement check on 10% of studies. Qualitative and most quantitative studies were assessed using the five-criteria questionnaire; one criterion was dropped for randomized control trials because we did not consider it indicative of quality (‘Did participants adhere to the assigned intervention?’). Mixed-methods studies were assessed using the relevant independent questionnaires for qualitative and quantitative work and a five-criteria questionnaire for mixed-methods; the lowest of the three scores was used as the quality score. Possible scores were 0-5 across study types (5 is the best). Because the primary aim of this work is to understand if and how research engaged water and sanitation and empowerment themes, all studies were retained regardless of scores.

In their meta-synthesis of sanitation and well-being, Sclar et al. (2018)^14^ note that qualitative research is explicitly unblinded, subjective, and self-reported, and thus likely to produce poor scores from bias assessment tools. Because this review is exploratory, we excluded bias assessment, reasoning that qualitative studies would have low bias scores by default (despite rich insights), and therefore the activity would introduce bias.

## Results

Figure 3 shows the review strategy, including reasons for exclusion. We included 257 articles— 129 qualitative, 54 quantitative, and 74 mixed methods—representing an estimated 1,600,348 participants (Supplemental Table 3 describes all included studies). Articles largely featured research from Asia (48%) and Africa (42%), and focused on adult participants (69%) (Table 2). The earliest paper was published in 1989; 82% (211) of articles were published since 2010 and 60% (153) since 2015 (Figure 4).

**Figure 3:**
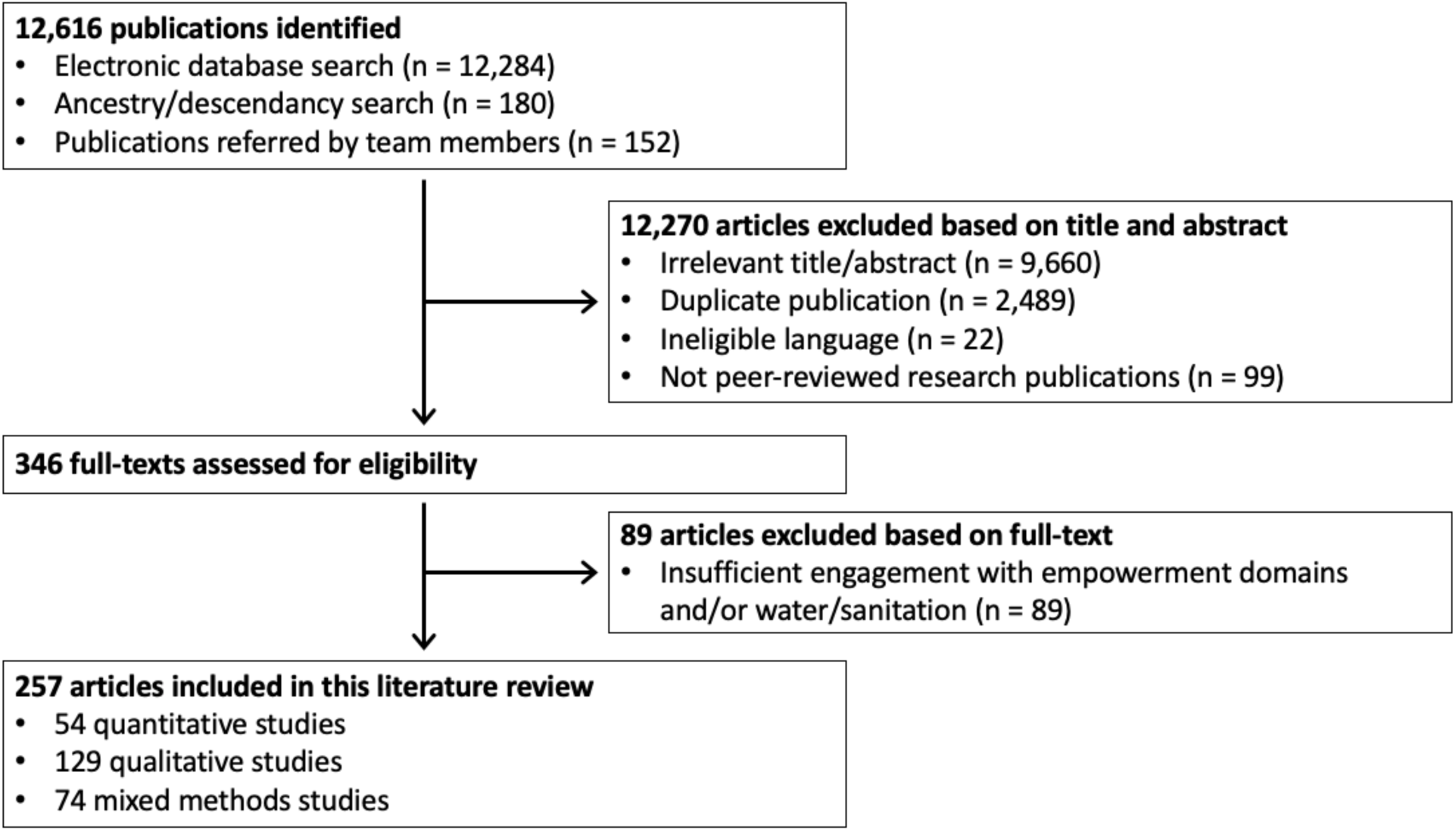
PRISMA flow diagram of publications considered for the review.

**Figure 4:**
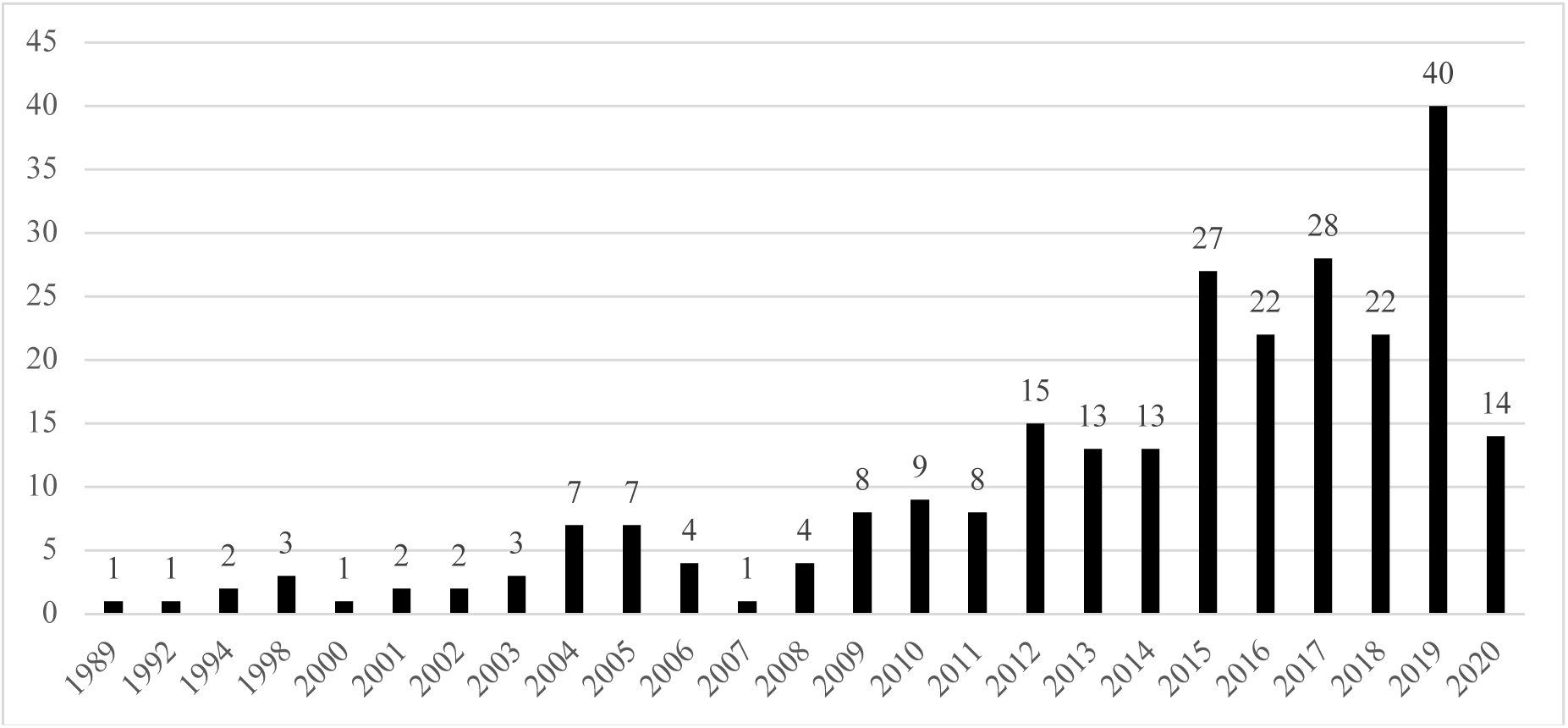
Number of Articles Included in Search by Date of Publication, through 4 May 2020 (N=257)

**Table 2.**
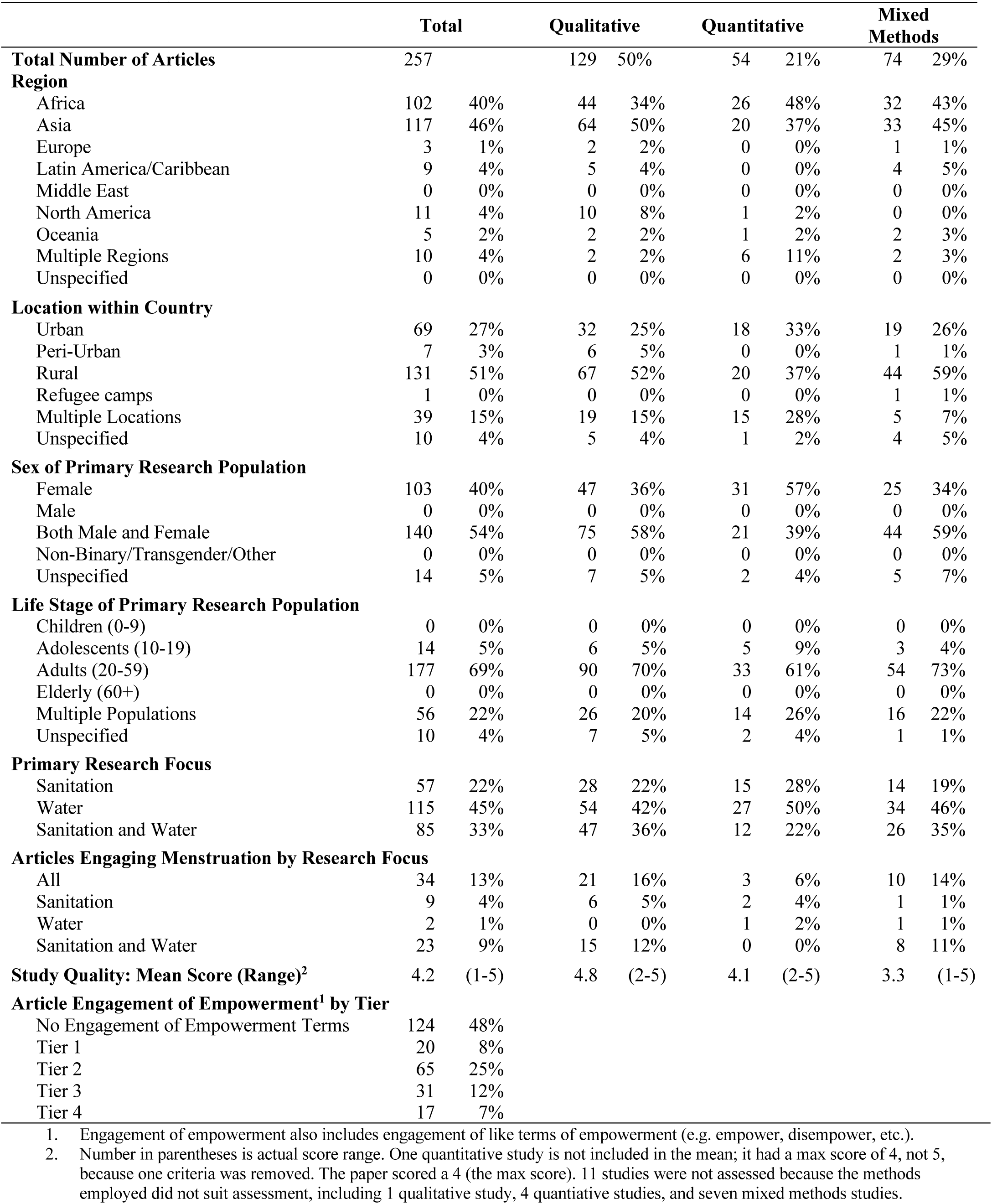
Summary Information about Included Sources (N= 257)

|  | **Total** |  | **Qualitative** |  | **Quantitative** |  | **Mixed Methods** |  |
| --- | --- | --- | --- | --- | --- | --- | --- | --- |
| **Total Number of Articles** | 257 |  | 129 | 50% | 54 | 21% | 74 | 29% |
| **Region** |  |  |  |  |  |  |  |  |
| Africa | 102 | 40% | 44 | 34% | 26 | 48% | 32 | 43% |
| Asia | 117 | 46% | 64 | 50% | 20 | 37% | 33 | 45% |
| Europe | 3 | 1% | 2 | 2% | 0 | 0% | 1 | 1% |
| Latin America/Caribbean | 9 | 4% | 5 | 4% | 0 | 0% | 4 | 5% |
| Middle East | 0 | 0% | 0 | 0% | 0 | 0% | 0 | 0% |
| North America | 11 | 4% | 10 | 8% | 1 | 2% | 0 | 0% |
| Oceania | 5 | 2% | 2 | 2% | 1 | 2% | 2 | 3% |
| Multiple Regions | 10 | 4% | 2 | 2% | 6 | 11% | 2 | 3% |
| Unspecified | 0 | 0% | 0 | 0% | 0 | 0% | 0 | 0% |
| **Location within Country** |  |  |  |  |  |  |  |  |
| Urban | 69 | 27% | 32 | 25% | 18 | 33% | 19 | 26% |
| Peri-Urban | 7 | 3% | 6 | 5% | 0 | 0% | 1 | 1% |
| Rural | 131 | 51% | 67 | 52% | 20 | 37% | 44 | 59% |
| Refugee camps | 1 | 0% | 0 | 0% | 0 | 0% | 1 | 1% |
| Multiple Locations | 39 | 15% | 19 | 15% | 15 | 28% | 5 | 7% |
| Unspecified | 10 | 4% | 5 | 4% | 1 | 2% | 4 | 5% |
| **Sex of Primary Research Population** |  |  |  |  |  |  |  |  |
| Female | 103 | 40% | 47 | 36% | 31 | 57% | 25 | 34% |
| Male | 0 | 0% | 0 | 0% | 0 | 0% | 0 | 0% |
| Both Male and Female | 140 | 54% | 75 | 58% | 21 | 39% | 44 | 59% |
| Non-Binary/Transgender/Other | 0 | 0% | 0 | 0% | 0 | 0% | 0 | 0% |
| Unspecified | 14 | 5% | 7 | 5% | 2 | 4% | 5 | 7% |
| **Life Stage of Primary Research Population** |  |  |  |  |  |  |  |  |
| Children (0-9) | 0 | 0% | 0 | 0% | 0 | 0% | 0 | 0% |
| Adolescents (10-19) | 14 | 5% | 6 | 5% | 5 | 9% | 3 | 4% |
| Adults (20-59) | 177 | 69% | 90 | 70% | 33 | 61% | 54 | 73% |
| Elderly (60+) | 0 | 0% | 0 | 0% | 0 | 0% | 0 | 0% |
| Multiple Populations | 56 | 22% | 26 | 20% | 14 | 26% | 16 | 22% |
| Unspecified | 10 | 4% | 7 | 5% | 2 | 4% | 1 | 1% |
| **Primary Research Focus** |  |  |  |  |  |  |  |  |
| Sanitation | 57 | 22% | 28 | 22% | 15 | 28% | 14 | 19% |
| Water | 115 | 45% | 54 | 42% | 27 | 50% | 34 | 46% |
| Sanitation and Water | 85 | 33% | 47 | 36% | 12 | 22% | 26 | 35% |
| **Articles Engaging Menstruation by Research Focus** |  |  |  |  |  |  |  |  |
| All | 34 | 13% | 21 | 16% | 3 | 6% | 10 | 14% |
| Sanitation | 9 | 4% | 6 | 5% | 2 | 4% | 1 | 1% |
| Water | 2 | 1% | 0 | 0% | 1 | 2% | 1 | 1% |
| Sanitation and Water | 23 | 9% | 15 | 12% | 0 | 0% | 8 | 11% |
| **Study Quality: Mean Score (Range)<sup>2</sup>** | 4.2 | (1-5) | 4.8 | (2-5) | 4.1 | (2-5) | 3.3 | (1-5) |
| **Article Engagement of Empowerment<sup>1</sup> by Tier** |  |  |  |  |  |  |  |  |
| No Engagement of Empowerment Terms | 124 | 48% |  |  |  |  |  |  |
| Tier 1 | 20 | 8% |  |  |  |  |  |  |
| Tier 2 | 65 | 25% |  |  |  |  |  |  |
| Tier 3 | 31 | 12% |  |  |  |  |  |  |
| Tier 4 | 17 | 7% |  |  |  |  |  |  |

1. Engagement of empowerment also includes engagement of like terms of empowerment (e.g. empower, disempower, etc.).
2. Number in parentheses is actual score range. One quantitative study is not included in the mean; it had a max score of 4, not 5, because one criteria was removed. The paper scored a 4 (the max score). 11 studies were not assessed because the methods employed did not suit assessment, including 1 qualitative study, 4 quantitative studies, and seven mixed methods studies.

### Study Quality

Supplemental Table 4 presents study quality scores for each included article. Mean study quality was 4.2 overall indicating good quality, 4.8 for qualitative studies, 4.1 for quantitative studies, and 3.3 for mixed methods studies (5 is maximum). (Table 2).

### Water and Sanitation Research Engaging Empowerment and Associated Domains

The terms “empower,” “empowerment,” “empowering,” “disempowerment,” or “disempowering” featured in the text of 133 (52%) articles. Only 17 (7%) articles included a specific definition or conceptualization of empowerment to inform their research (see Table 2; Supplemental Table 5 for specific definitions used);^29, 42–57^ 12 (5%) had a specific aim or research question focused broadly on empowerment.^29, 42, 43, 45, 50, 52, 54, 56, 58–61^

One hundred fifteen (45%) articles focused on water, 57 (22%) on sanitation, 85 (33%) on both; 34 (13%) engaged menstruation. Table 3 summarizes which studies contributed to each domain or subdomain; Supplemental Table 6 collates all menstruation-related articles by domain and subdomain. Of the domains of empowerment, the resources domain was the most represented (241 articles; 94%) (Supplemental Figure 1). 181 (70%) articles engaged at least two of the empowerment domains; 113 (44%) engaged all three (Supplemental Figure 1).

**Table 3.**
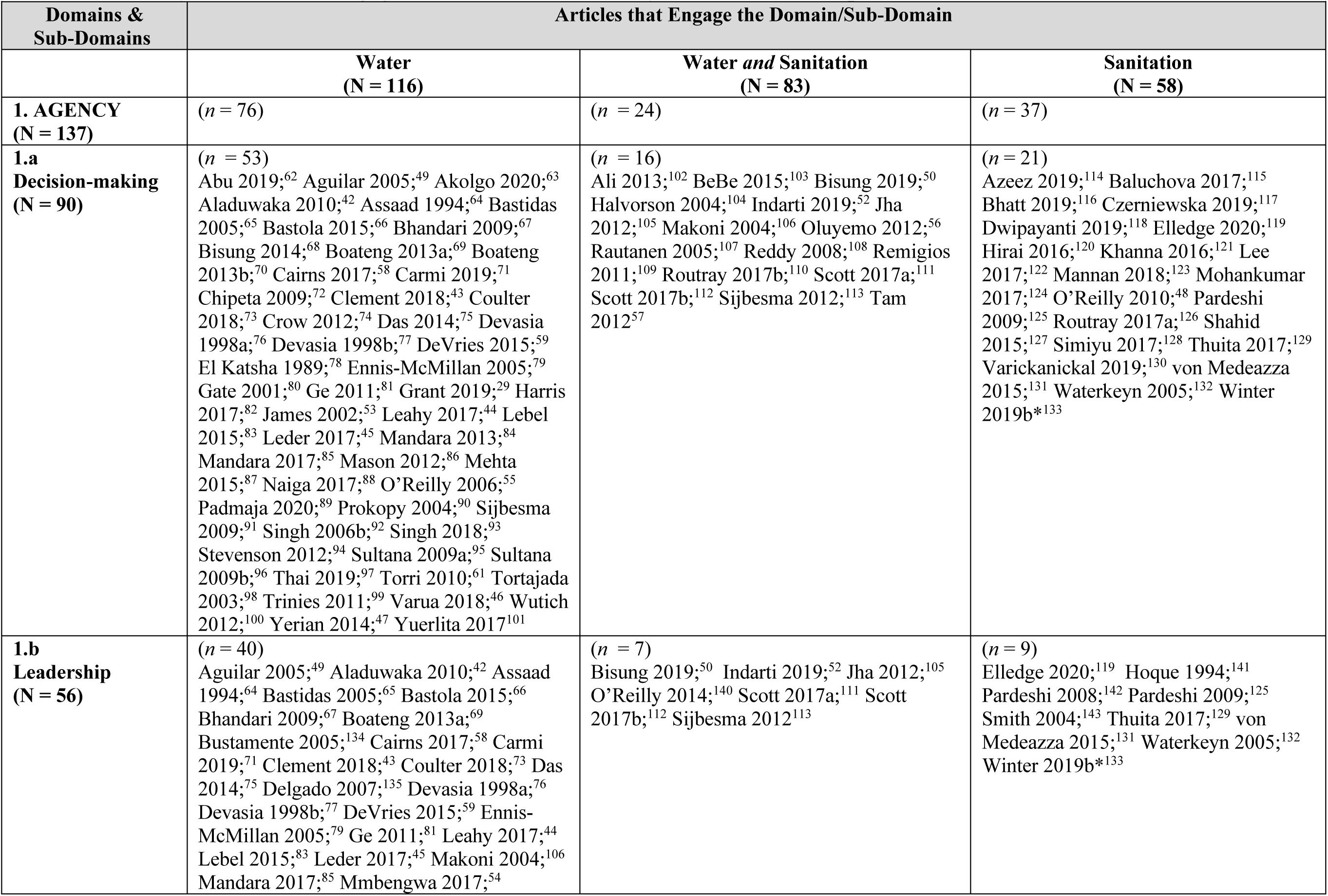
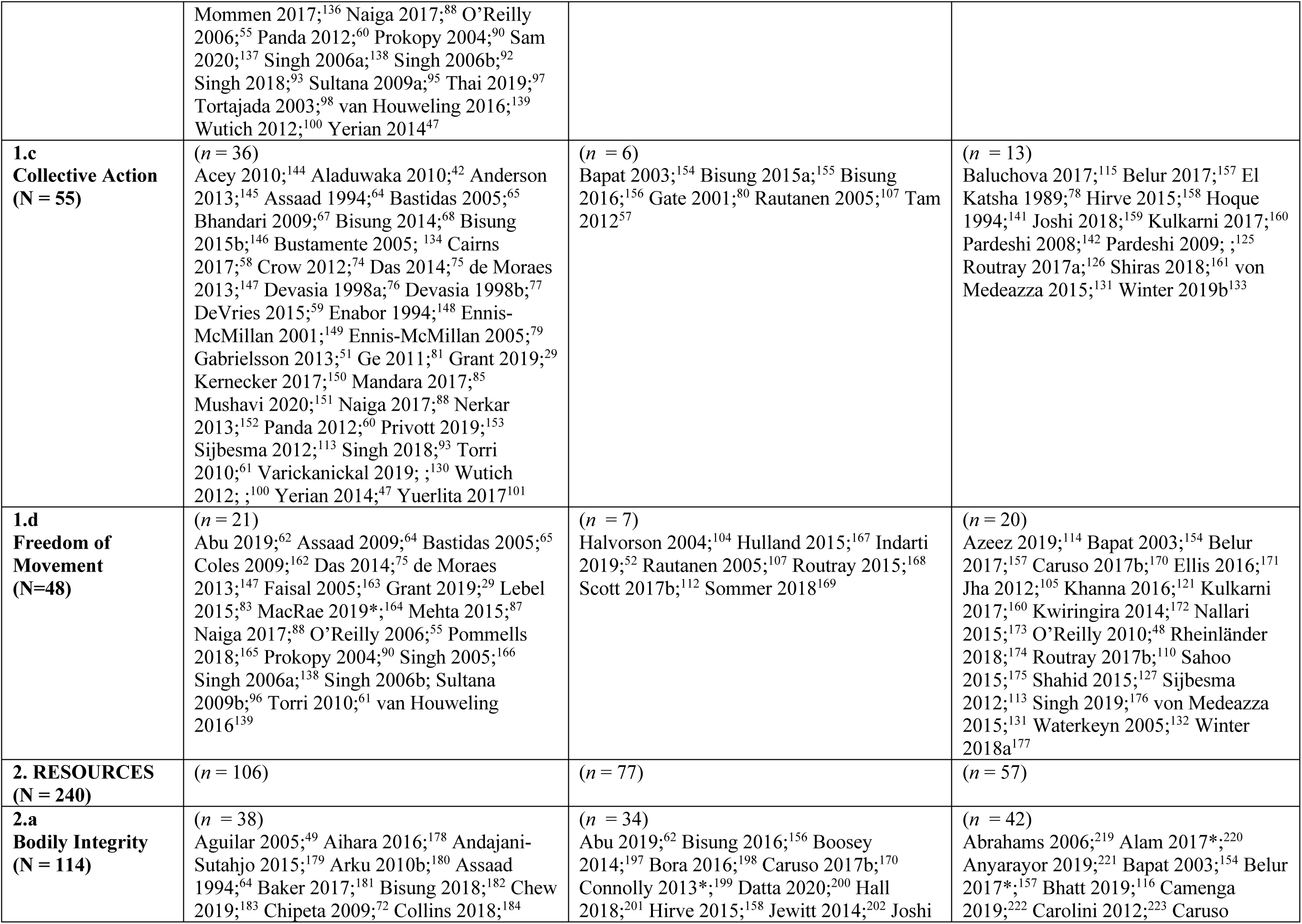
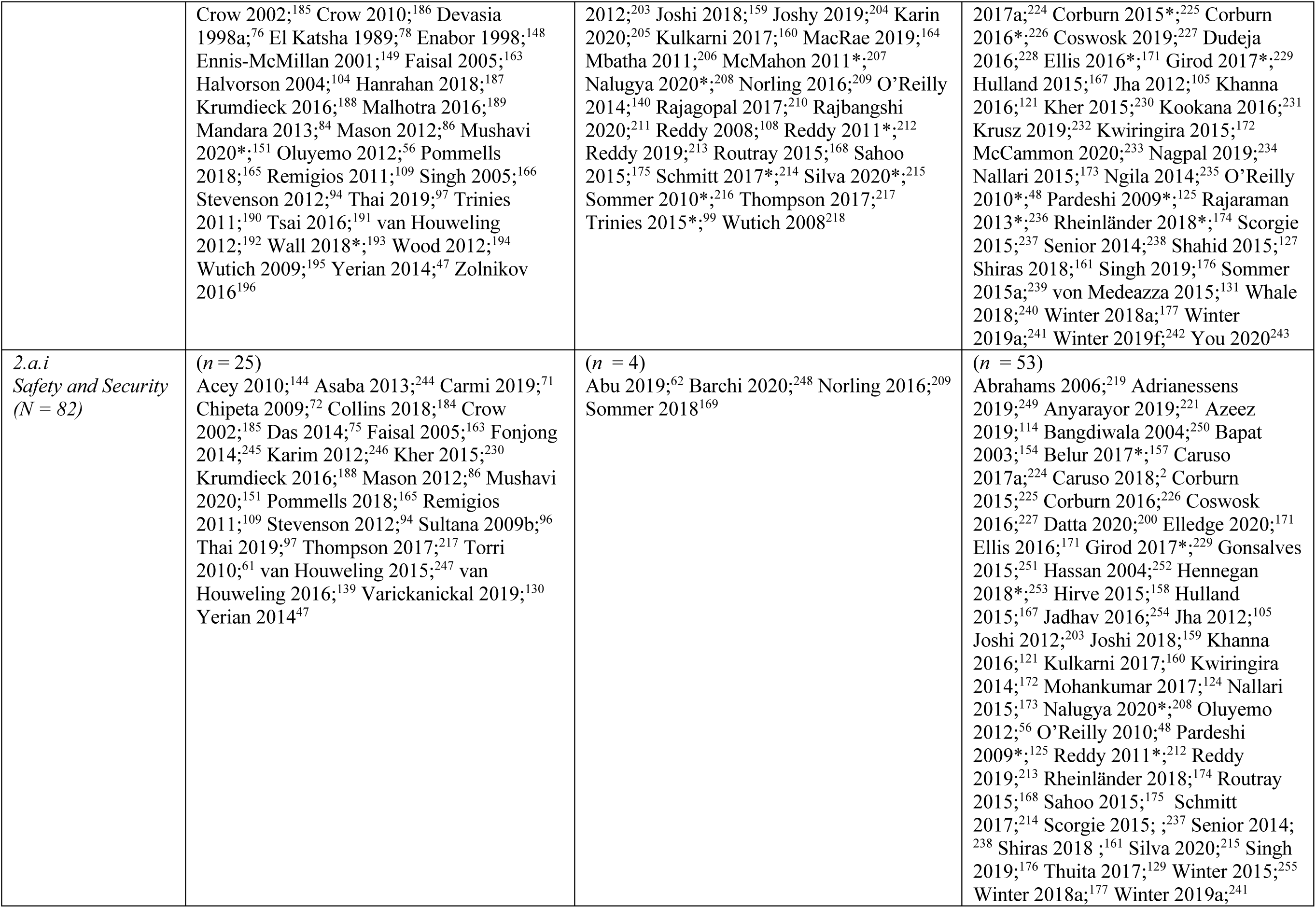
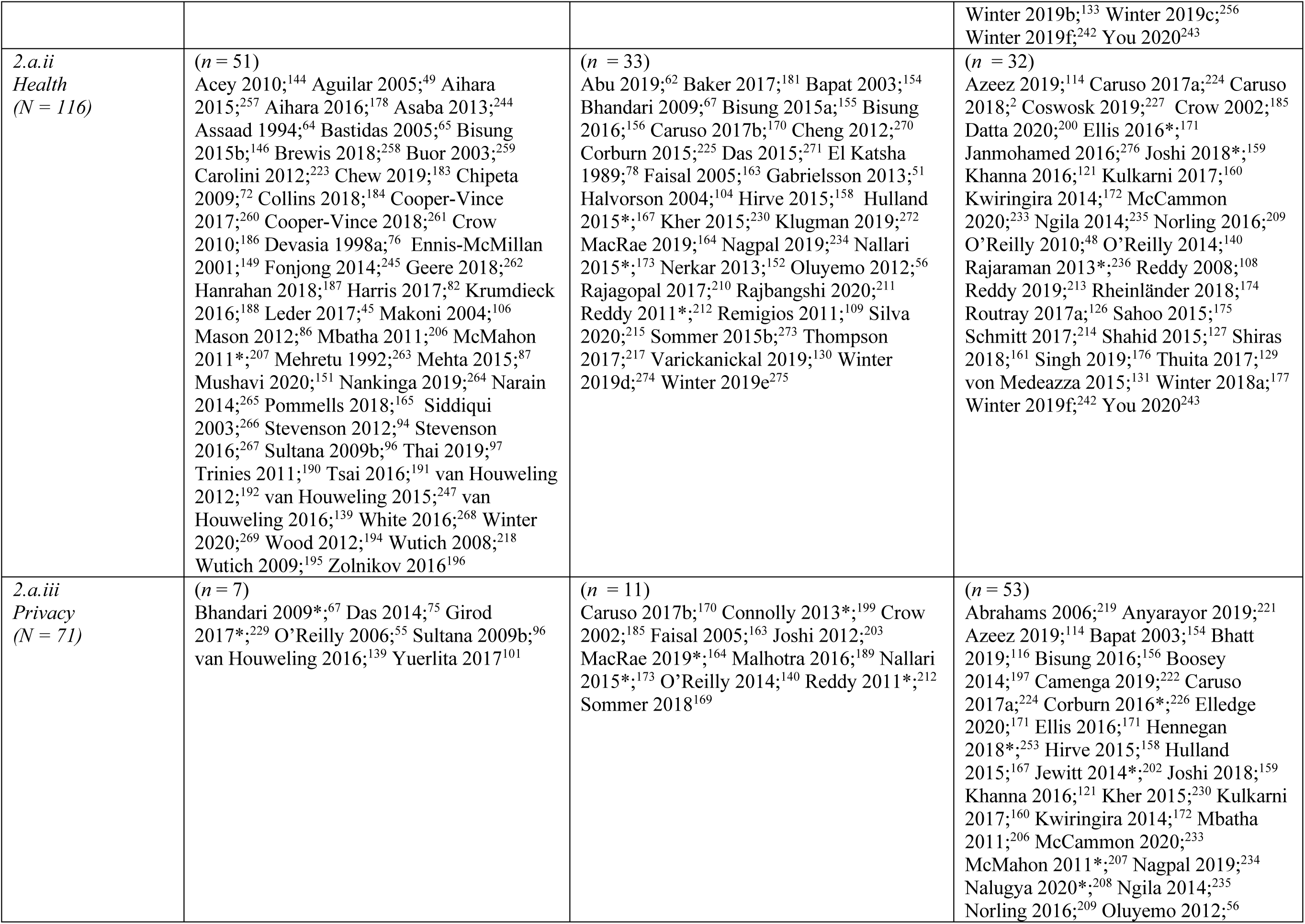
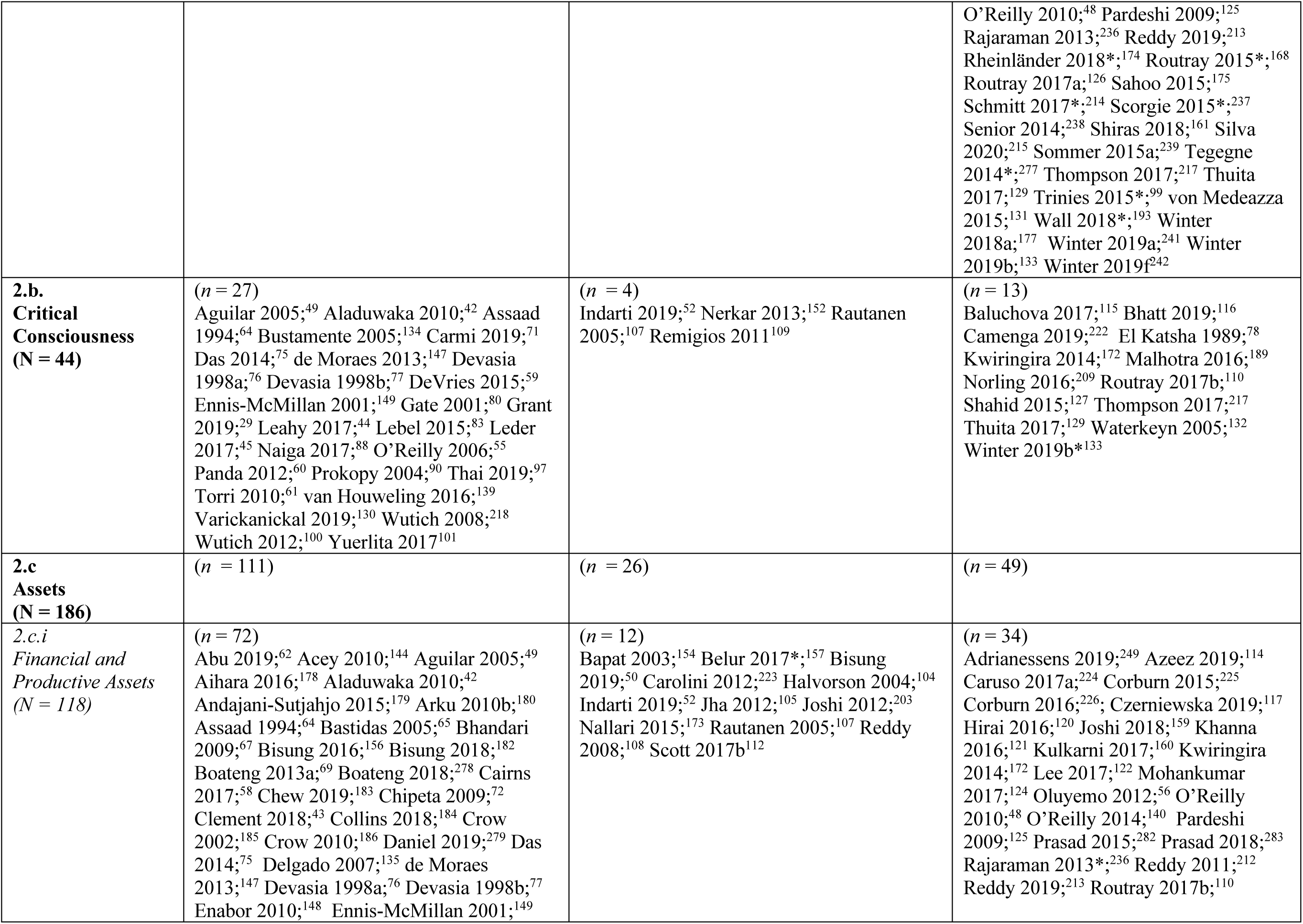
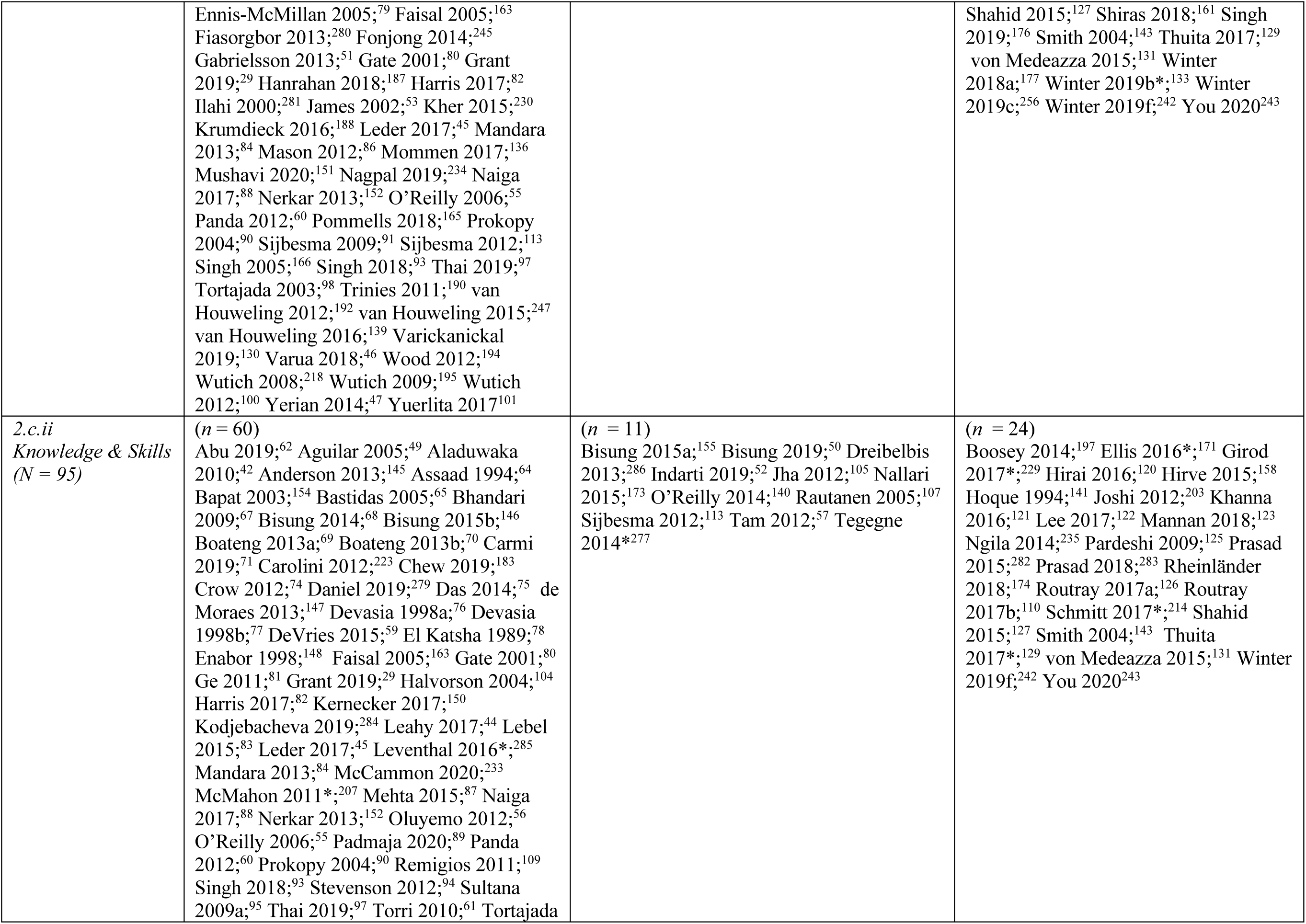
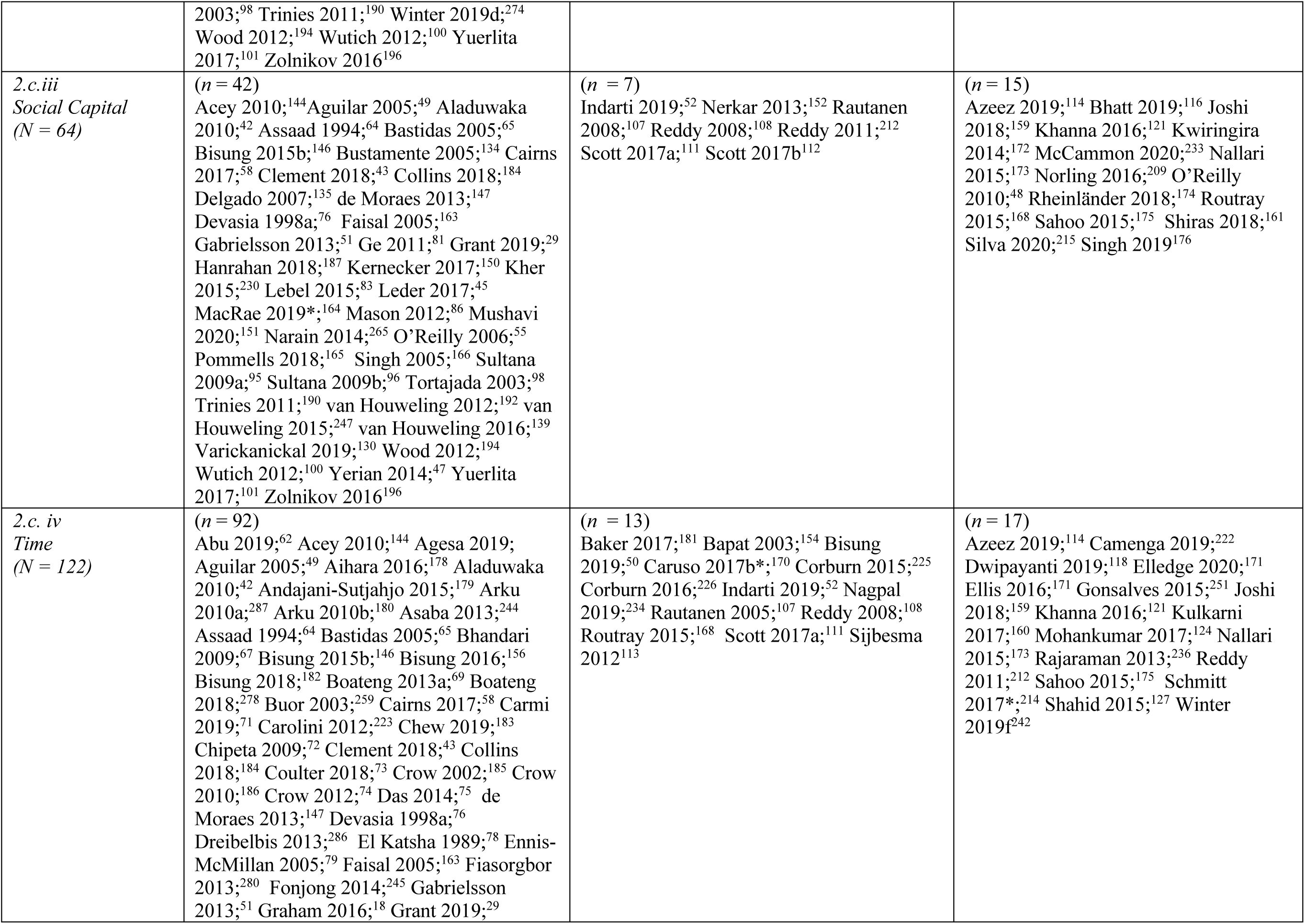
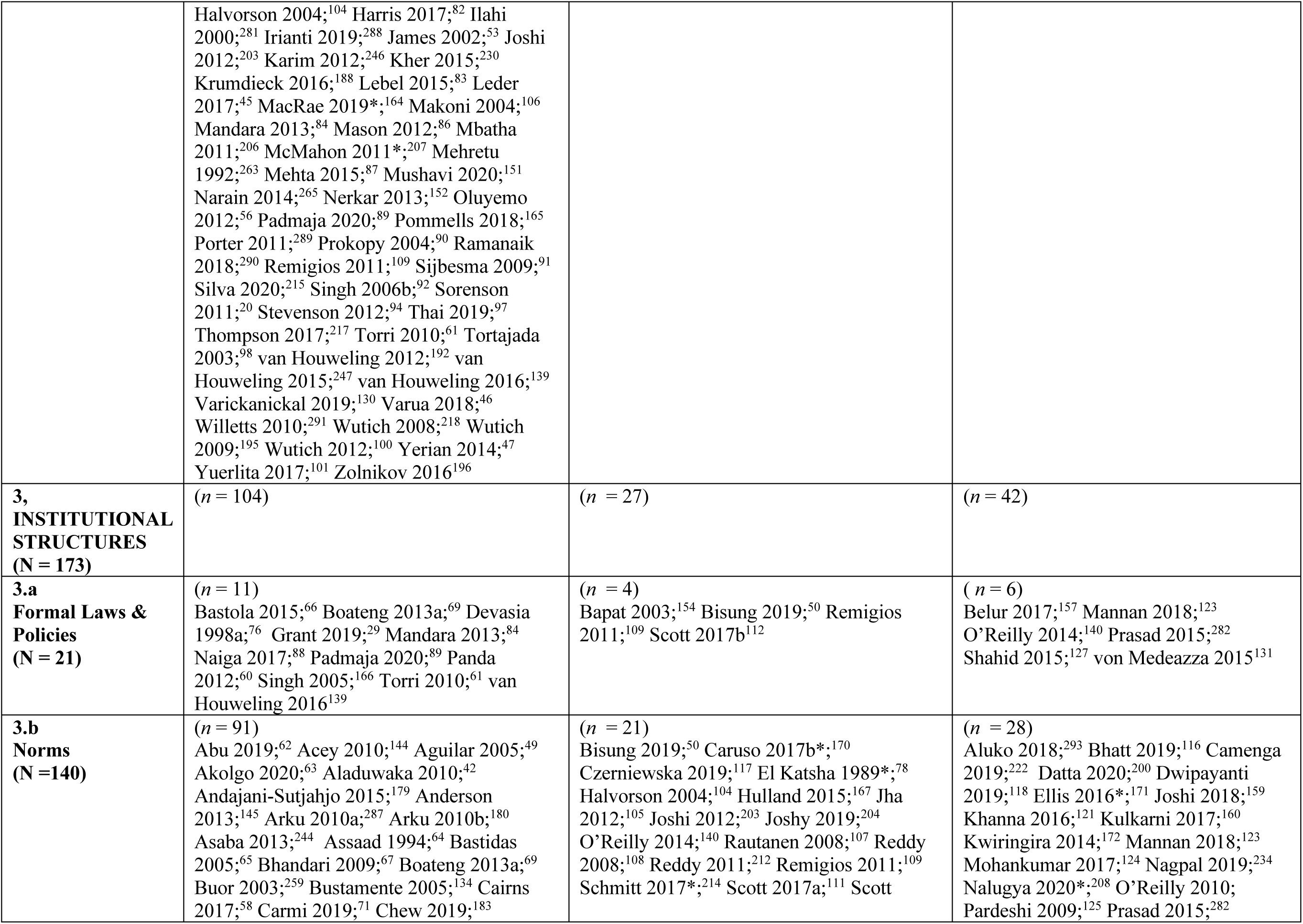
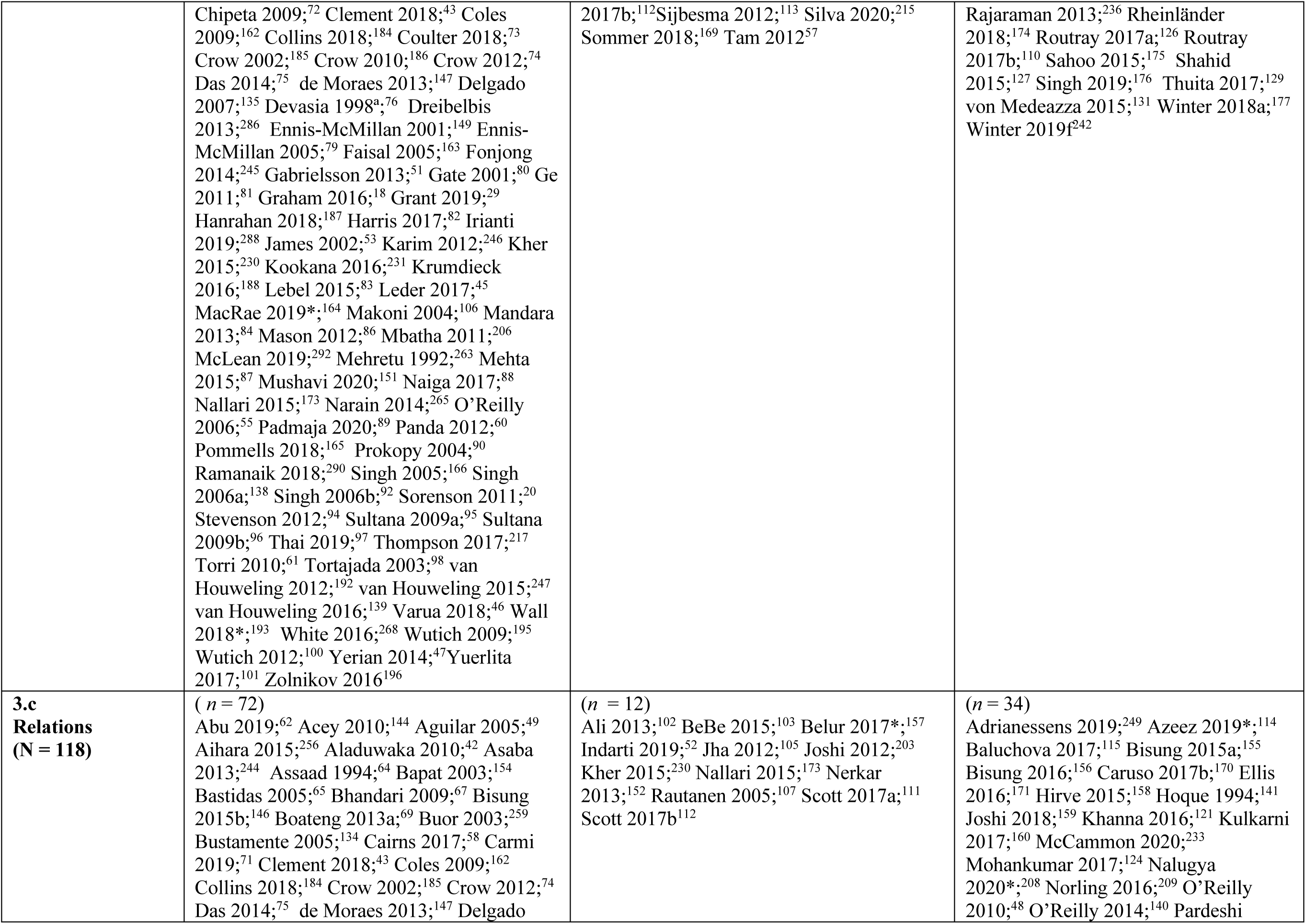
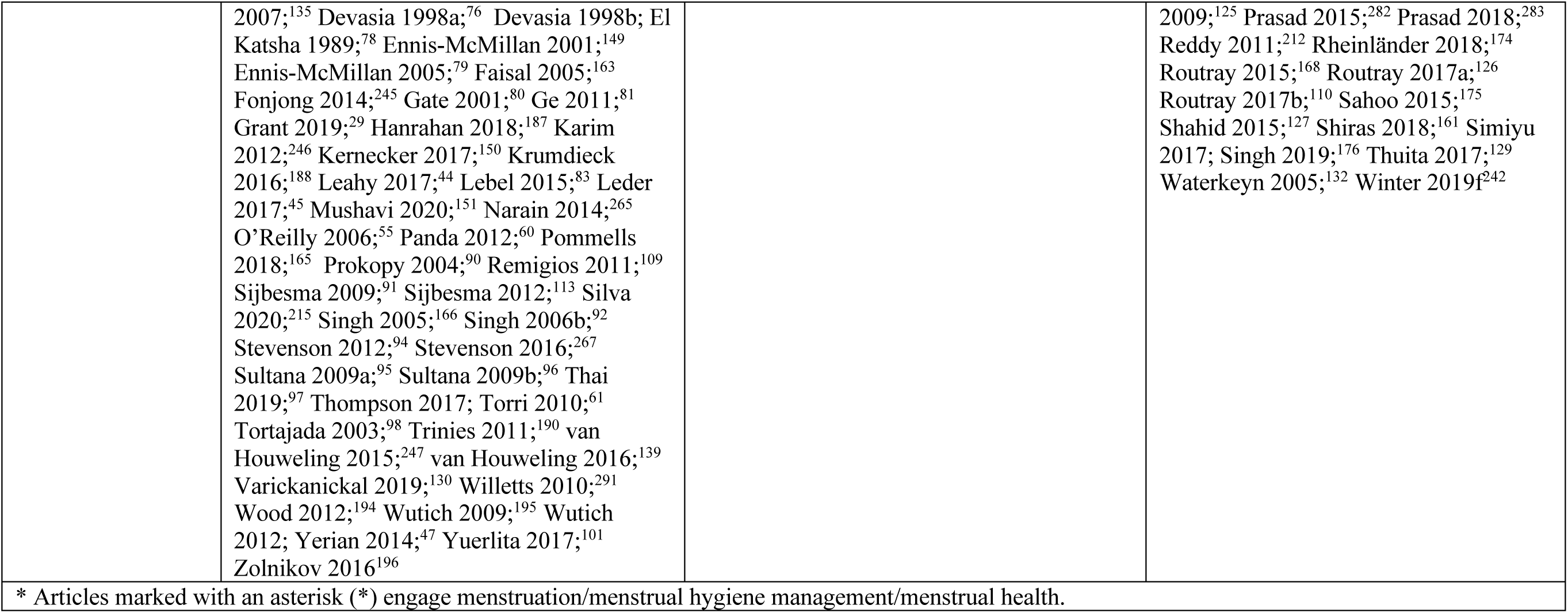
Summary of studies that engage water and/or sanitation by Sub-domains of empowerment. (N= 257)

## 1. Agency

As detailed below, women have reported exercising water and sanitation-related agency, including engagement in decision-making inside and outside the household, formal and informal leadership, and collective action. Still, women’s freedom of movement has minimally benefited from water and sanitation circumstances.

### 1.a Decision-Making

#### Household-level Decision-Making

Women have varied decision-making roles related to household water and sanitation. Women in Guatemalan savings groups reported having at least equal participation in household WASH decisions, with many reporting greater decision-making power than their husbands.^59^ Women in Bangladesh,^96^ Ethiopia,^94^ and India^46^ reported decision-making power over water collection and allocation, though some studies from India found men to make water collection decisions that did not account for women’s priorities.^87, 105^ A water security study from Nepal reported that some women found individual decision-making stressful and that seeking support from in-laws and husbands was culturally valued.^45^ With respect to sanitation, in India, the odds of having a latrine was significantly higher in households where women were the main decision makers^122^ and in Kenya, the likelihood of a household owning an improved sanitation facility was significantly higher when women had at least some input on decisions about major household purchases.^120^

Both the characteristics of women (e.g life stage) and the decisions themselves have influenced women’s involvement in WASH-related household decision-making. In Nepal^43^ and India,^53, 110^ younger and unmarried or newly married women typically had less decision-making power, and in rural Bangladesh,^96^, older women sometimes had greater decision-making power over household water collection. Women’s income-earning has enhanced their ability to make household water and sanitation decisions, particularly pertaining to small purchases or pay-per-use sanitation facilities.^29, 45, 91, 110, 133^ When large expenditures are involved, such as for latrine construction, women have been excluded from decision-making,^48, 55, 86, 97, 105, 109, 110, 117, 121, 128^ potentially resulting in latrines that fail to accomodate needs and thus remain unused.^48, 55, 110^ In Kenya, women reported limited input over home rental decisions, including which WASH services should be available.^130^

#### Extra-Household Decision-Making

Women have participated in a range of extra-household water and sanitation decision-making situations. In Indonesia, women exercised independent choice in selecting WASH-related jobs.^52^ In India, women became a part of decision-making processes in village-level sanitation meetings,^125^ water user committees,^75^ and other sanitation projects,^113^ and men believed women could speak up in meetings and expressed happiness that they could make joint decisions^131^. Women have voiced opinions about proposed activities and offered suggestions during informal water and sanitation meetings (Egypt),^64^ and both men and women noted that women are likely to speak out in public settings and take action on water-related issues given their role as household water managers (Mexico).^79^ Both men and women in Hmong communities in Vietnam agreed that women were likely to be listened to when they participated in water management meetings.^83^ In Bolivia, although women were rarely able to contribute in water-focused meetings, their contributions often shifted community conversations in important ways and women often had strong influence over water-related decisions, even when men had formal decision-making authority.^100^

Women also have been prevented from engaging in extra-household water and sanitation decision-making. Women reported being listed as water management group members, but never attending meetings (Bangladesh^95^), instructing husbands to relay water concerns to committees (Sri Lanka^42^), having husbands or sons attend meetings and make decisions in their place (India^92^), and deferring to men to speak during water-related meetings even though they bore water collection responsibility (Kenya^62^). In northern Kenya – where women primarily collect water for domestic use and men for livestock – women were not actively involved in decision-making around water source utilization and their needs were not prioritized.^47^ Women play increasingly greater roles in water committees in rural Malawi, but they remain dominated by men in urban areas.^72^

Several factors have influenced women’s involvement in extra-household water and sanitation decision-makng. Demographic factors, like class, caste, education, marital status, and age have been found to be influential.^45, 61, 67, 70, 84, 85, 89, 92, 95, 101, 102, 108, 111^ Various social factors constrain women’s involvement, like household responsibilities that conflict with meetings;^61, 62, 67, 69, 75, 88, 97, 101, 112, 119, 179^ women’s ability to attend but not speak or influence decisions;^61, 65, 100–102, 112^ fear, shyness, and lack of confidence compounded by social norms limiting attendance or ability to speak in front of men;^47, 58, 61, 62, 65, 67, 69, 75, 81, 83, 88, 95, 97, 101, 109, 112, 118, 119, 126^ lack of respect or interest in women’s opinion;^64^ and taboos governing the discussion of certain topics.^109^ Women’s lack of assets, especially land, has prevented entrée into decision-making spheres.^88^

#### Role of WASH Programs on women’s decision-making

Nongovernmental organizations (NGOs) have promoted women’s voices in WASH-related decision-making processes. They have encouraged women to speak up in culturally appropriate ways.^112, 291^ Trainings, information, and technical knowledge provided to women have contributed to the improvement of communication between men and women and led to women having a voice in household and community decision-making.^44, 61, 115, 291^ WASH programs have increased trust, acceptance, and respect for women as community decision-makers (India^115^ and Sri Lanka^42^), increased women’s roles in household decision-making (India^125^ and Vietnam^44^), and improved women’s reported ability to express themselves (Costa Rica^49^).

WASH programs also have discouraged women’s participation in decision-making by targeting men^88, 110^ or reinforcing gender norms.^57, 112^ In India, women were included in public forums because they are responsible for children’s health, while men were involved because they could demand improvements and travel outside the village.^112^ Additionally, women in India reported being provided water and sanitation, without consultation, resulting in services not meeting their needs.^108^ In Timor Leste, women’s participation in water and sanitation projects has been limited to activities like cleaning or preparing food.^57^

### 1.b. Leadership

Women have served as formal and informal leaders in water and sanitation initiatives. Formal positions include leadership on water and sanitation committees; leadership or management of WASH businesses, value chain companies, and organizations; leading community-level projects; and fundraising for and monitoring community-based water and sanitation initiatives.^45, 52, 69, 71, 75, 79, 98, 106, 112, 119, 125, 135, 140^ Women’s formal leadership has led to increased confidence among and toward women leaders.^42, 59, 112, 132, 143, 291^ In rural Vanuatu, water user committees with women in key posts met more regularly, functioned better, and collected more revenue than those with only men in these roles.^136^ Still, while quotas have resulted in more women in leadership positions,^137^ women remained underrepresented in committees and leadership.^69, 88, 123^ Documented informal leadership activities included arbitrating local disputes,^55^ disseminating water and sanitation information,^140, 146^ motivating family members to use latrines,^141^ and leading protests, like those during the “water wars” in Bolivia.^134^ Select water and sanitation programs in Costa Rica,^49^ Guatemala,^59^ India,^112, 131^ Sri Lanka,^42^ and Vietnam^44^ positively influenced the growth of women’s leadership and support for women leaders. In India, women’s participation in a community-led total sanitation campaign led to the emergence of female leaders who expanded their influence to other villages and activities.^131^

Several factors have influenced women’s engagement and acceptance in water and sanitation leadership positions, whether formal or informal. Individual-level factors like women’s marital status, religion, educational attainment, knowledge, age and social capital, including political ties and familial support, are influential.^45, 49, 52, 65, 81, 83, 85, 88, 135^ Barriers included limited confidence speaking in front of men or leading meetings;^60, 83, 112, 179^ constrained time due to responsibilities;^83, 179^ and relational factors, like men’s level of acceptance of female leaders.^42, 52, 69, 97, 112, 282^ In Mexico, women gained opportunities to serve as civil officers in municipal systems that govern piped water without opposition because the number of posts increased and men were not displaced.^79^ Even when leaders, women’s roles have sometimes been tokenistic, at lower levels than male peers, and not resulting in influential decision-making.^43, 46, 47, 65, 66, 69, 88, 90, 95, 119, 129, 137, 138, 179, 282^ In both Bolivia^58^ and India,^58, 90, 92^ women in formal leadership positions reported being represented by male relatives and husbands. Men usurping leadership roles in India^247^ have limited initiatives aiming to foster women’s leadership.

### 1.c. Collective Action

Women have engaged in varied forms of collective action to influence water and sanitation access, conditions, experiences, and opportunities. Women’s collective action has led to funding, demand, construction, repair, and maintenance of water services^42, 47, 59, 64, 75, 77, 111, 115, 149, 151, 154, 155, 179, 282^ and latrines.^115, 125, 133, 154, 156, 160, 161^ Women have organized to form lending groups to support women-led WASH businesses,^52^ provide community education about water safety,^42, 47, 59, 64, 75, 77, 111, 115, 149, 151, 154, 155, 179, 282^ monitor open defecation,^131^ and respond to sanitation-related harassment.^160^ Indigenous women in Bolivia and the U.S. have collectively defended water access.^134, 153^ Women’s involvement in WASH-related collective action has led to sustained involvement in water and sanitation issues^75, 149^ as well as collective action around non-WASH issues,^42, 112, 131^ and also has encouraged respect from community members, including men.^42, 75, 152, 291^

Myriad factors influenced collective action. Individual-level factors like class, caste, wealth, marital status, education, and interest in material benefits of participation influenced collective action invovlement.^68, 75, 111^ Facilitators of collective action included group solidarity, trust, and collective efficacy;^44, 52, 59, 75, 153, 160, 291^ the existence of other women’s groups, community groups, and unions;^75, 85, 155, 160^ women’s representation in community leadership;^59^ and spaces designated for women.^75^ Barriers to women’s involvement in collective action included hostility from men;^42^ norms that limit women speaking in front of men;^111, 126^ limitations on women’s participation by authority figures;^111^ opportunity costs when women have domestic and income-generating responsibilities;^62, 69, 75^ lack of incentive^55, 111^ or trust;^130^ limited guidance or training for engaging;^126^ and perceptions that actions are unlikely to have impact.^111, 155^

### 1.d. Freedom of Movement

Movement restrictions, which are most widely reported in India, influenced women and girls’ access to water ^87, 96, 164^ and defecation, urination, and menstrual management locations.^48, 121, 160, 167, 168, 170, 173, 175–177^ Research in India found that family members (often husbands, fathers, and in-laws) restricted water fetching and sanitation-related movements, most intensely for unmarried daughters^160, 168, 176^ and recently married and pregnant women,^164, 168, 175^ (less intensely for women of lower castes^121^ and widows^160^). Families have built latrines specifically to restrict movement,^48, 160^ though women do not always have access when needed if situated in spaces controlled by men.^48^ In India and Bangladesh, where women’s freedom of movement is restricted or being seen in public is considered shameful, women reported difficulty navigating public spaces for water or sanitation needs, experiencing stress, harm to reputations, or risked beatings.^87, 96, 173, 175^ To prevent shaming family members (Bangladesh^96^) or suffering beatings by men (India^87^), women collected water from closer, polluted sources rather than cleaner sources in male-dominated public spaces.

Women have described benefits of and preference for water and sanitation activities that require leaving the home. In Bangladesh, younger women enjoyed fetching water from distant water sources, providing opportunities to leave home and socialize.^96^ In rural India, some women who owned latrines preferred going for open defecation to visit friends and escape their home, mothers-in-law, and chores.^168^

Women’s freedom of movement influenced participation in community-level water and sanitation activities. Women in India,^75, 87^ Nepal,^45^ and Kenya^62^ reported needing permission from men or elders to participate in meetings outside the home,^62, 75, 87^ which limited their involvement in water and sanitation committees and access to information,^75, 87^ impacted their knowledge about water sources,^87^ and constrained water access and decision-making.^45, 87^ Restricted movement has limited women’s access to training opportunities and therefore engagement in piped water enterprises (Cambodia^29^), and has posed barriers to women’s WASH businesses (Indonesia^52^). Women in India experienced in development projects reported more spatial mobility and were more likely to participate in water committees.^75^ One program (India) intentionally aimed to enhance mobility by engaging women in sanitation promotion outside the home; yet, the very latrines promoted were found to increase women’s confinement.^48^

## 2. Resources

As described in the sections that follow, water and sanitation circumstances and conditions contribute to the deprivation of resources, particularly to bodily integrity, which includes health, safety and security, and privacy. Further, women’s control over resources, notably financial and productive assets, social capital, and time, influence their access to water and sanitation and, conversely, water and sanitation conditions influence access and control over resources.

### 2.a. Bodily Integrity

Bodily integrity includes safety and security, health, and privacy, a subdomain inductively identified.

Women’s and girls’ choices about and control over their bodies have been constrained due to water and sanitation conditions. Specifically, women limited bathing, and washing hands, clothes, menstrual materials, and bathrooms;^62, 72, 84, 86, 109, 130, 151, 183, 184, 186, 188, 215, 218, 267^ restricted food and water to avoid defecation and urination;^121, 154, 160, 165, 170, 178, 198, 210^ suppressed defecation and urination urges; ^2, 48, 167, 170, 174, 176, 177, 200, 203, 215, 221, 222, 224, 227, 233, 234, 236, 256^ delayed changing menstrual materials;^210, 226, 227, 233, 236^ and took anti-diarrheal medicines^160^ when lacking sufficient, safe or clean water and sanitation. Water improvements in Ethiopia reportedly decreased women’s economization and use of dirty water.^267^

Women’s ablity to meet their preferences is fundamental to ensuring bodily integrity. Multiple factors influence this ability, including conditions of water and sanitation facilities. Women and girls have described sanitation conditions as undesirable, dirty, disgusting, and nauseating^99, 121, 130, 154, 156, 157, 159, 171, 174, 203, 205, 206, 208–210, 212, 213, 215, 221–223, 226, 227, 230, 233, 237^ and lacking resources like disposal bins, soap, sufficient water, and buckets for washing or bathing, which are also needed during menstruation.^99, 130, 157, 171, 176, 189, 193, 201, 207, 208, 212, 213, 215, 232, 238–240^ Women used less preferred locations and sources for sanitation^116, 156, 159, 183, 205, 211, 215, 221, 226, 234, 241, 242, 256^ and water due to cost, distance, or access limitations.^56, 72, 76, 84, 151, 156, 163, 166, 180, 184, 185, 191, 194, 196, 203, 211, 215, 217^

#### 2.a.i Safety and Security

##### General Fears and Perceptions of Safety

General safety concerns were widely noted,^86, 105, 109, 125, 129, 133, 157, 163, 168, 172, 174, 177, 187, 188, 209, 212–215, 219, 224, 226, 227, 237, 241, 243^ including feeling unsafe collecting water,^86, 105, 163^ or using sanitation facilities at night,^154, 157, 176, 215, 241–243^ and when toilets lack doors, locks, lighting, or are far.^157, 176, 200, 209, 225–227, 238, 241^ Safety issues were described as particularly intense for adolescent girls, young women, and minorities.^2, 121, 160, 165, 167^

Research describing perceptions of improved safety all focuses on sanitation.^125, 208, 213, 243, 253^ Women in Nigeria with lockable latrines were significantly more likely to indicate that their latrine was safe than women without.^253^ Women and girls in Maharashtra, India, particularly when pregnant, menstruating, or adolescent, reported reduced fear when using toilets rather than open defecating.^125^ Women and girls have perceived sanitation facilities to be safer when there was lighting,^119, 243^ locks,^208^ a female caretaker,^213^ or an entrance shielded from the men’s side of a toilet block.^119^ In Uganda, a majority of female students who used toilets with lighting reported feeling safer, though many still felt unsafe going to the toilets alone because access paths remained unlit.^243^ Respondents in Kenya,^133^ Lebanon,^214^ and Ghana^221^ indicated that sex-separated latrines increase women and girls’ safety.

##### Harassment

General harassment, including taunting, teasing, name calling, and throwing things at women or girls is more frequently observed in sanitation^124, 154, 160, 161, 167, 173, 176, 249^ than water research.^154, 185^ Sexual harassment, including verbal harassment, peeping, flashing, and male masturbation is also more frequently documented in sanitation^157–160, 175, 176, 200, 203, 212, 217, 219^ than water research.^130, 200, 217, 245^ Women doing manual scavenging reported verbal abuse.^124^

##### Physical Violence

All research describing fear or experience of non-partner physical violence focused on sanitation, while all articles focusing on intimate partner physical violence and most focusing on interpersonal conflict focused on water.^47, 48, 62, 72, 75, 87, 96, 97, 109, 121, 129, 130, 157, 159–161, 163, 165, 167, 168, 174, 175, 177, 184, 187, 229, 237, 241, 243, 244, 246, 248, 250, 252, 255^

The majority of non-partner physical violence research focused on women’s general fear of physical attack by men or boys,^48, 72, 109, 121, 129, 157, 159–161, 163, 167, 168, 174, 175, 177, 184, 187, 237, 243, 255^ including “drunkards,”^177^ “bandits,”^161^ “idle youth”^241^ or “thieves”^241^ when accessing sanitation, with sanitation location emerging as important. Limited research documented actual experiences of physical violence from a non-partner. Research using Kenya Demographic and Health Survey data found women who defecated in the open had 40% greater odds of having experienced non-partner sexual and/or physical violence in the previous year compared to women who did not.^255^ In urban environments in sub-Saharan Africa, a greater association was found for women using a toilet shared with multiple households and experiencing non-partner violence than for women using private facilities.^248^ Women in India^121^ and Kenya^62^ shared stories of boys attacking and murdering girls who left home for defecation.

Inadequate water provision and collection behaviors were reported to influence intimate partner violence.^61, 87, 94, 97, 139, 151, 165, 184, 217, 246, 250, 252, 291^ Women reported violence from husbands if they did not provide water for bathing needs (Kenya,^184^ Ethiopia,^94^ Vietnam,^97^ Cameroon^217^), did not have water in the home (East Africa^165^), went to cleaner water sources considered too far (India^87^), did not prepare food on time because of water collection duties (Uganda^151^ and Mozambique^139^), or attended water-focused community meetings (India^61^). Due to an improved water source, a man in Vanuatu reported no longer hitting his wife because she stopped asking for his help to fetch water.^291^

Almost all articles documenting physical violence from interpersonal conflicts focused on water.^75, 96, 184, 229, 244^ Women in India experienced intrahousehold conflicts and beatings due to participation in a water supply project;^75^ young women in Bangladesh, especially new brides, suffered verbal and physical abuse from mothers-in-law when they failed to collect water quickly or did not collect enough;^96^ children and women in Uganda reported quarrels, including physical fights, while waiting in long water queues;^244^ and schoolgirls in Kenya reported that the school janitor would beat them if the toilet was unclean.^229^

##### Sexual Violence

Fear and experiences of sexual violence were reported by women and girls who had to leave home for water and sanitation needs.^56, 62, 94, 109, 121, 129, 139, 157, 159–161, 163, 165, 167, 169, 170, 172–175, 177, 184, 200, 217, 219, 224–226, 241, 244, 245, 247, 251, 254–256^ Research from India,^167, 254^ Kenya,^255^ and Nigeria^56^ found women openly defecating were particularly vulnerable to non-partner sexual violence compared to those using latrines. Fear of sexual assault was reported to be greatest at night,^109, 172, 224, 225, 241^ leading some women to use bags and buckets for sanitation needs.^161, 241, 256^ In India, those in urban settings reported greater fear of sexual assault when accessing sanitation than those in rural settings.^167, 175^ Qualitative research described how men in India hid to watch for, attack, and molest women openly defecting alone,^160^ and how a young woman in a Kenyan slum experienced an attempted rape when she walked ten minutes to the latrine.^225^ Some Kenyan women mentioned rape as a stressor (specifically during menstruation, which compels toilet use).^241^

Walking long distances and collecting water from certain sources reportedly exposed women and girls to sexual violence,^62, 139, 165, 217, 244, 245^ particularly as men studied women’s patterns.^165^ In refugee settings in Ethiopia and the Democratic Republic of Congo, caregivers advised girls to not bathe at the river or in community showers to avoid rape.^169^

#### 2.a.ii Health

Women reported different health concerns, mostly negative, based on life stage and circumstance. Notably, pregnant women expressed health fears related to sanitation-related superstitions (India^170, 175^), women with trouble controlling urination or defecation stressed about accessing sanitation sites (India^2^), women reliant on bags, buckets, or open defecation had substantially lower odds of reporting good health compared to other women (Kenya^242^), and circumcised women who had recently given birth found it difficult and painful to defecate and urinate (Kenya^129^).

##### Bodily Harm

Women and girls feared, risked, or experienced, varied harms related to their water and sanitation circumstances and activities.^48, 49, 64, 76, 87, 121, 127, 129, 130, 144, 154–156, 158–160, 163–165, 167, 168, 170, 172, 173, 175–177, 183–185, 187, 200, 212, 217, 230, 243, 244, 247, 262, 263, 282^ Many noted exacerbated experiences for women who were pregnant or elderly, had pre-existing conditions, and/or perform activities in harsh weather.^121, 129, 130, 155, 160, 163, 165, 167, 168, 170, 173, 175, 185, 217^

Reported risks of and actual harm linked to water and sanitation ranged widely, with reported water fetching related harms in particular resulting in serious and long-term consequences. Specific immediate water fetching harms included general and localized pain;^49, 76, 87, 109, 173, 184, 215, 217, 230, 262^ headaches and head injury;^215^ and injury from falls, trucks, and/or car accidents.^130, 144, 154, 184, 217^ Fears or experiences of harm from domestic and wild animals and insects were reported for both water collection and sanitation activities.^48, 121, 129, 156, 158–160, 163–165, 170, 173, 175–177, 184, 187, 200, 212, 215, 243, 244, 282^ Women and girls reported fear and actual experiences of injury at and when accessing sanitation locations, regardless of sanitation type.^121, 154, 158, 160, 170, 172, 175–177, 212, 243^ Women doing manual scavenging reported backaches (India).^127^

##### Illness and Infirmity

Women and girls’ experiences of illness and infirmity related to water and sanitation conditions are well documented.^49, 51, 56, 62, 67, 72, 97, 104, 108, 109, 121, 127, 130, 140, 154, 158–160, 163, 164, 170, 172, 174–177, 181, 183–185, 188, 194, 200, 206, 209, 211, 212, 217, 225–227, 233, 242, 245, 258, 259, 264, 270, 271, 273, 274, 276^

Water and sanitation access have been linked to women’s reproductive health outcomes. Two global studies (with data from 193^270^ and 144 countries^272^) and one focused on sub-Saharan Africa^273^ found increased access to improved water and sanitation to be significantly associated with decreased maternal mortality. In India, reproductive tract infections were more common among women who changed their menstrual materials outdoors rather than in a private room or latrine.^271^

Water and sanitation access have been linked to women’s nutrition and cardiovascular health outcomes. In rural Cambodia, women with nonimproved sanitation facilities had lower body mass index (BMI) and higher prevalence of anemia,^276^ and in urban India there was a positive correlation between women’s BMI scores and access to private toilets and a negative correlation between BMI and open defecation.^234^ Higher odds for anemia were observed among women with nonimproved drinking water sources in Uganda,^264^ and women in Kenya consumed poorer quality foods and a less diverse diet as a result of water insecurity.^184^ In Nepal, having intermediate and low water access was associated with higher systolic and diastolic blood pressure levels in women, while men’s blood pressure was not statistically different; elevated levels of blood pressure were highest for women with the least water access.^258^

Women qualitatively reported perceived linkages between various illnesses and their water and sanitation environments and experiences. Women reported experiencing or fearing diarrhea, vomiting, dysentery, cholera, hepatitis, schistosomiasis, and skin diseases due to polluted water,^51, 56, 72, 97, 130, 163, 164, 183, 185, 200, 206, 211, 217, 234, 245, 259, 274^ and experiencing diarrhea due to limitations on hygiene in times of water scarcity.^56, 109, 188^ Women in India worried about spreading disease if limited water constrained their ability to wash their hands during menstruation,^212^ and were concerned about fungal diseases when using dirty water inside sanitation facilities.^176^ When interventions improved water availability or treatment, women reported experiencing fewer vaginal infections^49^ and seeing fewer illnesses in their families.^67, 194^ Women who withheld food and water or suppressed urination and defecation when lacking access to safe, clean latrines reported experiencing urinary tract infections, headaches, stomach aches, constipation, diarrhea, and other illnesses.^121, 154, 158–160, 170, 176, 200, 234^ In India, women working in manual scavenging reported experiencing sickness, fever, and nausea.^127^

Women and girls identified multiple sanitation-related factors they linked to illness, infection, and disease spread. Factors included dirty public or school toilets,^62, 130, 154, 172, 177, 233^ open defecation fields,^56, 175, 176, 181^ feces within community spaces,^176^ foul odors from toilets,^174^ urinating on another person’s urine,^170^ using shared toilet seats at school,^209, 227^ consuming fish from canals where people defecated,^140^ unhygienic defecation practices and insufficient solid waste management,^104, 108, 163, 212, 225, 226^ and food contamination because of latrine proximity to cooking areas.^130^

##### Mental Health

Mental health is increasingly engaged in research on water^62, 76, 82, 86, 94, 109, 149, 151, 156, 163, 178, 184, 187, 188, 191, 195, 257, 260, 261, 266, 275^ and sanitation. ^2, 125, 154, 158, 159, 161, 164, 171, 174, 175, 215, 269, 275^

Water-related stress, anxiety, depression, or fear were reported by women in Ethiopia regarding water-related illnesses;^94^ in Bolivia,^218^ Brazil,^215^ India,^266^ Kenya,^62, 156, 184^ Mexico^149^ and Uganda^151, 191^ due to water insecurity; in Kenya^184^ and Canada^187^ due to interpersonal relations related to water; and in Bangladesh when collecting water after dark.^163^

Women reported myriad sanitation-related stressors, including fear of being attacked by men or ghosts,^163, 167, 170, 175, 215, 224^ being shamed for using open defecation sites,^158^ suppressing urination or defecation,^170^ withholding food and water,^170^ needing help to meet sanitation needs,^170^ being hurried while using shared facilities,^154, 171, 174^ sanitation-related costs,^156, 226^ and lacking privacy or being seen by others,^164^ especially men,^170, 175^ while openly defecating.^156, 158, 159, 170, 175, 224, 226^ Women working as manual scavengers in India reported feeling undignified and unworthy.^127^ Women coped with sanitation stressors by seeking social support, withholding food or drink, or changing the timing of sanitation behaviors, though adaptations could also cause stress.^154, 163, 170, 175^

Access to sanitation facilities was associated with mental well-being. Ownership of functional household latrines and enclosed bathing spaces were significantly associated with well-being among women in rural India.^2^ Toilet access in Kenya was associated with better mental health and well-being among women.^275^ Women who received latrines in Mozambique reported that they decreased their stress,^161^ and women in India reported that private latrines helped them to overcome the embarrassment, shame, and anxiety of open defecation.^125^

#### 2.a.iii Privacy

Women reported practicing hygiene behaviors at nonprivate water sources. Women and girls in Bangladesh,^185^ India,^170, 175, 212^ Indonesia,^101^ and Mozambique^161, 247^ described challenges obtaining privacy at water sources used for bathing, post-defecation cleansing, and washing, particularly during menstruation. In Mozambique, newly constructed handpumps provided women with easier, more reliable water access, but made achieving privacy for bathing and menstrual hygiene more difficult than at sources like rivers.^247^

Sanitation facilities often enable privacy. In India, women with latrines reported a greater sense of privacy,^125^ and adolescent girls and women found latrines particularly useful for maintaining privacy during menstruation and defecation, especially during the rainy season.^168, 208^ Women in Nigeria with lockable latrines were significantly more likely to report that their latrine was private (86% versus 64%).^253^

Privacy is challenging to obtain for those without sanitation facilities and for some using household or shared facilities. In urban India, privacy for open defecation is especially hard to obtain.^212^ Women in Nairobi, Kenya who rely on bags, buckets, or open defecation had lower odds of experiencing privacy.^242^ Household or shared toilets do not always prevent women from being seen or heard while defecating.^109, 170, 172^ Women reported privacy concerns in shared sanitation facilities in Zimbabwe,^109^ Mozambique,^161^ Kenya,^177^ India,^116^ and South Africa, where limited privacy at community ablution blocks was also noted.^237^ Privacy of household and shared latrines is compromised by broken^175^ or missing doors,^170, 234^ missing locks,^161, 177, 234, 237^ and poor construction.^161, 177, 226, 237^ Furthermore, women worried about being seen walking to latrines;^175, 213^ waiting in queues;^116^ having others hear or smell their activity;^161^ or the proximity of latrines to houses, main roads, or public spaces.^55, 161, 213^

Privacy concerns vary by gender and life stage. In India, women reported stricter privacy requirements for bathing than men,^203^ particularly among younger ^212^ or Muslim women.^140^ In Cameroon^217^ and India,^203^ women reported feeling less free than men to urinate or defecate openly due to privacy. In India, newly married women described stronger requirements for privacy due to reputation concerns,^121, 168, 175^ and older women reported going where privacy was compromised because they could not suppress urges.^159^

Women exercised various coping strategies to adapt to poor privacy conditions. They sought alternative locations by relieving themselves in containers in the home,^172, 177^ open defecating near the home^177^ or in sites protected by vegetation,^129, 156^ or by walking to farther defecation locations;^170^ suppressed needs by restricting eating and drinking and delaying urination, defecation,^48, 109, 121, 163, 167, 170, 174, 177, 234, 236^ and changing menstrual materials;^226^ created privacy for themselves by constructing or modifying structures for privacy^154^ or wearing skirts or dresses to create coverage;^215^ and responded to breaches in privacy when men passed by standing up (and soiling themselves) while open defecating.^121, 158, 160, 170, 212^

Insufficient sanitation privacy posed concerns for women and girls when away from home for work or school. In India, working women voiced concerns over doors without latches or a lack of facilities, resulting in the need to suppress lest they be seen by others, particularly men.^159, 236^ Similarly, Indian migrant women workers reported experiencing greater stress when openly defecating without privacy than from the risk of scorpion or snake bites.^158^ Girls in school reported insufficient privacy, particularly during menstruation.^99, 129, 171, 174, 189, 193, 197, 199, 202, 206–208, 216, 219, 233, 235, 238^ Where toilets were lockable and located away from boys’ toilets, girls experienced greater privacy.^174, 193, 197, 199, 202, 207, 208, 235, 239^ Girls have avoided eating and drinking during the school day to avoid using a sanitation facility (Phillippines^171^), brought friends with them to the toilets (Philippines,^171^ South Africa,^219^ and Sweden^209^), turned on the tap to prevent others hearing them (Sweden^209^), or chosen open defecation sites (Ghana^174^) to cope with privacy issues.

### 2.b. Critical Consciousness

The concept ‘critical consciousness’ was not explicitly engaged, though many engaged sub-constructs, including self-confidence and the identification and questioning of inequalities. Improved self-confidence, including willingness to speak up, was reported regarding women’s participation in WASH programs and campaigns in India,^75, 125, 131^ Sri Lanka,^42^ and Vietnam.^44^ Women in India reported improvements in self-confidence, dignity, and work and life circumstances due to improvements in water supply.^115, 265^ In Indonesia, some women reported feeling confident to challenge traditional gender roles and become leaders in the water sector.^52^

Awareness of their unequal position in society was reported to influence how women approached resolving water issues in Sri Lanka^42^ and Mexico,^149^ and sanitation issues in India.^131^ Women in rural India were aware of men’s unequal decision-making power, which was reinforced by NGOs; they noted that NGOs only approached male household heads as part of a national campaign to build household latrines.^110^ Women in Kenya raised complaints about service provision to government officials, but leaders ignored them because there are “no consequences for their inaction.”^130^

### 2.c Assets

#### 2.c.i Financial and Productive Assets

##### Financial and Productive Assets for Water and Sanitation Access and Participation

Access to financial and other assets impacted women’s water access. When women lacked control over income and assets, they had limited decision-making power over improving or accessing water sources.^51, 62, 72, 104, 109, 180, 185, 187, 244, 278^ Constrained finances limited women’s access to preferred water sources,^50, 72, 97, 144, 156, 180, 183–185^ desired water quantity,^72, 178^ and water treatment methods.^72, 97, 104, 190, 279^ Women reported spending money to access water sources with shorter wait times^47, 184^ or higher quality,^84, 184^ or to compensate male neighbors, laborers, or drivers for water collection.^151, 187^ Water was a major expense for many women.^51, 72, 97, 184, 186, 278^ In Uganda, women were four times more willing to contribute funds to water provision than men.^88^ In Kenya, purchasing water during the dry season limited money available for food.^184^ In Nigeria, women with water access issues found alternative sources, or purchased, bartered, or obtained water through credit.^144^ Furthermore, limited control over assets like carts, bicycles, and wheelbarrows made water collection more time-consuming,^84, 109, 244^ or, in sub-Arctic communities where women lack snowmobiles and guns for defense against bears, more dangerous.^187^

Limited income and asset access and control, including over land, constrained women’s access to sanitation^105, 110, 111, 121, 133, 154, 157, 160, 172, 173, 177, 203, 225, 226, 256^ and limited independent decision-making about latrine construction.^105, 110, 121^ Women reported using public pay-per-use toilets^154, 157, 160, 213, 225, 256^ and incurring higher costs than men due to more frequent need.^172, 177, 256^ Limited household income has forced women to prioritize needs like food over sanitation, whether investing in or paying to use toilets.^121, 133, 172^ Women in Kenya^226, 256^ and India^173^ adapted by using prefered pay-per-use locations less often or strategically, like for defecation only.^173, 226, 256^ Women have faced difficulties accessing government subsidies^111, 121^ or loans large enough for latrine construction^117^ and reported willingness to make financial sacrifices, like paying higher rent^177^ or accepting a lower wage job,^160, 203^ to gain toilet access. In India, even women with economic and decision-making power could not build latrines without land ownership.^110, 173, 176^ In Nairobi, women reported that greater financial stability and more control over resources would help them access sanitation.^133^

Finances and assets have influenced women’s representation or participation in community water and sanitation initiatives. Women have faced financial or asset-related barriers to participation in community-level initiatives, including a lack of land ownership for water committees (Peru^135^ and Uganda^88^), an inability to sustain required monetary contributions for self-help groups (India^58^), and a lack of financial incentive and opportunity costs (India^55, 75, 111^ and Ecuador^65^). In India, some families pushed women to attend watershed development meetings because of perceived monetary benefit.^58^

##### Water and Sanitation Income Generation

Water and sanitation have increased women’s income by providing job opportunities. Specifically, women have engaged in water vending,^75^ meter reading and water tank cleaning,^42^ water management,^29, 107^ water filter and toilet pan selling,^52^ and rainwater harvesting container construction.^107, 147^ They have also engaged in water-dependent income-generating activities, like horticulture or pottery-making,^45, 192, 247^ or sanitation-related activities, like toilet cleaning,^203^ latrine construction,^107^ serving as toilet attendants,^249^ or doing scavenging work.^124, 127, 283^ Finally, water and sanitation improvements have freed up time for other income-generating activities.^42, 53, 91, 160, 180, 196, 281^

Women’s income generation has been constrained by poor water access, limited employment opportunities, or by WASH initiatives themselves. Specifically, inadequate water access constrains income generation when time is needed instead for collecting water, or when water available is insufficient for income-generating activities.^29, 42, 53, 67, 72, 86, 91, 152, 160, 180, 184, 186, 188, 192, 196, 217, 230, 245, 281^ In some cases, women have not been able to get water and sanitation-related jobs, or have only had access to stigmatized or minimum wage jobs. In Kenya, women wanted to build toilets, but men got the contracts because of beliefs that the work was inappropriate for women, enabling men to earn and control local development.^129^ In India, women working as manual scavengers reported receiving little money, and relying on leftover food and used clothing for survival.^127^ A water project in Sri Lanka engaged women to work for free, but paid men involved to avoid conflict.^42^ Finally, water initiatives have threatened women’s incomes; in India, some women refused to support a water expansion project because it would impact their water vending income.^75^

##### Adverse Effects of Water and Sanitation Conditions on Financial Assets

Poor WASH conditions have indirect adverse effects on women’s financial assets, often by impacting health. Women reported expenses related to health seeking when they or their children contracted WASH-related illnesses (Thailand,^179^ India,^234^ and Kenya^226^), increased expenditures on pay-per-use toilets when sick and decreased wages when missing work to care for the sick (Kenya^225^), and lack of control over financial resources to invest in diarrheal disease prevention and treatment (Pakistan^104^). In India, women going for defecation feared being bitten by dogs, which could impact income due to doctor visits,^159^ and those lacking a place to change menstrual pads at work reported lost income from missing or leaving work to address needs.^236^ Conversely, in Costa Rica, a handpump project reduced women’s expenditures caring for the sick.^49^

#### 2.c.ii Knowledge and Skills

Women have specific household-level water and sanitation-related knowledge.^104, 110, 145^ In Kenya, women’s roles as primary water collectors instilled knowledge about water access, quality, and quantity.^62^ In Pakistan, mothers and grandmothers were a source of knowledge and influenced household decision-making about water and hygiene,^104^ while in indigenous communities in Canada and the U.S., older women were responsible for teaching younger generations about water-related responsibilities.^145, 153^

Women noted various types and sources of knowledge that influenced engagement in community-level activities. They reported the importance of knowledge related to village council processes to aquire toilet subsidies,^121^ community meetings and events to enable participation,^57, 90, 95, 101^ community members’ needs to evaluate water requests,^100^ and technical and business-related expertise to manage water enterprise businesses.^29^ Perceived limited knowledge has inhibited women’s participation in community decision-making bodies, collective action, and WASH businesses, or advantaged more educated women.^29, 60, 62, 65, 67, 68, 70, 83, 88–90^ Women reported acquiring knowledge from other women at public meetings about water issues (Vietnam^97^) and from close friends about WASH activities (Indonesia^52^).

Water and sanitation initiatives have enabled women to gain awareness, knowledge, or practical skills,^44, 49, 52, 55, 60, 69, 75, 80, 90, 107, 125–127, 131, 141, 143, 146, 243^ some doing so primarily to support project goals.^49, 60, 125, 131^ Women engaged in a program in Vietnam received education on WASH, technical skills, and women’s rights, which contributed to their confidence to speak up and negotiate household and community-level decisions.^44^ Women in Nepal received technical training in water and sanitation to enable job opportunities, however not all later found paid work outside the home.^107^

Training initiatives have upheld gender norms. In a project in Ghana, men received skilled technical WASH training, while women were trained in unskilled jobs like cleaners and hygiene officers.^69^ In India, an initiative attempted to exploit women’s knowledge of household water management and convince them to donate time and knowledge to the project.^55^

#### 2.c.iii Social Capital

Women formed and maintained social connections when collecting water^43, 64, 96, 101, 139, 265^ and relied on social capital from family, friends, and community members for multiple water needs. They leveraged social capital to access water when queues were long or water was scarce, ^47, 96, 100, 150, 151, 165, 184, 265^ jump water queues,^154, 165^ get water from private wells,^265^ haul water using carts,^215^ receive improved home water access,^45^ wait on water deliveries,^86^ or get water during menstruation.^164^ In India, water management was found to be a cooperative task with men collecting water while women made decisions about quantity, quality, and use.^166^ Male partners in Kenya^184^ and Mozambique^139^ helped with the physical labor or cost of water collection in rare circumstances, such as pregnancy, illness, or birth. In Kenya, women faced difficulties collecting water when they lacked childcare assistance.^130^ In Malawi, women were persuaded by friends and relatives to begin and maintain water treatment.^194^

Social capital has facilitated sanitation access. Women and girls have formed and maintained social connections when accessing sanitation locations,^116, 168, 175^ and sought social support to ensure privacy and safety and to protect reputations when urinating or defecating.^159, 168, 170, 173–176, 209, 212^ In India, schoolgirls asked friends to clean the school toilets so they could use them without fear of illness,^233^ yet women reported receiving scant sanitation-related cleaning support from men.^212^

Women accessed social networks to address water and sanitation problems.^51, 65, 76, 81, 121, 135, 144, 179^ They used social networks to lobby officials to solve water problems (Thailand^179^), raise complaints with local authorities about water issues (Ecuador^65^ and Nigeria^144^), leverage village council connections to access latrine construction subsidies (India^121^), and create strategic alliances with male household members to push their water-related priorities in the community (Bangladesh^95^).

Women leveraged social support to assume public roles. Women needed or sought family support to take public water management roles (Vietnam^83^), disseminate water and sanitation knowledge as community facilitators (Costa Rica^49^), become masons and water technicians (Nepal^107^), establish water enterprises (Cambodia^29^), participate in WASH activities and operate WASH businesses (Indonesia^52^), attend trainings on cistern building (Brazil^147^), and gain water user association membership (Peru^135^). In Bolivia, a lack of spousal support curtailed women’s participation in water and sanitation governance.^58^ Participation in water management projects helped women in Sri Lanka expand their social networks^42^ as did women’s participation in water-focused mobilizations in Bolivia.^134^ Some women in Indonesia chose jobs in the WASH sector and participated in WASH activities because they provide socializing opportunities.^52^

#### 2.c.iv Time

Women are the primary water-collectors globally,^18, 20^ devoting considerably more time to water-related tasks than men. Men were more likely to collect from closer sources,^20^ where queues were shorter^139, 247^ or they had priority access,^47^ or with assets like bicycles, motorcycles, wheelbarrows, or donkey carts to assist.^84, 109, 139, 244^ Seasonal changes impacted time spent on water acquisition: during the dry season, women waited for often unpredictable and therefore disruptive water deliveries (Philippines^86^); spent extra time collecting water when sources dried up (Ghana^183^); slept or cooked at sources when lines were long (Mozambique^247^); and walked long distances in the dark (Mozambique^139^). Women in Vietnam relied on water sources that became more time consuming to access due to climate-related shortages.^97^ Increasing urban population density in India has exacerbated water scarcity and time required for collection.^230^ Constraints on women’s time limit access to safe water or resources needed to treat or boil water.^78, 104, 183, 185, 194^ Women also spend considerable time caring for family members ill with water-related diseases.^49, 97, 163, 179, 225, 226, 234, 245, 291^

Women expended great amounts of time meeting sanitation needs and fulfilling sanitation-related responsibilities.^50, 52, 107, 108, 111, 113, 114, 118, 119, 121, 124, 127, 154, 159, 160, 168, 170, 171, 173, 175, 176, 181, 205, 212, 214, 222, 225, 226, 234, 236, 242, 251^ Women in India,^111, 181^ Kenya,^242^ and South Africa^251^ reported long walks to sanitation sites when they lacked private home toilets, and refugee women in Bangladesh^205^ reported waiting in long toilet queues. In urban India, women reported going for sanitation in early morning or late at night to avoid queues or ensure privacy,^154, 173, 176, 212, 234^ or open defecated rather than walking to and queuing at latrines.^154^ Time spent on sanitation has caused women to be late for work or suppress needs to avoid being late,^154, 159^ work longer days if they needed to leave to use the toilet,^236^ fail to complete household chores,^212^ or be scolded or punished for taking too long.^175^ Women spent time assisting others with sanitation-related needs^121^ and cleaning toilets, sometimes rising early to balance domestic duties with wage labor.^108, 113^ Women working as manual scavengers in India reported limited control over taking breaks, and not being able to take time off if ill or to participate in festivals, weddings, or other celebrations.^124, 127^

Opportunity costs exist related to time. Time devoted to water-related tasks limited women’s time for other household chores,^29, 42, 53, 67, 72, 86, 91, 139, 152, 160, 180, 184, 186, 188, 192, 196, 217, 230, 245, 257, 281^ rest and leisure,^76, 130, 183, 184, 230, 257^ and income-generating activities.^62, 108, 130, 184, 230, 257^ Women enlisted daughters to help with water or sanitation-related tasks,^62, 72, 87, 108, 109, 130, 151, 152, 163, 173, 230, 289, 290, 294^ or manual scavenging work,^124^ potentially impacting their daughters’ education. At schools in Swaziland, girls asked to collect water spent less time in class.^206^ Women’s participation was constrained in WASH businesses by restrictions related to overtime work (Indonesia)^52^ and in public life, such as water management, due to the time needed for household chores or income-generating activities, especially when participation is unpaid.^62, 65, 67, 83, 90, 92, 97, 101, 118, 119^

When women and girls have reclaimed time related to water and sanitation, they participated in leisure or rest,^74, 139, 168, 182, 247, 281, 282^ productive activities,^42, 53, 91, 97, 152, 160, 180, 182, 192^ education and vocational training,^42^ or other domestic chores.^43, 121, 139, 180, 247^ Time savings have resulted in improved relationships because women were better able to complete chores or spend time with family.^94, 139, 180, 196, 281, 291^ In Bangladesh, a tube well installation made water collection less time consuming, making older women more willing to help younger women with water collection.^163^

However, interventions have increased time burden; a water intervention in India prompted men, who previously bathed in public ponds, to demand that women fetch water for home bathing.^265^

## 3. Institutional Structures

As outlined below, institutional structures—including formal laws and policies, norms, and relations—influence women’s agency and resources related to water and sanitation.

### 3.a Formal Laws and Policies

Women have participated in WASH governance to varying extents. In Bangladesh,^163^ Ghana,^69^ India,^66, 92, 131, 166^ Kenya,^47^ and Uganda,^88^ governments and organizations that set up community water and sanitation committees have required the inclusion of women^47, 66, 69, 88, 92, 166^ or encouraged and supported women’s participation.^131, 163^ In one project in India, rules mandated women be on water committees, but women did not always know they were on them.^66^

Inclusion does not guarantee participation, voice, or decision-making power. Women’s participation in WASH-related governance has been constrained by a lack of awareness about the rules (Uganda^88^), a lack of transparency or distrust in political institutions (Kenya^73^), husbands or sons attending meetings in their place (India^92^), and being ignored by men (Kenya^47^). Committees with women have divided roles along gendered lines, with men in powerful positions, like president, and women in less powerful positions, like cleaner.^69, 139^ Research in Uganda found women constituted less than a third of members and were rarely in leadership positions on water user committees despite guidelines to have women comprise 50% of membership and serve in influential positions.^88^

WASH policies, and uneven policy awareness, implementation, and enforcement, have posed barriers to women. Female entrepreneurs in Cambodia found certain policies made it hard to run water enterprises.^29^ Even when policies exist to address women’s and girls’ needs, they are not always known or enforced. In India, both men and women lacked awareness of policies, acts, and regulations that prioritized gender equitable access to resources and participation in a watershed intervention.^89^ A comparative study in Tanzania, Ghana, Cambodia, and Ethiopia, found some countries mandated sex-segregated school toilets, but adherence was inconsistent.^239^ Lack of clear responsibility and accountability has resulted in policy or guidance failure. Women Sanitary Complexes in India were reportedly not maintained according to guidelines due to disagreements over responsibility.^125^ In refugee camps in Lebanon and Myanmar, a lack of detailed guidelines and clarity about responsibility affected government actors’ ability to provide sufficient water and sanitation for female refugees, particularly to support menstruation-related needs.^214^

### 3.b. Norms

Three themes were widely discussed related to norms: roles and responsibilities, restrictions, and shame and honor.

#### Roles and Responsibilities

Research discussed social norms that govern men’s and women’s gendered water- and sanitation-related roles, focusing largely on gendered division of labor and roles in public life. ^18, 20, 45, 47, 48, 52, 62, 69, 74, 75, 83, 87–89, 96, 97, 109–112, 117, 118, 121, 126, 129, 139, 151, 165, 170–173, 176, 179, 184, 187, 195, 200, 206, 215, 229, 231, 244, 247, 259, 265, 268, 282, 287, 288, 290–293^

Women and girls are largely responsible for household water collection.^18, 20, 48, 62, 74, 75, 88, 89, 96, 97, 109, 117, 139, 151, 165, 173, 179, 184, 187, 195, 215, 231, 247, 259, 268, 282, 287, 288, 290–292^ Providing sufficient water for the household was described as important to being a good wife; failure to do so has resulted in shame and violence.^139, 165, 259^ Normative roles extended beyond the home; girls at school in Swaziland were expected to fetch water rather than boys.^206^ Gender intersected with other social identities to result in further marginalization or disadvantage related to roles and expectations. In India, lower caste women were expected to give priority water access to women from upper castes, particularly when water was scarce,^265^ and were reported to lack access to handpumps constructed within upper caste areas, even when tasked with repairing them.^92, 138^ In Malawi, women with disabilities experienced difficulty collecting water, though they are often still expected to fulfill this role.^268^

Norms have influenced the extent that men engage in water collection and how they are perceived.^96, 184, 244, 265, 282^ Men in Uganda who assisted with water collection were deemed emotionally unstable or bewitched,^151, 244^ and boys in India were reportedly embarrassed to be seen helping girls carry water.^173^ In Rwanda, men preferred water collection over other traditionally female tasks, though still viewed it as a woman’s task.^292^ Norms of femininity and masculinity were found to be more fluid among lower castes in northwest India, enabling men to help women with water collection more easily than men in upper caste households.^265^ When men or boys do collect water, it has been more acceptable when they consider it ‘helping’ women^247^ or use technology – like bicycles, wheelbarrows, or motorcycles – while women and girls are expected to carry water.^109, 151, 215, 244, 247^ Exceptionally, in Inuit communities, men are primary water collectors. They carry guns for protection against polar bears near water sources – something unacceptable for women to do; women experienced anxiety accessing water when men migrate for work.^187^

Normative expectations affect women’s and girls’ sanitation-related practices. Rural Indian women reportedly suppressed urination and defecation urges when caregiving and household obligations were pressing^170^ and were often responsible for assisting others, including children, adolescent girls, and elderly family members, with sanitation needs.^48, 117, 121, 170, 176^ Women assumed more responsibilities for latrine cleaning than men because cleaning is typically considered women’s work.^172, 293^

Norms govern public water and sanitation participation. In several countries, it was more acceptable for men to participate in the public sphere and serve in leadership roles,^52, 62, 89, 97, 109, 111, 112, 118, 126, 129^ and to have technological jobs, like as handpump mechanics, while women were discouraged.^52, 69, 247^ These normative beliefs are sometimes supported by men, women, and NGOs. In Thailand, women believed men were better suited for public leadership roles, and men questioned women’s participation and problem-solving capacities related to village water resources.^179^ In sanitation planning initiatives in Kenya, a man justified women’s exclusion from participation in the sanitation initiatives stating, women’s brains were “like that of a child.”^129^ NGOs have targeted men as household heads and ignored women in decision-making and public participation.^110^ In rare cases, cultural beliefs facilitated women’s public participation; women were described as more trustworthy on water user committees in Uganda and Ghana.^69, 88^

#### Restrictions

Normative societal and familial rules have defined the boundaries of acceptable water and sanitation-related behaviors for women.^47, 48, 52, 73, 75, 87–89, 96, 97, 110–112, 116, 126, 160, 163–165, 167, 168, 173–176, 183, 204, 220, 247^ Norms related to women’s movement (discussed in the ‘Freedom of Movement’ section), asset ownership, and menstrual status have influenced women’s water and sanitation access, behaviors, and participation in public life. Specifically, women’s restricted access to property in rural India required them to seek permission from husbands or fathers-in-law to access land for latrine construction.^110^ In Nepal, norms preventing women from sharing latrines with men has compelled open defecation.^116^ Women in India reported not being able to use sanitation facilities^204^ or touch water when menstruating due to perceptions that menstruation is polluting.^164, 167^

Restrictive social norms regarding women’s movement and roles have limited women’s attendance and participation in water governance and repair, and latrine construction. In Cambodia^29^ and Indonesia,^52^ female entrepreneurs reported that norms related to women’s freedom of movement and household roles made their engagement in piped water enterprises difficult. Even when they had technical skills to repair handpumps, women who lacked privilege and social access in India were not able to physically access handpumps to service them.^138^ In some communities in India, Kenya, and Uganda, women were prevented from speaking or sitting with men in public;^47, 88, 111, 126^ women who did participate in water- and sanitation-related public life could be punished for violating these proscriptions, whether scolded, beaten, gossiped about, or assumed to be neglecting children and household responsibilities.^47, 73, 88, 112^ Research from Kenya found that contracts for latrines and water facilities were almost exclusively awarded to men since technical work was not considered appropriate for women.^129^ In rural India, separate sanitation meetings were sometimes held because men and women could not sit and speak together in meetings,^126^ while in urban India, where norms were less restrictive, women have been able to mobilize and participate in water supply projects.^75^

#### Shame and Honor

Women and girls have experienced sanitation- and menstruation-related shame; norms related to shame and honor have been leveraged to change water and sanitation conditions and behaviors.^48, 109, 125, 126, 131, 159, 160, 165, 167, 168, 170, 172, 174, 175, 177, 199, 215, 216, 236^

Women and girls voiced shame related to sanitation and menstruation behaviors and experiences. Specifically, shame was reported regarding open defecation,^125, 159, 174, 215, 236^ particularly when their bodies may be exposed to males,^48, 167, 168, 170, 175, 236^ and related to menstruation, including when seen carrying^199^ or washing and drying menstrual materials,^170^ or if menstrual blood was visible on toilets^174^ or hands.^216^ As a result of sanitation- and menstruation-related shame, newly married women in India reported stress about their reputations within the family, while unmarried women and girls worried about their reputations outside the family, family honor, and marriage prospects.^160, 167, 170, 175^ Women have also reported reducing water intake during work hours to avoid the shame of asking employers for sanitation access.^159^ Finally, women and girls in India have experienced shame and stigma when blamed for experiencing sanitation-related sexual assault.^160^

Norms related to shame and honor have been leveraged, perpetuated, and exploited in attempts to improve water and sanitation conditions.^48, 55, 96, 126, 160^ Women have convinced husbands to invest in tube wells within their compounds to prevent inappropriate movement through public spaces (Bangladesh^96^) and to construct latrines to prevent daughters from potentially engaging in clandestine relationships when going for open defecation (India^160^). Households in India have built new sanitation facilities to protect the reputations of their daughters and daughters-in-law,^160, 168^ with encouragement from national-level sanitation campaigns messages about respecting women and girls’ privacy,^125, 131^ the impact of women’s privacy on family status,^48^ and the shame of women exposing themselves.^126^ Latrines have been marketed in India as means of confining women to the household, thus elevating the public status of the family.^48^

#### Shifting Norms

Water and sanitation-related norms, including roles, responsibilities, and restrictions, have shifted, but impacts are variable. Water initiatives have increased expectations and work for women: the introduction of piped water resulted in women having to fetch water for their husband’s bathing needs (India)^265^ and doing all the clothes washing, instead of sharing responsibility (Vietnam)^44^. Some water and sanitation initiatives have shifted norms and expectations in women’s favor.^42, 111, 112, 291^ WASH projects in Vanuatu resulted in men increasingly assisting with responsibilities like cooking and hygiene.^291^ In Sri Lanka, a women’s group’s successful water project demonstrated women’s capacity to conduct public WASH projects.^42^ In India, NGOs used facilitators to challenge norms that limited women’s ability to speak in village health committees that address WASH issues.^111, 112^ However, initiatives that diverged from normative roles have faced resistance. When a project attempted to put household water connections in women’s names, women objected (India).^75^ Men with new homestead water access did not like their increased involvement in what they perceived as women’s work (Kenya).^74^ Norm change has emerged from changing circumstances; some displaced Syrian refugee girls did not face the same menstrual restrictions they did back home, though female refugees from Myanmar did not experience any shift.^214^

### 3.c Relations

Relations have both facilitated and hindered water-related behaviors and experiences. In India, social networks played an important role in women’s decision to purchase new water filters^190^ and aided women’s water collection when queues were long.^154^ In Bangladesh,^96^ India,^265^ and Uganda^151^ women relied on social networks to access water, but noted that relationships could be strained or unreliable when water was scarce. Women avoided quarrels at collection sites in Kenya by waking early to get water.^62^

Relations have also facilitated and hindered sanitation-related behaviors and experiences. Women frequently accompanied one another or asked men to accompany them for safety when open defecating,^121, 159–161, 168, 173, 175, 237^ and to avoid harassment from community members when going to latrines and open defecation sites after dark.^121, 129, 154, 168, 176, 214, 224^ Additionally, women in Kenya reported quarrels over contributions toward sanitation activities and maintenance of shared toilets,^156^ and female toilet attendants in Europe reported poor treatment by patrons who refused payment.^249^ Relations influenced sanitation-related school experiences; girls reported bullying from boys if they were known to have defecated in the school toilet (Sweden^209^) and if boys knew girls were menstruating because of which toilets they used (India^233^).

#### Change Agents and Gatekeepers

Actors with whom women and girls have relationships can serve as change agents, who deviate from the status quo and enhance women and girls’ empowerment, or gatekeepers, who maintain the status quo and constrain women and girls’ choice and voice.

Few articles discussed change agents. In Nepal, encouragement from mothers-in-law or husbands influenced women’s decisions to take leadership positions in water-user groups.^45^ In a few households in rural India, men asked younger women – like daughters-in-law – for input on issues like latrine site selection, though this was uncommon.^110^ In Cambodia^29^ and Indonesia^52^, familial and organizational support helped women establish and manage WASH enterprises.

Within the home, men were often gatekeepers, holding household decision-making power. Indian women brought up toilet construction, but men dismissed it as an unnecessary expense.^121^ Even higher status women, like mothers-in-law, deferred to husbands or income-earning sons to make latrine construction decisions.^110^ Women in some cases need or value permission from in-laws or husbands to participate in activities outside the home, including water user groups and water management committees^45, 62, 97^ and have faced discouragement from men when seeking to engage in collective action.^52^

Outside the home, women encountered various gatekeepers. Women are gatekeepers to other women; in India, upper caste women have scolded or punished Dalit women seeking to bathe in the same area.^175^ Men in Sri Lanka expressed hostility to women’s leadership in repeated complaints to the police that the women leading a water project had to address.^42^ Women reported that they faced criticism and confusion from local leaders and members of their communities when they sought to engage in WASH-related work (Indonesia).^52^ Students in Uganda who feared using the school toilets reported that male teachers would embarrass and deny them passes to go home when they started their periods.^208^

#### Influence of water and sanitation on relations

Water and sanitation conditions have strained intra-household relations. Women and men reported getting angry with family members over water issues (Bolivia),^195^ and women reported feeling bad about unreasonable water collection expectations (Uganda)^151^ and disputes occurring when husbands used too much water for bathing or when children spilled water (Kenya).^47^ Husbands became angry, quarrelsome, or physically abusive when women did not complete chores or meals on time due to water collection demands,^94, 139, 151, 165^ when there was not enough water available for bathing and other purposes,^94, 97, 165, 184, 217^ or when they were asked to assist with water collection.^291^ In Mozambique, women reported that long absences from home for water collection could lead to conflicts because husbands suspected infidelity.^139^ Dependence on others for water assistance could result in stress and anxiety for women in Mexico^149^ and Inuit communities in Canada.^187^ In India, women reported that their participation in a water project led to intrahousehold conflict; some women reported experiencing beatings when they spoke up at meetings.^75^ In rural India, sanitation-related conflicts varied by life stage. Adolescent girls were scolded by parents for taking too long or talking to boys when going for urination/defecation; newly married women were scolded for not following household sanitation rules;^175^ women with children were scolded for abandoning children to meet sanitation needs; and women across life stages worried about upsetting others if they asked for assistance meeting sanitation needs.^224^

Water and sanitation conditions have strained inter-household relations. Disputes with neighbors at water collection locations were widely reported.^47, 94, 130, 151, 154, 165, 175, 184, 195, 244^ Water collection sites could be places of tension and danger, including sexual exploitation, for women and girls in Zimbabwe^109^ and Uganda.^151, 165^ In Kenya, disputes arose among women competing for water access, and verbal and physical fights erupted when women collecting payments favored women in the queue.^47^ Inter-household sanitation-related conflicts were reported related to shared toilets,^173^ where women practice open defecation,^175, 176^ and accessing sanitation facilities.^161^

Water and sanitation initiatives have both negatively and positively affected relations. Negative effects in India included increased expectations of wives to bring water home, quarrels, and conflicts at water points.^265^ Positive effects included increased respect and support for women by men, changed division of labor, and increased ability of women to negotiate with husbands (Vanuatu^291^); reduced conflict between husbands and wives (Mozambique^139^, Kenya,^74^ and Vanuatu ^291^); improved relationships between men and women (India^91^) and in families (Kenya^196^ and Vietnam^44^); heightened status of women in and outside the home and positive attitudes and support for women’s collective work (India^42^); and greater acceptance of women performing WASH roles outside the home (India^115^).

## Discussion

We synthesized evidence on water, sanitation, and women and girls’ empowerment from 257 peer-reviewed empirical research articles, resulting in the most comprehensive synthesis of gender-focused water and sanitation research to date. Our review is more expansive than recent WASH and gender or empowerment-focused reviews that restricted inclusion based on publication date,^31^ only extracted data from titles and abstracts,^31^ or focused more narrowly on the intersection of gender, water, and health^32^ or on identifying the dimensions of empowerment used in WASH research^30^. The majority (86%) of included research was from Africa and Asia and focused on adults (69%). No studies focused primarily on men or transgender or non-binary individuals, revealing patterns of research inequities that should be addressed to include more diverse geographies and populations. Our search and synthesis was grounded in an existing model of empowerment,^33^ which we iteratively expanded based on our review to include ‘freedom of movement’ within the Agency domain and ‘privacy’ within the Resources domain. Agency was the least commonly engaged domain among included articles (122; 47%); the Resources domains was dominant (241; 94%). This review not only contributes insights to those working in the WASH sector, it also augments understanding of the role of water and sanitation in discourse on empowerment and gender equality more broadly.

Water and sanitation research that engages empowerment and related domains is extensive and growing, yet clear conceptualization of empowerment remains limited. We found that 82% (211) of included articles were published since 2010, providing evidence that aligns with anecdotal observations that empowerment is increasingly a focus in water and sanitation research.^27, 31^ Both the large body of work identified and its growth, which is consistent with growth in the broader field of women’s empowerment,^295^ further justify the need to assess learning to date. Importantly, 124 (48%) included articles did not use ‘empowerment’ or related words (e.g. “empower”) in their text, but were included because they specifically reported on empowerment sub-domains (e.g safety, decision-making) aligned with our guiding model. Of the 133 (52%) articles that did use empowerment or related terms in their text, only 17 (13%; or 7% of total) provided a definition or conceptualization of women’s empowerment to inform their work. Our assessment aligns with a reflection by Dery et al. (2019) that the definition of empowerment in WASH research lacks clarity.^30^ As this line of research continues, it is essential that researchers use empowerment-related terminology deliberately. Without solid conceptualization, whether by articulating a clear guiding definition or framework a priori, or by eliciting a local understanding of empowerment via the research (See Bisung and Dickin (2019)^50^), terms may be misused or become devoid of meaning.^296^ The tool we created and used (Figure 1) can be adopted to assess engagement of empowerment concepts in future research.

We identified only one article that used a quantitative tool to assess women’s empowerment related to WASH, the Empowerment in WASH Index.^297^ Additional tools are under development, like the Agency, Resources, and Institutional Structures for Sanitation-related Empowerment (ARISE) Scales,^298^ and the Water, Sanitation, and Hygiene Gender Equality measure (WASH-GEM).^299^ Additionally, in a perspective piece, Kayser et al. (2019) propose four priority areas for assessing gender equality and empowerment related to WASH.^24^ The momentum around measuring empowerment and WASH, while exciting, merits caution. In her critical review of current practices for measuring women’s empowerment, Richardson (2018) provides three recommendations to ensure sound measurement: measures should draw on theory; analytic methods should be used to minimize implicit judgement and bias; and comprehensive information should be collected (like data from men or complementary qualitative data).^296^ Yount et al. (2018) note that empowerment measures should rigorously assess validity and reliability.^300^ Furthermore, research on empowerment from other sectors teaches that a multitude of measures can also hinder learning by limiting comparability between studies,^301^ and that tools should be selected intentionally based on context and need.^302^ If WASH-related empowerment is to be monitored at scale, a consensus on indicators and associated measures will be needed eventually. As tools emerge, there will be a need to review and assess the similarities, differences, strengths, weaknesses, specific uses, and gaps to continue strengthening this line of research.

Although existing measurement is limited, this review illuminates how water and sanitation circumstances and conditions have resulted in myriad negative impacts to women’s well-being that remain unmonitored and under-evaluated. Illustratively, considerable research shows how compromised water and sanitation environments have contributed to women’s risk or experience of harassment or physical and sexual violence; ^47, 48, 56, 62, 72, 75, 87, 94, 96, 97, 109, 121, 124, 129, 130, 139, 154, 157–161, 163, 165, 167–170, 172–177, 184, 185, 187, 200, 203, 212, 217, 219, 224–226, 229, 237, 241, 243–252, 254–256^ compromised mental well-being;^2, 62, 76, 82, 86, 94, 109, 125, 149, 151, 154, 156, 158, 159, 161, 163, 164, 171, 174, 175, 178, 184, 187, 188, 191, 195, 215, 257, 260, 261, 266, 269, 275^ resulted in illness, infirmity, and bodily harm^48, 49, 51, 56, 62, 64, 67, 72, 76, 87, 97, 104, 108, 109, 121, 127, 129, 130, 140, 144, 154–156, 158–160, 163–165, 167, 168, 170, 172–177, 181, 183–185, 187, 188, 194, 200, 206, 209, 211, 212, 217, 225–227, 230, 233, 242–245, 247, 258, 259, 262–264, 270, 271, 273, 274, 276, 282^ or limiting of hygiene, food, and water;^62, 72, 84, 86, 109, 121, 130, 151, 154, 160, 165, 170, 178, 183, 184, 186, 188, 198, 210, 215, 218, 267^ and suppression of urination, defecation and menstrual hygiene needs,^2, 48, 167, 170, 174, 176, 177, 200, 203, 210, 215, 221, 222, 224, 226, 227, 233, 234, 236, 256^ among other impacts. However, estimates of the burden of inadequate WASH remain focused on disease outcomes.^8^ Our review shows that the true burden of inadequate WASH on well-being is likely far greater, supporting calls to collect and report sex-disaggregated and gender-specific data that also considers intersectionalities,^303, 304^ and to not discount or ignore impacts predominantly or only experienced by women and girls.^305^ Finally, this review confirms the need to set goals for measuring, monitoring, and reporting the specific impacts of WASH on women and girls.^22^

Despite the numerous studies that have documented impacts of water and sanitation conditions on women and girls, there has been scant action in response, warranting greater investment in programs and evaluations to create and assess change. For example, the Safety and Security domain is well-researched; 82 (32%) articles document links between WASH and safety and security yet only one study evaluated a program that improved perceived safety.^243^ Furthermore, WASH programs have the potential to catalyze change across domains and be transformative, but only a few studies have documented these linkages and transformative change. Research in Ghana showed how a water project resulted in time savings, which further benefited well-being among women.^180^ A water intervention in Kenya also enabled time savings, which led to improved intra-familial relationships.^196^ WASH programming in Vietnam provided knowledge and skills, which women reported improved their decision-making, public participation, and relations.^44^ Programs aiming to achieve transformative change should be evaluated to deepen the knowledge base on WASH and empowerment. To start, more research on Agency is warranted. Agency is “the heart of empowerment” and transformation,^33^ yet is the domain least studied. Research should aim to understand Agency in the context of WASH, including its relationship with other domains. Finally, political will and investment are necessary to ensure women and girls are prioritized in WASH initiatives. At a minimum, WASH initiatives should engage women meaningfully and not cause harm by bypassing them.^88, 108, 110^

### Strengths and Limitations

This review has leveraged an existing model of empowerment to review full texts of an extensive volume of research, resulting in the most comprehensive synthesis on gender and water and sanitation to date. This review does not capture studies not available in English and research has continued to emerge since the search, somewhat limiting the comprehensiveness. Additionally, other models of empowerment exist that could have framed the review. Still, this review provides a valuable base and resource from which to develop programming and further research. While this review captures research on menstruation, it only does so in the context of water and sanitation research and cannot be considered a comprehensive review of menstruation and empowerment. Hygiene was not a focus and should be considered in future reviews. Finally, while we intended to register the review, we began extraction to inform other work prior to registering and recent revisions to guidelines now stipulate extraction should not have started prior to registration and thus we were no longer eligible.

### Conclusion

Water and sanitation research specifically engaging women’s and girls’ empowerment in a well-defined or conceptualized manner is limited. However, a substantial body of research examining domains and sub-domains of empowerment exists that should be leveraged to develop and evaluate programs focused on improving the life outcomes of women and girls. Importantly, the integration of a gender lens into WASH research, and program and policy planning and evaluation, can enable the identification of inequities and potential harms and benefits,^303^ and should be mainstreamed across the WASH sector^23^. Emergent discussions about ‘transformative WASH’ call for interventions that radically reduce fecal contamination^306^ and chemicals^307^ to impact health. However, our findings underscore that a more comprehensive ‘transformative WASH’ that includes gender-transformative approaches to challenge and reduce systemic constraints on women’s and girls’ resources and agency is not only warranted but long overdue.

## Supporting information

Supplemental Materials: Water, Sanitation, and Women's Empowerment: A systematic review and qualitative metasynthesis

## Data Availability

All data produced in the present work are contained in the manuscript

## ACKNOWLEDGEMENTS

We are grateful for the artistic contributions of Fernanda Kuri who assisted the authors in adapting the empowerment conceptual model by Van Eerdewijk et al. (2017) to be specific to this work (Figure 2).

This work was supported by the Bill & Melinda Gates Foundation (OPP1191625). Under the grant conditions of the Foundation, a Creative Commons Attribution 4.0 Generic License has already been assigned to the Author Accepted Manuscript version that might arise from this submission.

## Notes

### Competing Interest Statement

The authors have declared no competing interest.

