## Supplemental Materials: Water, Sanitation, and Women's Empowerment: A systematic review and qualitative metasynthesis for "Water, Sanitation, and Women’s Empowerment: A systematic review and qualitative metasynthesis"

**Supplemental Table 1: PRISMA Checklists for (a) Abstract and (b) Full Manuscript**

| 1a. PRISMA Checklist for Abstracts | | | |
|---|---|---|---|
| **Section and Topic** | **Item #** | **Checklist item** | **Reported (Yes/No)** |
| **TITLE** | | | |
| Title | 1 | Identify the report as a systematic review. | Yes |
| **BACKGROUND** | | | |
| Objectives | 2 | Provide an explicit statement of the main objective(s) or question(s) the review addresses. | Yes |
| **METHODS** | | | |
| Eligibility criteria | 3 | Specify the inclusion and exclusion criteria for the review. | Yes |
| Information sources | 4 | Specify the information sources (e.g. databases, registers) used to identify studies and the date when each was last searched. | Yes |
| Risk of bias | 5 | Specify the methods used to assess risk of bias in the included studies. | Yes |
| Synthesis of results | 6 | Specify the methods used to present and synthesise results. | Yes |
| **RESULTS** | | | |
| Included studies | 7 | Give the total number of included studies and participants and summarise relevant characteristics of studies. | Yes |
| Synthesis of results | 8 | Present results for main outcomes, preferably indicating the number of included studies and participants for each. If meta-analysis was done, report the summary estimate and confidence/credible interval. If comparing groups, indicate the direction of the effect (i.e. which group is favoured). | Yes |
| **DISCUSSION** | | | |
| Limitations of evidence | 9 | Provide a brief summary of the limitations of the evidence included in the review (e.g. study risk of bias, inconsistency and imprecision). | Yes |
| Interpretation | 10 | Provide a general interpretation of the results and important implications. | Yes |
| **OTHER** | | | |
| Funding | 11 | Specify the primary source of funding for the review. | Yes |
| Registration | 12 | Provide the register name and registration number. | No |

| 1b. PRISMA Checklist for Full Manuscript[1] | | | |
|---|---|---|---|
| **Section and Topic** | **Item #** | **Checklist item** | **Location where item is reported** |
| **TITLE** | | | |
| Title | 1 | Identify the report as a systematic review. | Title |
| **ABSTRACT** | | | |
| Abstract | 2 | See the PRISMA 2020 for Abstracts checklist. | Abstract table above |
| **INTRODUCTION** | | | |
| Rationale | 3 | Describe the rationale for the review in the context of existing knowledge. | Intro section |
| Objectives | 4 | Provide an explicit statement of the objective(s) or question(s) the review addresses. | Final intro paragraph |
| **METHODS** | | | |
| Eligibility criteria | 5 | Specify the inclusion and exclusion criteria for the review and how studies were grouped for the syntheses. | Study eligibility section in Methods |
| Information sources | 6 | Specify all databases, registers, websites, organisations, reference lists and other sources searched or consulted to identify studies. Specify the date when each source was last searched or consulted. | Search strategy section in Methods |
| Search strategy | 7 | Present the full search strategies for all databases, registers and websites, including any filters and limits used. | Search strategy section in Methods and Supplemental Table 2 |
| Selection process | 8 | Specify the methods used to decide whether a study met the inclusion criteria of the review, including how many reviewers screened each record and each report retrieved, whether they worked independently, and if applicable, details of automation tools used in the process. | Study eligibility section in Methods |
| Data collection process | 9 | Specify the methods used to collect data from reports, including how many reviewers collected data from each report, whether they worked independently, any processes for obtaining or confirming data from study investigators, and if applicable, details of automation tools used in the process. | Analysis section in Methods |
| Data items | 10a | List and define all outcomes for which data were sought. Specify whether all results that were compatible with each outcome domain in each study were sought (e.g. for all measures, time points, analyses), and if not, the methods used to decide which results to collect. | Analysis section in Methods |
| | 10b | List and define all other variables for which data were sought (e.g. participant and intervention characteristics, funding sources). | Analysis |

| 1b. PRISMA Checklist for Full Manuscript[1] | | | |
|---|---|---|---|
| **Section and Topic** | **Item #** | **Checklist item** | **Location where item is reported** |
| | | Describe any assumptions made about any missing or unclear information. | section in Methods |
| Study risk of bias assessment | 11 | Specify the methods used to assess risk of bias in the included studies, including details of the tool(s) used, how many reviewers assessed each study and whether they worked independently, and if applicable, details of automation tools used in the process. | Study quality appraisal section in Methods |
| Effect measures | 12 | Specify for each outcome the effect measure(s) (e.g. risk ratio, mean difference) used in the synthesis or presentation of results. | N/A |
| Synthesis methods | 13a | Describe the processes used to decide which studies were eligible for each synthesis (e.g. tabulating the study intervention characteristics and comparing against the planned groups for each synthesis (item #5)). | N/A |
| | 13b | Describe any methods required to prepare the data for presentation or synthesis, such as handling of missing summary statistics, or data conversions. | N/A |
| | 13c | Describe any methods used to tabulate or visually display results of individual studies and syntheses. | N/A |
| | 13d | Describe any methods used to synthesize results and provide a rationale for the choice(s). If meta-analysis was performed, describe the model(s), method(s) to identify the presence and extent of statistical heterogeneity, and software package(s) used. | Analysis section in Methods |
| | 13e | Describe any methods used to explore possible causes of heterogeneity among study results (e.g. subgroup analysis, meta-regression). | N/A |
| | 13f | Describe any sensitivity analyses conducted to assess robustness of the synthesized results. | N/A |
| Reporting bias assessment | 14 | Describe any methods used to assess risk of bias due to missing results in a synthesis (arising from reporting biases). | N/A |
| Certainty assessment | 15 | Describe any methods used to assess certainty (or confidence) in the body of evidence for an outcome. | Analysis section in Methods |
| **RESULTS** | | | |
| Study selection | 16a | Describe the results of the search and selection process, from the number of records identified in the search to the number of studies included in the review, ideally using a flow diagram. | First paragraph in Results |
| | 16b | Cite studies that might appear to meet the inclusion criteria, but which were excluded, and explain why they were excluded. | Flow diagram in results |
| Study characteristics | 17 | Cite each included study and present its characteristics. | First paragraph in Results and Supplemental |

| 1b. PRISMA Checklist for Full Manuscript[1] | | | |
|---|---|---|---|
| **Section and Topic** | **Item #** | **Checklist item** | **Location where item is reported** |
| | | | Table 3 |
| Risk of bias in studies | 18 | Present assessments of risk of bias for each included study. | Study Quality section in Results |
| Results of individual studies | 19 | For all outcomes, present, for each study: (a) summary statistics for each group (where appropriate) and (b) an effect estimate and its precision (e.g. confidence/credible interval), ideally using structured tables or plots. | First paragraph in Results and Table 3 |
| Results of syntheses | 20a | For each synthesis, briefly summarise the characteristics and risk of bias among contributing studies. | N/A |
| | 20b | Present results of all statistical syntheses conducted. If meta-analysis was done, present for each the summary estimate and its precision (e.g. confidence/credible interval) and measures of statistical heterogeneity. If comparing groups, describe the direction of the effect. | N/A |
| | 20c | Present results of all investigations of possible causes of heterogeneity among study results. | N/A |
| | 20d | Present results of all sensitivity analyses conducted to assess the robustness of the synthesized results. | N/A |
| Reporting biases | 21 | Present assessments of risk of bias due to missing results (arising from reporting biases) for each synthesis assessed. | N/A |
| Certainty of evidence | 22 | Present assessments of certainty (or confidence) in the body of evidence for each outcome assessed. | N/A |
| **DISCUSSION** | | | |
| Discussion | 23a | Provide a general interpretation of the results in the context of other evidence. | Throughout Discussion |
| | 23b | Discuss any limitations of the evidence included in the review. | Discussion, second to final paragraph |
| | 23c | Discuss any limitations of the review processes used. | Discussion, second to final paragraph |
| | 23d | Discuss implications of the results for practice, policy, and future research. | Throughout Discussion |
| **OTHER INFORMATION** | | | |
| Registration | 24a | Provide registration information for the review, including register name and registration number, or state that the review was not | Not |

| 1b. PRISMA Checklist for Full Manuscript[1] | | | |
|---|---|---|---|
| **Section and Topic** | **Item #** | **Checklist item** | **Location where item is reported** |
| and protocol | | registered. | registered |
| | 24b | Indicate where the review protocol can be accessed, or state that a protocol was not prepared. | Not published |
| | 24c | Describe and explain any amendments to information provided at registration or in the protocol. | N/A |
| Support | 25 | Describe sources of financial or non-financial support for the review, and the role of the funders or sponsors in the review. | End of manuscript |
| Competing interests | 26 | Declare any competing interests of review authors. | End of manuscript |
| Availability of data, code and other materials | 27 | Report which of the following are publicly available and where they can be found: template data collection forms; data extracted from included studies; data used for all analyses; analytic code; any other materials used in the review. | Will make endnote database available upon publication |

[1] Page MJ, McKenzie JE, Bossuyt PM, Boutron I, Hoffmann TC, Mulrow CD, et al. The PRISMA 2020 statement: an updated guideline for reporting systematic reviews. BMJ 2021;372:n71. doi: 10.1136/bmj.n71

**Supplemental Table 2: PubMed Search Strategy[1].**

| | |
|---|---|
| **Search 1: Empowerment** | "institutional structures" or "bodily integrity" or "critical consciousness" or assets or “collective action” or “gender norms” “community norms” or “family norms” or “gender relations” or “family relations” or “community relations” or power or empowerment or empower or leadership or agency or autonomy or equity or equality or decision-making |
| **Search 2: WASH** | "toilet facilities" [MeSH] or "hygiene"[ MeSH] or toilet or latrine or sanitary or "sanitation"[MeSH] or wash or washing or bath or bathe or "baths"[MeSH] or bathing or "urination"[MeSH] or urinat* or feces or faeces or fecal or "menstrual hygiene products" [MeSH] or menstrual or menstruation or “water collecting” or “water collection” or “collecting water” or “water fetching” or “fetching water” |
| **Search 3: Women** | "Women"[Mesh] or women or woman or girl* or "gender identity"[mesh] or gender or sex or female* |
| **Search 4: Exclusions** | "Animals"[Mesh] NOT ("Animals"[Mesh] AND "Humans"[Mesh]) |
| **Final search** | 1 AND 2 AND 3 AND 4 |

1. In MEDLINE and EMBASE, a search string was employed to eliminate animal-only studies

### Supplemental Table 3: Included Studies and Key Characteristics.

<table>
<tr><th>Study ID</th><th>Study Type</th><th>Data Type</th><th>Methods</th><th>Country</th><th>Location</th><th>Sample Size</th><th>Participants</th><th>Empowerment Findings</th><th>Water, Sanitation, and Menstruation Findings</th><th>Quality Appraisal</th><th>Tier of Empowerment</th></tr>
<tr><td colspan="12"><u>KEY</u><br><br>Study Type: Int. (intervention); Obs. (observational)<br><br>Data Type: Qual (qualitative); Quant (quantitative)<br><br>Methods: CI (cognitive interview); FGD (focus group discussion); IDI (in-depth interview); KII (key informant interview); PRA (participatory rural appraisal)<br><br>Empowerment Findings: BI (bodily integrity); S (safety); H (health); P (privacy); F (financial resources); SC (social capital); T (time); K (knowledge); CC (critical consciousness); DM (decision-making); FOM (freedom of movement); CA (collective action); L (leadership); R (relations); N (norms); FLP (formal laws and policies)<br><br>Water, Sanitation, and Menstruation Findings: M (menstruation); S (sanitation); W (water)<br><br>Quality Appraisal: range = 0-5; NA indicates the type of study or article was not suited to appraisal using the MMAT; * indicates that the study is an RCT, and the maximum score is 4<br><br>Tier of Empowerment: range = 1 – 4</td></tr>
<tr><td>Abrahams 2006</td><td>Obs.</td><td>Qual</td><td>FGD; IDI; observation; PRA</td><td>South Africa</td><td>urban, semi-rural</td><td>81</td><td>81 adolescent school girls (aged 16+)</td><td>BI; S; P</td><td>S</td><td>5</td><td>2</td></tr>
<tr><td>Abu 2019</td><td>Obs.</td><td>Qual</td><td>KII</td><td>Kenya</td><td>rural</td><td>12</td><td>12 community leaders, members, and WASH stakeholders</td><td>BI; H; S; F; T; K; DM; FOM; N; R</td><td>S; W</td><td>5</td><td>3</td></tr>
<tr><td>Acey 2010</td><td>Obs.</td><td>Mixed</td><td>household surveys; interviews</td><td>Nigeria</td><td>urban</td><td>787</td><td>783 surveys and 4 interviews with adult women and men</td><td>H; S; F; T; SC; CA; N; R</td><td>W</td><td>3</td><td>-</td></tr>
<tr><td>Adriaenssens 2019</td><td>Obs.</td><td>Mixed</td><td>survey; IDI</td><td>Belgium</td><td>urban</td><td>117</td><td>117 adult male and female toilet attendants</td><td>S; F; R</td><td>S</td><td>5</td><td>2</td></tr>
<tr><td>Agesa 2019</td><td>Obs.</td><td>Quant</td><td>secondary data analysis: (Kenya Integrated Household Budget Survey)</td><td>Kenya</td><td>country-wide</td><td>807,843</td><td>807,843 male and female youth (14 – 23 years)</td><td>T</td><td>W</td><td>4</td><td>-</td></tr>
</table>

| | | | | | | | | | | | |
|---|---|---|---|---|---|---|---|---|---|---|---|
| Aguilar 2005 | Int. | Qual | observation, anthropological methods | Costa Rica | rural | NR | male and female adults | BI; H; CC; F; T; SC; K; DM; L; N; R | W | NA | 4 |
| Aihara 2015 | Obs. | Quant | Survey | Nepal | urban | 372 | 372 married adult women (18-60 years) | H; R | W | 4 | - |
| Aihara 2016 | Obs. | Quant | Survey | Nepal | urban | 267 | 267 adult postnatal women (mean age = 26) | BI; H; F; T | W | 5 | - |
| Akolgo 2020 | Obs. | Mixed | Survey; IDI | Ghana | rural | 167 | 167 adult household heads/representatives and key stakeholders | DM; N | W | 3 | 2 |
| Aladuwaka 2010 | Int. | Qual | KII; FGD; IDI; observation | Sri Lanka | rural | NR | NR | CC; F; T; SC; K; DM; L; CA; N; R | W | 5 | 4 |
| Alam 2017 | Obs. | Quant | Survey; facility inventory | Bangladesh | country-wide | 2332 | 2,332 adolescent school girls (11 – 17 years) | BI | M; S | 4 | 1 |
| Ali 2013 | Int. | Qual | FGD; IDI; observation; PRA | Bangladesh | Rural | NR; at least 14 | 14 interviews with adult men and women, WASH staff, committee members (FGD, PRA, and observation participants not reported) | DM; R | S; W | 5 | 2 |
| Aluko 2018 | Obs. | Quant | Survey | Nigeria | urban | 312 | 312 household heads/most senior adult member of the household (mean age = 46.1) | N | S | 4 | - |
| Andajani-Sutjahjo 2015 | Obs. | Qual | Observation; FGD; IDI | Thailand | peri-urban | 132 | 132 adult men and women (30 – 61 years) | BI; F; T; N | W | 5 | - |
| Anderson 2013 | Obs. | Qual | IDI | Canada | NR | 11 | 11 adult indigenous women in positions of responsibility | K; CA; N | W | 5 | - |
| Anyarayor 2019 | Obs. | Mixed | Survey; IDI | Ghana | NR | 353 | 353 adolescent school girls in boarding facilities | BI; S; P | S | 3 | 1 |
| Arku 2010a | Int. | Qual | FGD; IDI | Ghana | rural | 45 | 45 married adult women, their husbands, and single women | T; N | W | 5 | - |

| | | | | | | | | | | | |
|---|---|---|---|---|---|---|---|---|---|---|---|
| Arku 2010b | Int. | Qual | FGD; IDI; observation | Ghana | rural | 340 | 340 married adult women and their husbands (ages 20+) | BI; F; T; N | W | 5 | - |
| Asaba 2013 | Obs. | Mixed | Survey; FGD; KIIs; observation | Uganda | rural | NR; at least 612 | 602 surveys, 10 FGDs, and KIIs with household heads (including child-heads) and key water actors | H; S; T; N; R | W | 4 | - |
| Assaad 1994 | Int. | Mixed | KAP surveys; PAR; observation; IDI; FGD | Egypt | rural | NR | adult women | BI; H; CC; F; T; SC; K; DM; L; CA; FOM; N; R | W | 5 | 2 |
| Azeez 2019 | Obs. | Qual | IDI | India | rural | 30 | 30 women and girls of reproductive age (16 – 45 years) | H; S; P; F; T; SC; DM; FOM; R | S | 5 | - |
| Baker 2017 | Obs. | Quant | Survey | India | rural | 4020 | 4,020 women and girls (14 – 45 years) | BI; H; T; | S; W | 5 | - |
| Baluchova 2017 | Int. | Mixed | Survey; IDI; observation | India | rural | 58 | 58 household heads | CC; DM; CA; R | S | 3 | 2 |
| Bangdiwala 2004 | Obs. | Quant | Survey | Chile, Egypt, India, Philippines | urban | 3975 | 3,975 women and girls (15 – 49 years) | S | S | 4 | - |
| Bapat 2003 | Obs. | Qual | IDI | India | urban | NR | adult women | BI; H; S; P; F; T; K; CA; FOM; R; FLP | S; W | NA | - |
| Barchi 2020 | Obs. | Quant | Survey | Burkina Faso, Cameroon, Comoros, Cote d'Ivoire, DRC, Gabon, Ghana, Kenya, Liberia, Malawi, Mali, Mozambique, Namibia, Nigeria, Sierra | country-wide | 138,097 | 138,097 women of reproductive age (15 – 49 years) | S | S; W | 4 | - |

| | | | | | | | | | | | |
|---|---|---|---|---|---|---|---|---|---|---|---|
| | | | | Leone, Tanzania, Togo, Uganda, Zambia, and Zimbabwe | | | | | | | |
| Bastidas 2005 | Obs. | Mixed | FGD; IDI; participatory activities | Ecuador | rural | 60 | 60 adult men and women | H; F; T; SC; K; DM; L; CA; FOM; N; R | W | 2 | - |
| Bastola 2015 | Int. | Mixed | FGD; survey; PRA | India | rural | 248 | 248 adult women | DM; L; FLP | W | 3 | 3 |
| BeBe 2015 | Obs. | Quant | survey | Pakistan | urban | 1329 | 1,329 adult women, local public officials | DM; R | S; W | 4 | - |
| Belur 2016 | Obs. | Mixed | Survey; FGD; IDI | India | urban | NR; at least 148 | 142 surveys, 6 IDIs, and FGDS with adult women (ages 18+) | BI; S; F; CA; FOM; R; FLP | M; S; W | 4 | 2 |
| Bhandari 2009 | Int. | Mixed | Survey; FGDs; RRA | Nepal | rural | 140 | 140 adult women | H; P; F; T; K;DM; L; CA; N; R | M; S; W | 2 | 3 |
| Bhatt 2019 | Obs. | Qual | FGD; IDI | Nepal | rural | 35 | 20 IDIs and 15 FGD participants, adult men and women who practice open defecation | BI; P; CC; SC; DM; N | S | 5 | - |
| Bisung 2014 | Obs. | Quant | Survey | Kenya | rural | 452 | 452 adult household heads/representative (ages 15+) | K; DM; CA | W | 5 | 1 |
| Bisung 2015a | Obs. | Qual | IDI; photovoice | Kenya | rural | 8 | 8 adult women (22 – 54 years) | H; K; CA; R | S; W | 5 | 2 |
| Bisung 2015b | Obs. | Qual | FGD; IDI; photovoice | Kenya | rural | 8 | 8 adult women (22 – 54 years) | H; T; SC; K; CA; R | W | 5 | 1 |
| Bisung 2016 | Obs. | Qual | FGD; KII | Kenya | rural | NR; at least 19 | 9 KIIs and 10 FGDs with adult women and men (aged 18+) | BI; H; P; F; T; CA; R | S; W | 5 | - |

| | | | | | | | | | | | |
|---|---|---|---|---|---|---|---|---|---|---|---|
| Bisung 2018 | Int. | Quant | Survey | Kenya | rural | 557 | 557 adult household heads/representative (18 – 90 years) | BI; F; T; | W | 5 | - |
| Bisung 2019 | Obs. | Qual | participatory methods | Ghana and Burkina Faso | rural; urban | 58 | 58 WASH community stakeholders and local government officials | F; T; K; DM; L; N; FLP | S; W | 5 | 4 |
| Boateng 2013a | Int. | Mixed | FGD; IDI; survey | Ghana | rural | 297 | 297 adult household heads/representatives (mean age = 41.8) | F; T; K; DM; L; N; R; FLP | W | 3 | 3 |
| Boateng 2013b | Int. | Mixed | FGD; survey | Ghana | rural | 295 | 295 adult household heads/representatives (mean age = 41.8) | K; DM | W | 3 | 2 |
| Boateng 2018 | Obs. | Mixed | Cognitive interviews; surveys | Kenya | urban, peri-urban, rural | 251 | 10 CIs and 241 surveys with post-partum women (18 – 39 years) | F; T | W | 5 | - |
| Boosey 2014 | Obs. | Mixed | Survey; IDI; FGD; toilet assessment | Uganda | rural | NR; at least 158 | 140 surveys, 6 FGDs, and 12 IDIs with adolescent schoolgirls (13 – 16 years), teachers | BI; P; K | S; W | 4 | 2 |
| Bora 2016 | Obs. | Mixed | IDI; FGD; survey | India | urban | 30 | 30 adult female police officers | BI | S; W | 2 | |
| Brewis 2019 | Obs. | Quant | Survey | Nepal | nation-wide | 14739 | 14,739 adult household heads/representatives | H | W | 4 | - |
| Buor 2004 | Obs. | Quant | Survey | Ghana | urban | 210 | 210 adolescent girls and adult women (ages 12+) | H; T; N; R | W | 4 | 2 |
| Bustamente 2005 | Obs. | Qual | | Bolivia | urban | NR | adult women | CC; SC; L; CA; N; R | W | NA | 2 |
| Cairns 2017 | Int. | Mixed | FGD; IDI; observation; survey | Bolivia, Lesotho, India | urban, rural | NR | key water users, development workers, community members | F; T; SC; DM; L; CA; N; R | W | 3 | 3 |
| Camenga 2019 | Obs. | Qual | FGD | United States of America | urban | 360 | 360 adolescent schoolgirls, adult women (11 – 93 years) | BI; P; CC; T; N; R | S | 5 | - |
| Carmi 2019 | Obs. | Mixed | Survey; IDI | Palestine, Lebanon, and Jordan | NR | 42 | 42 adult women working in the water sector | S; CC; T; K; DM; L; N; R | W | 3 | 3 |

| | | | | | | | | | | | |
|---|---|---|---|---|---|---|---|---|---|---|---|
| Carolini 2012 | Obs. | Qual | IDI | Mozambique | peri-urban | 27 | 27 household representatives (mostly female household heads) (18 – 80 years) | BI; H; F; T; K | S; W | 2 | - |
| Caruso 2017a | Obs. | Mixed | FGD; IDI; survey | India | rural | 1523 | 1,523 married and unmarried women, elderly women (aged 18+) | BI; H; S; P; F | S | 3 | - |
| Caruso 2017b | Obs. | Qual | IDI; FGD | India | rural | 115 | 115 married and unmarried women, elderly women (18 – 75 years) | BI; H; P; T; FOM; N; R | M; S; W | 5 | 2 |
| Caruso 2018 | Int. | Quant | Survey | India | rural | 1347 | 1,347 married and unmarried women, elderly women (18 – 100 years) | H; S; | S | 5 | - |
| Cheng 2012 | Obs. | Quant | NR | 193 countries | country-wide | NR | adult household heads/representatives | H | S; W | 5 | - |
| Chew 2019 | Obs. | Qual | IDI; KII; observation | Ghana | rural | 30 | 30 adult women (aged 18+) | BI; H; F; T; K; N | W | 5 | - |
| Chipeta 2009 | Obs. | Qual | Participatory methods | Malawi | urban, peri-urban | NR | adolescent girls, adult women | BI; H; S; F; T; DM; N | W | 5 | 2 |
| Clement 2018 | Int. | Qual | FGD; IDI; KII; observation; participatory methods | Nepal | rural | NR; at least 57 | adult men and women; 11-12 women and men engaged in village mapping, 2-3 in transect walk, 22 household interviews, 13 KIIs, 9-12 participants in FGD | F; T; SC; DM; L; N; R | W | 5 | 4 |
| Coles 2009 | Obs. | Qual | Case studies | Sudan, India, Nepal | rural | NR | NR | FOM; N; R | W | NA | 3 |
| Collins 2019 | Obs. | Qual | IDI; participatory methods including go along interviews | Kenya | | 40 | 40 pregnant and post-partum women | BI; H; S; F; T; SC; N; R | W | 5 | - |
| Connolly 2013 | Obs. | Qual | KII; observation; participatory methods | Cambodia | urban, rural | 161 | 161 girls in and out of school (16 – 19 years), adult key informants | BI; P | M; S; W | 5 | 2 |
| Cooper-Vince 2017 | Obs. | Quant | Survey | Uganda | rural | 808 | 257 adult women caregivers of children aged 5-17 (mean age = 33.5); 551 children (mean age = 9.2) | H | W | 5 | - |

| | | | | | | | | | | | |
|---|---|---|---|---|---|---|---|---|---|---|---|
| Cooper-Vince 2018 | Obs. | Quant | Survey | Uganda | rural | 1603 | 1,603 adult men and women (aged 18+ and emancipated minors 16 – 18 years) | H | W | 5 | - |
| Corburn 2015 | Obs. | Mixed | Survey; FGD; mapping data | Kenya | urban | NR; at least 650 | 650 surveys and FGDs with household representatives | BI; H; S; F; T | M; S; W | 3 | - |
| Corburn 2016 | Obs. | Mixed | FGD; survey; spatial data | Kenya | urban | NR; at least 650 | 650 surveys and FGDs with adolescent schoolgirls, adult heads of household | BI; S; P; F; T | M; S; W | 3 | - |
| Coswosk 2019 | Obs. | Qual | FGD; IDI; observation | Brazil | urban | 41 | 39 adolescent schoolgirls and schoolboys (13 – 17 years), 2 members of school staff | BI; H; S; | S | 5 | 2 |
| Coulter 2018 | Obs. | Quant | survey | Kenya | rural | 153 | 153 women in water resource user associations (17 – 87 years) | T; DM; L; N | W | 5 | 2 |
| Crow 2002 | Obs. | Qual | NR | Bangladesh | rural | NR | NR (17 – 87 years) | BI; H; S; P; F; T; N; R | S; W | NA | - |
| Crow 2010 | Obs. | Qual | observation; participatory methods; KII; IDI | Kenya | urban | 14 | 14 IDIs with water traders, landlords, water users, board members plus KIIs and case studies | BI; H; F; T; N | W | 5 | 2 |
| Crow 2012 | Obs. | Mixed* | Spring census; FGD; IDI; survey | Kenya | rural | 156 | 156 adult men and women in households and institutions | T; K; DM; CA; N; R | W | 4 | - |
| Czerniewska 2019 | Obs. | Qual | IDI | Tanzania | peri-urban, rural | 55 | 55 adult heads of household/representatives (20 – 69 years) | F; DM; N | S; W | 5 | 1 |
| Daniel 2019 | Obs. | Quant | Survey | Nepal | rural | 512 | 512 adult women | F; K | W | 3 | - |
| Das 2014 | Int. | Mixed | FGD; IDI; survey | India | urban | 386 | 386 adult men and women | S; P; CC; F; T; K; DM; L; CA; FOM; N; R | W | 5 | 3 |

| | | | | | | | | | | | |
|---|---|---|---|---|---|---|---|---|---|---|---|
| Das 2015 | Obs. | Quant | Survey; biological sample collection | India | urban | 486 | 486 adult women seeking hospital care (18 – 45 years) | H | S; W | 5 | - |
| Datta 2020 | Obs. | Qual | IDI; KII; participatory methods; digital capacity building workshops | India | urban | NR; at least 17 | 17 adult women, key informants | BI; H; S; N | S; W | 5 | 2 |
| de Moraes 2013 | Int. | Qual | Case study; IDI; participant observation | Brazil | rural | 36 | 36 adult rural women and NGO workers | CC; F; T; SC; K; CA; FOM; N; R | W | 5 | 3 |
| Delgado 2007 | Obs. | Qual | Ethnography | Peru | rural | 2 | 2 women who are separated but not divorced from their husbands | F; SC; L; N; R | W | NA | - |
| Devasia 1998a | Obs. | Qual | Participatory methods | India | rural | NR | adult men and women | BI; H; CC; F; T; SC; K; DM; L; CA; N; R; FLP | W | 4 | - |
| Devasia 1998b | Int. | Qual | participatory methods | India | rural | 955 | 955 adult women | CC; F; K; DM; L; CA; R | W | 4 | - |
| Devries 2015 | Int. | Qual | Group interviews | Guatemala | urban, rural | 956 | 956 respondents from savings groups | CC; K; DM; L; CA | W | - | 3 |
| Dreibelbis 2013 | Int. | Quant* | Survey, structured questionnaires, observations | Kenya | rural | 11,956 | head teachers from 175 schools, 3,815 heads of households, 7,966 schoolgirls and schoolboys (boy mean age = 10.7; girl mean age = 10.3) | T; K; N | S; W | 4 | - |
| Dudeja 2016 | Obs. | Quant | Survey | India | urban | 211 | 211 adolescent schoolgirls (mean age = 14.9) | BI | S | 4 | 2 |
| Dwipayanti 2019 | Obs. | Qual | IDI; KII | Indonesia | rural | 34 | 34 adult heads of household/representatives | T; DM; N | S | 5 | 2 |
| El Katsha 1989 | Obs. | Mixed | Survey; observation | Egypt | rural | 312 | 312 adult women | BI; H; CC; T; K; | M; S; W | 3 | - |

| | | | | | | | | | | | |
|---|---|---|---|---|---|---|---|---|---|---|---|
| | | | | | | | | DM; CA; N; R | | | |
| Elledge 2020 | Obs. | Mixed | Survey; IDI | US, UK, Austria, India, South Africa | NR | 17 | 17 BMGF grantee teams working in sanitation technologies | S; P; T; DM; L | S | 3 | - |
| Ellis 2016 | Obs. | Qual | FGD; observation | Philippines | urban, rural | 79 | 79 adolescent schoolgirls (11 – 18 years) | BI; H; S; P; T; K; FOM; N; R | M; S | 5 | - |
| Enabor 1998 | Int. | Quant | Survey; water quality testing | Nigeria | urban | 324 | 324 adult women | BI; F; K; CA; | W | 3 | - |
| Ennis-Millan 2001 | Obs. | Qual | IDI; observation | Mexico | peri-urban | NR; at least 41 | 41 adult men and women, civil and religious authorities (20 – 99 years) | BI; H; CC; F; CA; N; R | W | 5 | - |
| Ennis-McMillan 2005 | Obs. | Qual | IDI; observation; informal conversations; community assemblies | Mexico | peri-urban | 21 | 21 adult men and women | F; T; DM; L; CA; N; R | W | 5 | - |
| Faisal 2005 | Obs. | Mixed | FGD; KII; survey | Bangladesh | rural | NR | adult women, mostly married women | BI; H; S; P; F; T; SC; K; FOM; N; R | S; W | 2 | 3 |
| Fiasorgbor 2013 | Obs. | Qual | FGD; KII; observation | Ghana | urban | 368 | 368 adolescent schoolchildren (aged 12+), adult men and women | F; T | W | 5 | - |
| Fonjong 2014 | Obs. | Mixed | FGD; IDI; observation; survey | Cameroon | NR | 167 | 167 adult men and women | H; S; F; T; N; R | W | 4 | 2 |
| Gabrielsson 2013 | Obs. | Mixed | FGD; IDI; survey; participatory methods | Kenya | rural | NR; at least 181 | 181 elderly farmers (aged 60+), government stakeholders, widows, and farming groups | H; F; T; SC; CA; N | S; W | 4 | 4 |
| Gate 2001 | Obs. | Qual | Case study | India | rural | NR | adult women | CC; F; K; DM; CA; N; R | S; W | 5 | 2 |
| Ge 2011 | Obs. | Qual | IDI; KII; observation; FGD; survey | China | rural | 38 | 38 adult migrant returnees | SC; K; DM; L; CA; N; R | W | 5 | 2 |

| | | | | | | | | | | | |
|---|---|---|---|---|---|---|---|---|---|---|---|
| Geere 2018 | Obs. | Quant | Survey | South Africa, Ghana, and Vietnam | peri-urban; rural; urban | 3,365 | 3,365 minors, adult men and women (mean age = 29.7) | H | W | 3 | 1 |
| Girod 2017 | Obs. | Qual | FGD; KII; observation; anonymous question sessions | Kenya | urban | NR; at least 12 | 6 KIIs and 6 FGDs with adolescent schoolgirls, head teachers | BI; S; P; K | M; S; W | 5 | 1 |
| Gonsalves 2015 | Obs. | Quant | Secondary data analysis | South Africa | urban | 12,000 | ~12,000 adult women (15 – 49 years) | S; T | S | NA | - |
| Graham 2016 | Obs. | Quant | Secondary data analysis | Burkina Faso, Burundi, Cameroon, CAR, Cote d'Ivoire, Ethiopia, Gambia, Ghana, Guinea, Lesotho, Liberia, Madagascar, Malawai, Mali, Mauritania, Mozambique, Namibia, Niger, Nigeria, Sao Tome & Principe, Sierra Leone, Somalia, Swaziland, Zimbabwe | country-wide | 267,607 | analysis covered 267,607 women and children; no targeted population for DHS or MICS | T; N | W | 4 | - |
| Grant 2019 | Obs. | Qual | IDI | Cambodia | rural | 27 | 27 government stakeholders, female water entrepreneurs | CC; F; T; SC; K; DM; CA; FOM; N; R; FLP | W | 5 | 4 |
| Hall 2018 | Obs. | Qual | KII | Australia | rural | 17 | 17 representatives from government, research, NGOs, and water utility | BI | S; W | 5 | - |
| Halvorson 2004 | | Qual | FGD; IDI; observation | Pakistan | rural | 65 | 65 mothers and female caregivers of children under 5 | BI; H; F; T; K; DM; FOM; N | S; W | 5 | - |

| | | | | | | | | | | | |
|---|---|---|---|---|---|---|---|---|---|---|---|
| Hanrahan 2019 | Int. | Qual | FGD; observation | Canada | rural | 5 | 5 women | BI; H; F; SC; N; R | W | 5 | 2 |
| Harris 2017 | Obs. | Quant | Survey | Ghana and South Africa | urban | 493 | 493 household representatives | H; F; T; K; DM; N | W | 4 | - |
| Hassan 2004 | Obs. | Quant | Survey | Chile, Egypt, India, Philippines | urban | 3,995 | 3,995 adult heads of household/representatives (15 – 49 years) | S | S | 4 | 4 |
| Hennegan 2018 | Obs. | Quant | Survey | Nigeria | urban | 1,994 | 1,994 women and girls who had a menstrual period within the 3 months prior to the survey (aged 15+) | S; P | M; S | 5 | - |
| Hirai 2016 | Obs. | Quant | Secondary data analysis | Kenya | country-wide | 4,556 | 4,556 married and cohabitating adult women (15 – 49 years) | F; K; DM | S | 4 | - |
| Hirve 2015 | Obs. | Mixed | FGD; KII; participatory methods; survey, free-listing | India | rural | NR; at least 357 | 308 survey participants, 49 free-listing participants, and FGDS with adult and elderly women, adolescent girls (aged 13+) | BI; H; S; P; K; CA; R | S; W | 5 | 2 |
| Hoque 1994 | Int. | Mixed | Observation; survey | Bangladesh | rural | NR | adult women | K; L; CA; R | S | 4* | - |
| Hulland 2015 | Obs. | Qual | IDI; participatory methods | India | urban, rural | 60 | 60 adolescent girls, newly married, pregnant, and established adult women (14 – 24 years) | BI; H; S; P; FOM; N | M; S; W | 5 | - |
| Ilahi 2000 | Obs. | Quant | Secondary data analysis | Pakistan | rural | 2,505 | 2,505 adult women (mean age = 33.8 years) | F; T | W | NA | - |
| Indarti 2019 | Obs. | Qual | IDI | Indonesia | rural | 18 | 18 women engaged in the WASH sector (business owners, mobilizers, public sector employees) (23 – 61 years) | CC; F; T; SC; K; DM; L; FOM; R | S; W | 5 | 4 |
| Irianti 2019 | Obs. | Quant | Secondary data analysis | Indonesia | country-wide | 155,392 | 155,392 household representatives | T; N | S | 4 | 1 |
| Jadhav 2016 | Obs. | Quant | Secondary data analysis | India | national | 75,619 | 75,619 adolescent girls, adult women (15 – 49 years) | S | W | 4 | - |

| | | | | | | | | | | | |
|---|---|---|---|---|---|---|---|---|---|---|---|
| James 2002 | Int. | Qual | FGD; IDI; PRA | India | rural | 112 | 112 FGD participants and interviews with female members and leaders of micro-enterprise groups and controls, husbands of members; adult men | F; T; DM; N | W | 5 | 4 |
| Janmohamed 2016 | Obs. | Quant | Survey; biological sample collection; anthropometrics | Cambodia | rural | 544 | 544 pregnant women in their first trimester (mean age = 26.5years) | H | S | 5 | - |
| Jewitt 2014 | Obs. | Qual | FGD; IDI; participatory methods | Kenya | urban | 70 | 70 adolescent schoolgirls, teachers and headteachers, sex workers, former students (aged 13+) | BI; P | M; S; W | 5 | 2 |
| Jha 2012 | Int. | Qual | Observation | India | rural | NR | NR | BI; S; F; K; DM; L; FOM; N; R | S; W | 4 | 3 |
| Joshi 2012 | Obs. | Qual | NR | Bangladesh | urban | NR | adult men and women | BI; S; P; F; T; K; N; R | S; W | NA | - |
| Joshi 2018 | Obs. | Qual | FGD; IDI; observation | India | urban | 36 | 36 women waste-pickers | BI; H; S; P; F; T; SC; CA; R | M; S; W | 5 | 2 |
| Joshy 2019 | Obs. | Mixed | FGD; IDI; participatory methods; survey | India | rural | 117 | 117 adolescent schoolgirls, adult women, adult men, key stakeholders (13 – 53 years) | BI; N | S; W | 3 | 2 |
| Karim 2012 | Obs. | Mixed | IDI; KII; survey | Bangladesh | rural | 217 | 217 married women, households, key informants | S; T; N; R | W | 4 | - |
| Karin 2020 | Obs. | Mixed | FGD; KII; survey; case studies | Bangladesh | refugee camps | 102 | 102 adult women, adolescent girls | BI | S; W | 4 | 2 |
| Kernecker 2017 | Obs. | Qual | IDI; observation; participatory methods | Mexico | rural | 19 | 19 adult women | SC; K; CA; R | W | 5 | - |
| Khanna 2016 | Obs. | Qual | FGD; KII | India | rural | NR; at least 84 | 84 adult women, key informants | BI; H; S; P; F; T; SC; K; DM; FOM; N; R | S | 5 | - |

| | | | | | | | | | | | |
|---|---|---|---|---|---|---|---|---|---|---|---|
| Kher 2015 | Obs. | Mixed | IDI; survey | India | urban | 300 | 300 adult women | BI; H; S; P; F; T; SC; N; R | S; W | 4 | 1 |
| Klugman 2019 | Obs. | Quant | Secondary data analysis | 144 countries | country-level | n/a | n/a | H | S; W | 4 | 2 |
| Kodjebacheva 2019 | Obs. | Qual | FGD; IDI | United States of America | urban | 100 | 100 female college students (18 – 45 years) | K | W | 5 | - |
| Kookana 2016 | Obs. | Quant | Survey; school attendance records | India | rural | 72 | 72 adolescent schoolgirls and schoolboys (13 – 14 years) | BI; N | S; W | 3 | 3 |
| Krumdieck 2016 | Obs. | Quant | survey | Kenya | NR | 323 | 323 pregnant women of mixed HIV status | BI; H; S; F; T; N; R | W | 3 | - |
| Krusz 2019 | Obs. | Qual | Participatory methods | Australia | urban, rural, remote | 17 | 17 adult women including indigenous women and non-indigenous co-researchers and practitioners | BI | S | 5 | - |
| Kulkarni 2017 | Obs. | Qual | FGD; IDI | India | urban | 112 | 112 adult women | BI; H; S; P; F; T; CA; FOM; N; R | S; W | 5 | 3 |
| Kwiringira 2014 | Obs. | Qual | FGD; KII, participatory methods | Uganda | urban | NR; at least 27 | 15 KIIs and 12 FGDs with adult men and women, landlords, local leaders | BI; H; S; P; CC; F; SC; FOM; N | S | 5 | - |
| Leahy 2017 | Int. | Mixed | IDI; pocket voting | Vietnam | rural | 187 | 187 adult men and women from different ethnic groups, people with disabilities (aged 18+) | CC; K; DM; R | W | 3 | 4 |
| Lebel 2015 | Obs. | Mixed | IDI; survey | Thailand | peri-urban, rural | 1,055 | 1,055 adult men and women | CC; T; SC; K; DM; L; FOM; N; R | W | 4 | - |
| Leder 2017 | Int. | Qual | FGD; IDI; life histories; observations; household survey | Nepal | rural | 100 | ~100 adult men and women | H; CC; F; T; SC; K; DM; L; N; R | W | 5 | 4 |

| | | | | | | | | | | | |
|---|---|---|---|---|---|---|---|---|---|---|---|
| Lee 2017 | Obs. | Quant | Survey | India | rural | 19,124 | 19,124 adult men and women | F; K; DM | S | 4 | 2 |
| Leventhal 2016 | Int. | Quant | Questionnaires | India | rural | 3,363 | 3,363 schoolgirls (mean age = 12.97) | K | M; W | 3* | - |
| MacRae 2019 | Obs. | Qual | FGD; IDI; free listing | India | rural | 114 | 114 adult women (aged 18+) | BI; H; P; T; SC; FOM; N | M; S; W | 5 | - |
| Makoni 2004 | Obs. | Mixed | Survey; participatory methods | Zimbabwe | rural | NR | adult men and women | H; T; DM; L; N | S; W | 2 | 2 |
| Malhotra 2016 | Obs. | Quant | Survey | India | rural | 1,800 | 1,800 adolescent girls (10 – 19 years) | BI; P; CC | S; W | 3 | 2 |
| Mandara 2013 | Obs. | Mixed | FGD; KII; observation; survey; case studies | Tanzania | rural | NR; at least 221 | 221 household interviews, and KIIs and FGDs with adult men and women | BI; F; T; K; DM; N; FLP | W | 3 | - |
| Mandara 2017 | Obs. | Mixed | FGD; KII; observation; survey; case studies | Tanzania | rural | NR; at least 221 | 221 surveys, and IDIs and FGDs with adult men and women | | W | 2 | 2 |
| Mannan 2018 | Obs. | Qual | IDI | Bangaldesh | urban | 5 | 5 representatives from sanitation stakeholders | K; DM; N; FLP | S | 2 | 3 |
| Mason 2012 | Obs. | Qual | IDI | Philippines | urban | 22 | 22 adult men and women (22 – 60 years) | BI; H; S; F; T; SC; DM; N | W | 5 | - |
| Mbatha 2011 | Obs. | Qual | IDI | Swaziland | rural | 16 | 16 adolescent schoolgirls (14 – 17 years), school administrators | BI; H; P; T; N | S; W | 5 | - |
| McCammon 2020 | Obs. | Qual | IDI | India | urban | 70 | 70 young women, mostly unmarried (15 – 24 years) | BI; H; P; SC; K; R | S; W | 5 | - |
| McLean 2019 | Int. | Qual | IDI | Rwanda | urban, peri-urban, rural | 15 | 15 married couples (18 - 49 years) | N | W | 5 | 1 |
| McMahon 2011 | Obs. | Qual | FGD; IDI; field notes | Kenya | rural | 57 | 57 adolescent schoolgirls (12 – 16 years), teachers | BI; H; P; T; K | M; S; W | 5 | 2 |
| Mehretu 1992 | Obs. | Quant | Survey | Zimbabwe | rural | 331 | 331 participants | H; T; N | W | 3 | - |

| | | | | | | | | | | | |
|---|---|---|---|---|---|---|---|---|---|---|---|
| Mehta 2015 | Obs. | Mixed | FGD; survey | India | rural | NR; at least 180 | 180 surveys and FGDs with adult men and women | H; T; K; DM; FOM; N | W | 5 | - |
| Mmbengwa 2014 | Obs. | Mixed | IDI; observation; survey | South Africa | Country-wide | 409 | 409 representatives of water boards | L | W | 2 | 4 |
| Mohankumar 2015 | Obs. | Qual | FGD; IDI | India | | 5 | 5 female Dalit manual scavengers | S; F; T; DM; N; R | S | 5 | - |
| Mommen 2017 | Obs. | Quant | survey | Vanuatu | rural | 365 | 365 members of water committees | F; L | W | 4 | 2 |
| Mushavi 2020 | Obs. | Mixed | IDI; survey | Uganda | rural | 1,642 | 1,642 adolescents, women caregivers for children under 5 (aged 16+) | BI; H; S; F; T; SC; CA; N; R | M; W | 5 | - |
| Nagpal 2019 | Obs. | Mixed | IDI; survey; observation | India | urban | 350 | 350 adult household heads (aged 18+) | BI; H; P; F; T; N | S; W | 2 | - |
| Naiga 2017 | Obs. | Mixed | FGD; IDI; KII; participatory methods; survey | Uganda | rural | NR; at least 871 | 871 adult household heads; participant numbers NR for FGDs and participatory workshops | CC; F; K; DM; L; CA; FOM; N; FLP | W | 4 | - |
| Nallari 2015 | Obs. | Qual | FGD; IDI | India | urban | 30 | 30 adolescent girls | BI; H; S; P; F; T; SC; K; FOM; N; R | M; S; W | 5 | 2 |
| Nalugya 2020 | Int. | Mixed | FGD; IDI; observation; survey | Uganda | NR | NR; at least 859 | 859 students aged 13-21, parents, teachers; participant numbers NR for FGDs | BI; S; P; N; R | M; S; W | 5 | - |
| Nankinga 2019 | Obs. | Quant | Secondary data analysis | Uganda | nation-wide | 10,956 | 10,956 women of reproductive age (15 – 49 years) | H | W | 4 | 2 |
| Narain 2014 | Int. | Qual | IDI; KII; observation; PRA | India | rural | 60 | 60 adult men and women | H; T; SC; N; R | W | 5 | - |
| Nerkar 2013 | Int. | Qual | FGD | India | rural | 45 | 45 adult men and women (aged 18+) | H; CC; F; T; SC; K; CA; R | S; W | 5 | 3 |

| | | | | | | | | | | | |
|---|---|---|---|---|---|---|---|---|---|---|---|
| Ngila 2014 | Obs. | Quant | Survey | Kenya | NR | 468 | 468 adolescent schoolgirls, teachers | BI; H; P; K | S | 4 | - |
| Norling 2 | Obs. | Qual | IDI | Sweden | urban | 21 | 21 adolescent schoolgirls and schoolboys (16 – 18 years) | BI; H; S; P; CC; SC; R | S; W | 5 | - |
| Oluyemo 2012 | Obs. | Mixed | IDI; survey | Nigeria | towns | 558 | 558 adult men and women | BI; H; S; P; F; T; K; DM | S; W | 2 | 4 |
| O'Reilly 2006 | Int. | Qual | IDI; long-term participant observation | India | rural | NR | adult women, project staff | P; CC; F; SC; K; DM; L; FOM; N; R | W | 5 | 4 |
| O'Reilly 2010 | Int. | Qual | IDI; field notes | India | rural | NR | adult women | BI; H; S; P; F; SC; DM; FOM; N; R | M; S | NA | 4 |
| O'Reilly 2014 | Obs. | Mixed | IDI; observation | India | rural | 607 | 607 adult men and women (aged 18+) | BI; H; P; F; K; L; N; R; FLP | S; W | 3 | - |
| Padmaja 2020 | Int. | Mixed | FGD; survey; GILIT qualitative tool | India | rural | 700 | 700 adult men and women | T; DM; L; N; FLP | W | 3 | 2 |
| Panda 2012 | Int. | Qual | FGD; KII | India | rural | NR | adult men and women | CC; F; K; L; CA; N; R; FLP | W | 5 | 2 |
| Pardeshi 2008 | Int. | Mixed | FGD; KII; participatory methods; survey | India | rural | NR; at least 416 | 416 members of total sanitation cell; NR survey | L; CA | S | 3 | - |
| Pardeshi 2009 | Int. | Mixed | FGD; IDI; participatory methods; survey | India | rural | NR | adult women | BI; S; F; K; DM; L; CA; N; R | M; S | 2 | 2 |

| | | | | | | | | | | | |
|---|---|---|---|---|---|---|---|---|---|---|---|
| Pommells 2018 | Obs. | Qual | FGD; KII | Rwanda, Tanzania, Uganda, and Kenya | urban | 36 | 36 participants in FGDs with social and health professionals working in women's or maternal health, university students in a graduate public health leadership program | BI; H; S; F; T; SC; FOM; N; R | W | 5 | 1 |
| Porter 2011 | Obs. | Mixed | FGD; IDI; KII; participatory methods | Ghana | rural | NR | in-school and out-of school children, parents, settlement leaders, teachers, health workers, transport operators | T | W | 2 | - |
| Prasad 2015 | Obs. | Qual | FGD | India | rural | 120 | ~120 adult men and women | F; K; N; R; FLP | S | 5 | 2 |
| Prasad 2018 | Obs. | Qual | Ethnography | India | urban | NR | NR | F; K; R | S | NA | - |
| Privott 2019 | Obs. | Qual | IDI; panel discussion | United States of America | | 6 | 6 indigenous women "water protectors" | CA | W | 5 | 1 |
| Prokopy 2004 | Obs. | Mixed | FGD; IDI; observation; PRA; survey | India | rural | NR | systems operators and NGO representatives | CC; F; T; K; DM; L; N; R | W | 3 | 3 |
| Rajagopal 2017 | Int. | Mixed | FGD; IDI; survey | India | urban | 270 | 270 in-school and out-of school adolescent girls (10 – 20 years) | BI; H | S; W | 3 | 2 |
| Rajaraman 2013 | Obs. | Qual | IDI | India | urban | 48 | 48 adult women construction workers, domestic workers, street vendors, and garment factory workers | BI; H; P; F; T; N | M; S | 5 | - |
| Rajbangshi 2020 | Obs. | Qual | FGD | India | rural | 134 | 134 female plantation workers (18 – 55 years) | BI; H | S; W | 5 | - |
| Ramanaik 2018 | Int. | Qual | IDI | India | urban | 36 | 36 adolescent schoolgirls (13 – 17 years) | T; N | W | 5 | 3 |
| Rautanen 2008 | Int. | Mixed | IDI; observation; survey | Nepal | rural | 201 | 9 interviews, 95 workshop participants, and 97 surveys with adult women who have been trained in technical skills (16 – 55 years) | CC; F; T; SC; K; DM; CA; FOM; N; R | S; W | 3 | 2 |

| | | | | | | | | | | | |
|---|---|---|---|---|---|---|---|---|---|---|---|
| Reddy 2008 | Obs. | Mixed | FGD; participatory methods; survey | India | urban | NR; at least 64 | 64 minors, adult women and men; participant numbers NR for FGDs and PRA | BI; H; F; T; SC; DM; N | S; W | 3 | 3 |
| Reddy 2011 | Obs. | Mixed | FGD; observation; participatory methods; survey | India | urban | 32 | minors, adult women and men from 32 households | BI; H; S; P; F; T; SC; N; R | M; S; W | 4 | - |
| Reddy 2019 | Obs. | Qual | IDI | India | urban | 21 | 21 adult women | BI; H; S; P; F | S; W | 5 | - |
| Remigios 2011 | Obs. | Qual | FGD; IDI; observation | Zimbabwe | urban | NR | minors, adult women and men, city officials, town engineer | BI; H; S; CC; T; K; DM; N; R; FLP | S; W | 5 | 2 |
| Rheinländer 2019 | Obs. | Qual | FGD; IDI; observation | Ghana | peri-urban | 37 | 37 adolescent schoolgirls (15 – 23 years), school staff | BI; H; S; P; SC; K; FOM; N; R | M; S | 5 | 1 |
| Routray 2015 | Obs. | Qual | FGD | India | rural | 95 | 95 married and unmarried adult men and women (aged 16+) | BI; S; P; T; SC; FOM; R | M; S; W | 5 | 3 |
| Routray 2017a | Int. | Qual | FGD; IDI; observation | India | rural | NR; at least 80 | 80 adult men and women, 16-18-year-old boys and girls, NGO field staff; number of participants in FGDs NR | H; P; K; DM; CA; N; R | S | 5 | - |
| Routray 2017b | Obs. | Mixed | FGD; IDI; survey | India | rural | 560 | 560 young married men and women, male and female heads of household (23 – 86 years) | CC; F; K; DM; FOM; N; R | S; W | 4 | - |
| Sahoo 2015 | Obs. | Qual | IDI | India | urban, rural | 56 | 56 adolescent girls, newly married women, pregnant women, and established women (aged 16+) | BI; H; S; P; T; SC; FOM; N; R | S; W | 5 | - |
| Sam 2020 | Int. | Qual | FGD; IDI; observations; case study approach | Ghana | rural | 40 | 40 adult male and female residents of study communities, traditional leaders, WATSAN committee members, town committee members (18 – 76 years) | L | W | 5 | 3 |

| | | | | | | | | | | | |
|---|---|---|---|---|---|---|---|---|---|---|---|
| Schmitt 2017 | Obs. | Qual | FGD; KII; participatory methods | Myanmar and Lebanon | urban, peri-urban, camps | 205 | 205 internally displaced and refugee adolescent girls and adult women (14 – 49 years) | BI; H; S; P; T; K; N | M; S; W | 5 | - |
| Scorgie 2016 | Obs. | Qual | IDI; participatory methods; photovoice | South Africa | Unclear. Near Durban, South Africa, possibly peri-urban | 21 | 21 adult women (18 – 35 years) | BI; S; P; | M; S | 5 | 2 |
| Scott 2017a | Int. | Qual | FGD; IDI; observation | India | rural | NR; at least 92 | 74 adult men and women; 18 FGDs | T; SC; DM; L; N; R | S; W | 5 | - |
| Scott 2017b | Int. | Qual | FGD; IDI; observation | India | rural | NR; at least 92 | 74 adult men and women; 18 FGDs | F; SC; DM; L; FOM; N; R; FLP | S; W | 5 | 3 |
| Senior 2014[234] | Int. | Mixed | FGD; survey | Australia | urban | 240 | 240 schoolboys and schoolgirls (6 – 12 years) | BI; S; P | S | 3 | - |
| Shahid 2015 | Obs. | Qual | FGD; interaction | India | rural | NR | adult women | BI; H; CC; F; T; K; DM; FOM; N; R; FLP | S | 4 | 1 |
| Shiras 2018 | Int. | Qual | FGD; IDI; observation | Mozambique | urban | 143 | 143 adult men and women (18+) | BI; H; S; P; F; SC; CA; R | S | 5 | 1 |
| Siddiqui 2003 | Obs. | Quant | Survey | India | urban | 280 | 280 adult men and women | H | W | 3 | - |
| Sijbesma 2009 | Mixed | Mixed | FGD; PRA; review of account books; case studies | India | rural | NR | adult female members of enterprise groups and their husbands | F; T; DM; R | S; W | 3 | 2 |
| Sijbesma 2012 | Int. | Qual | Participatory methods | Bangladesh, India, Sri Lanka | urban, peri-urban | NR | adolescent boys and girls, women | F; T; K; DM; L; CA; | W | 5 | 3 |

| | | | | | | | | | | | |
|---|---|---|---|---|---|---|---|---|---|---|---|
| | | | | | | | | FOM; N; R | | | |
| Silva 2020 | Obs. | Qual | IDI; observation | Brazil | rural | 41 | 41 adult men and women | BI; helath; S; P; T; SC; N; R | M; S; W | 5 | 3 |
| Simiyu 2017 | Obs. | Qual | IDI; inspection of sanitation location | Kenya | urban | 39 | 39 tenants, landlords, caretakers | DM; R | S | 3 | - |
| Singh 2005 | Obs. | Qual | IDI; KII; observation; case studies | India | rural | 495 | 495 adult men and women | BI; F; SC; FOM; N; R; FLP | W | 5 | 1 |
| Singh 2006a | Obs. | Qual | FGD; IDI; KII; observation | India | rural | 420 | 420 men and women from various castes and ethnic groups | L; FOM; N | W | 5 | - |
| Singh 2006b | Obs. | Qual | FGD; KII; observation; case studies | India | rural | NR | men and women from various castes and ethnic groups | T; DM; L; FOM; N; R | W | 5 | 2 |
| Singh 2018 | Obs. | Mixed | FGD; observation; survey | India | rural` | NR; at least 300 | 300 adult men and women; number of FGD participants NR | F; K; DM; L; CA | W | - | 2 |
| Singh 2019 | Obs. | Qual | IDI; observation | India | urban | 60 | 60 adult women (20 – 50 years) | BI; H; S; F; SC; FOM; N; R | S | 4 | 2 |
| Smith 2004 | Int. | Mixed | FGD; survey | South Africa | urban | NR; at least 300 | 300 heads of household; number of FGD participants NR | F; K; L | S | 5 | 2 |
| Sommer 2010 | Obs. | Qual | IDI; observation; participatory methods; review of curriculum and attendance records | Tanzania | urban, rural | NR; at least 16 | 16 adolescent schoolgirls, drop-out adolescent girls (16 – 19 years), important adults in girls' lives | BI | M; S; W | 5 | 2 |
| Sommer 2015a | Obs. | Qual | KII; observation; participatory methods | Tanzania, Ghana, Cambodia, and Ethiopia | urban, rural | 450 | ~450 in-school and out-of-school adolescents, teachers, parents, health workers (16 – 19 years) | BI; P | S | 5 | 2 |
| Sommer 2015b | Obs. | Quant | Secondary data analysis | 32 sub-Saharan African countries | country-wide | NR | adult women | H | S; W | 5 | - |

| | | | | | | | | | | | |
|---|---|---|---|---|---|---|---|---|---|---|---|
| Sommer 2018 | Obs. | Qual | FGD; participatory methods | Democratic Republic of the Congo, Ethiopia | Rural; refugee camps | 330 | 330 caregivers and adolescent girls in conflict-affected villages and refugee camps (10 – 19 years) | S; P; FOM; N | S; W | 5 | 3 |
| Sorenson 2011 | Obs. | Quant | Secondary data analysis | 44 countries | country-wide | NR | heads of household | T; N | W | 4 | - |
| Stevenson 2012 | Obs. | Mixed | Freelisting, ranking, FGD; household surveys; water insecurity questionnaire | Ethiopia | rural | 497 | 497 female heads of household or wives of heads of household (mean age = 39) | BI; H; S; T; K; DM; N; R | W | 4 | 1 |
| Stevenson 2016 | Int. | Quant | Survey | Ethiopia | rural | 123 | 123 female heads of household or wives of heads of household | H; R | W | 4 | - |
| Sultana 2009a | Obs. | Qual | FGD; IDI; observation; case studies | Bangladesh | rural | 232 | 232 adult men and women; number of FGD participants NR | SC; K; DM; L; N; R | W | - | 2 |
| Sultana 2009b | Obs. | Qual | FGD; IDI; observation; case studies | Bangladesh | rural | 232 | 232 adult men and women; number of FGD participants NR | H; S; P; SC; DM; FOM; N; R | W | - | 2 |
| Tam 2012 | Obs. | Qual | IDI; face-to-face contact | Timor-Leste | rural | 17 | 17 adult men and women | K; DM; CA; N | S; W | 5 | 4 |
| Tegegne 2014 | Obs. | Mixed | FGD; IDI; survey | Ethiopia | rural | 608 | 595 surveys, 9 IDIs, and 4 FGDs with adolescent schoolgirls, female school drop-outs, female teachers | P; K | M; S; W | 5 | 2 |
| Thai 2019 | Int. | Qual | FGD; IDI | Vietnam | rural | NR | adult women, government officials | BI; H; S; CC; F; T; K; DM; L; N; R | W | 4 | 2 |
| Thompson 2017 | Obs. | Qual | Participatory methods | Cameroon | urban, rural | 130 | 130 adult men and women | BI; H; S; P; CC; T; N; R | S; W | 5 | 1 |
| Thuita 2017 | Obs. | Qual | FGD | Kenya | rural | 20 | 20 adolescent schoolgirls, adult women (17 – 80 years) | H; S; P; CC; F; K; DM; L; N; R | M; S | 5 | - |
| Torri 2010 | Int. | Mixed | FGD; IDI; observation | India | rural | 44 | 44 adult women (20 – 52 years) | S; CC; T; K; DM; | W | 5 | 3 |

| | | | | | | | | | | | |
|---|---|---|---|---|---|---|---|---|---|---|---|
| | | | | | | | | CA; FOM; N; R; FLP | | | |
| Tortajada 2003 | Obs. | Qual | IDI | Morocoo | urban | NR | Senior-level staff members of major water-related institutions | F; T; SC; K; DM; L; N; R | W | 5 | - |
| Trinies 2011 | Int. | Qual | IDI | India | rural | 15 | 15 adult women | BI; H; F; SC; K; DM; R | W | 5 | - |
| Trinies 2015 | Obs. | Qual | IDI; KII | Mali | urban, rural | NR; at least 40 | 40 adolescent schoolgirls (12 – 17 years), school employees | BI; P | M; S; W | 5 | - |
| Tsai 2016 | Obs. | Quant | Survey | Uganda | rural | 531 | 531 women with young children, men who live with participating women | BI; H | W | 5 | - |
| Van Houweling 2012 | Obs. | Mixed | FGD; IDI; survey | Senegal | rural | NR; at least 2,012 | 1,860 surveys, 137 IDIs, and 15 FGDs with adult men and women | BI; H; F; SC; N | W | 3 | 2 |
| Van Houweling 2015 | Int. | Qual | FGD; IDI; KII; observation; participatory methods | Mozambique | rural | NR; at least 170 | 155 interviews and 15 FGDs with adult men and women | H; S; F; T; SC; N; R | W | 4 | 3 |
| Van Houweling 2016 | Int. | Qual | FGD; IDI; observation | Mozambique | rural | NR; at least 75 | 60 IDIs and 15 FGDs with adult men and women | H; S; P; CC; F; T; SC; L; FOM; N; R; FLP | W | 5 | 3 |
| Varickanickal, 2019 | Obs. | Qual | IDI; photovoice | Kenya | rural | 13 | 13 women of various ages, with and without children (aged 17+) | H; S; CC; F; T; SC; DM; CA; R | S; W | 5 | 2 |
| Varua 2018 | Obs. | Qual | FGD; IDI | India | rural | 95 | 95 adult women (19 – 67 years) | F; T; DM; N | W | 2 | 4 |
| von Medeazza 2015 | Int. | Qual | Case studies | India | rural | NR | NR | BI; H; P; F; T; DM; N | S; W | 5 | 3 |

| | | | | | | | | | | | |
|---|---|---|---|---|---|---|---|---|---|---|---|
| Wall 2018 | Obs. | Qual | FGD; IDI; KII; case histories | Ethiopia | rural | 322 | 322 religious leaders, principals, teachers, nurses, pre-menarchal girls, menstruating adolescent girls, adolescent boys, married women of reproductive age, married men, post-menopausal women | BI; P; N | M; S; W | 5 | - |
| Waterkeyn 2005 | Int. | Mixed | IDI; survey; observation | Zimbabwe | rural | 928 | 908 surveys and 20 interviews with minors, adult club members of intervention groups, control individuals who are not members | CC; DM; L; FOM; R | S | 3 | - |
| Whale 2018 | Obs. | Qual | IDI; participatory methods | United Kingdom | NR | 20 | 20 schoolgirls and schoolboys with continence problems (11 – 19 years) | BI | S | 5 | - |
| White 2016 | Obs. | Qual | IDI; observation; participatory methods; photovoice | Malawi | urban, peri-urban, rural | 51 | 51 people with disabilities, caregivers of people with disabilities (8 – 87 years) | H; N | W | 5 | 2 |
| Willetts 2010 | Int. | Qual | Participatory methods | Fiji and Vanuatu | rural | NR | adult men and women | T; R | W | 5 | 2 |
| Winter 2015 | Obs. | Quant | Secondary data analysis | Kenya | country-wide | 6,191 | 6,191 adolescent girls, women (15 – 49 years) | S | S | 4 | - |
| Winter 2018 | Obs. | Qual | IDI | Kenya | urban | 55 | 55 adult women (18 – 72 years) | BI; H; S; P; F; FOM; N | S | 5 | 1 |
| Winter 2019a | Obs. | Mixed | IDI; survey | Kenya | urban | NR | adult women | BI; S; P | S | 3 | - |
| Winter 2019b | Obs. | Qual | IDI | Kenya | urban | 55 | 55 adult women (18 – 72 years) | S; P; CC; F; DM; L; CA; | S | 5 | - |
| Winter 2019c | Obs. | Mixed | IDI; survey; sanitation walks | Kenya | rural | 205 | 205 adult women (aged 18+) | S; F | S | 4 | - |
| Winter 2019d | Obs. | Quant | Survey | Kenya | urban | 552 | 552 adult women (aged 18+) | H; K | S; W | 5 | - |

| | | | | | | | | | | | |
|---|---|---|---|---|---|---|---|---|---|---|---|
| Winter 2019e | Obs. | Quant | Survey | Kenya | urban | 552 | 552 adult women (aged 18+) | H | S; W | 4 | - |
| Winter 2019f | Obs. | Quant | Survey | Kenya | urban | 55 | 55 adult women (aged 18+) | BI; H; S; P; F; T; K; N; R | S | 2 | - |
| Winter 2020 | Obs. | Quant | Survey | Kenya | urban | 361 | 361 adult women living with a partner, married, in a relationship, recently separated, divorced, widowed, or in a casual relationship in the year before the study (aged 18+) | H | W | 4 | 2 |
| Wood 2012 | Int. | Qual | IDI | Malawi | rural | 55 | 55 program participants, friends and relatives, husbands of program participants, health workers, WaterGuard vendors | BI; H; F; SC; K; R | W | 5 | - |
| Wutich 2008 | Obs. | Mixed | IDI; KII; survey; diary of water acquisition and use | Bolivia | urban | 72 | 72 heads of household | BI; H; CC; F; T | S; W | 4 | - |
| Wutich 2009 | Obs. | Mixed | IDI; survey; field notes; participant observation | Bolivia | urban | 48 | 48 heads of household | BI; H; F; T; N; R | W | 4 | - |
| Wutich 2012 | Obs. | Mixed | IDI; observation | Bolivia | urban | 73 | 73 heads of household | CC; F; T; SC; K; DM; L; CA; N; R | W | 3 | - |
| Yerian 2014 | Obs. | Qual | KII: FGD: semi-structured observation | Kenya | rural | NR; at least 106 | 10 KIIs and 96-160 FGD participants including representatives from water management or conflict resolution groups adult men and women (FGDs) (aged 18+) | BI; S; F; T; SC; DM; L; CA; N; R | W | 5 | 3 |
| You 2020 | Int. | Mixed | FGD; IDI; survey; system performance data | Uganda | town of 15,000 with rural and more urban areas | NR; at least 789 | 789 adolescent schoolgirls (11 – 19 years); number of participants in FGDs NR | BI; H; S; F; K | S | 1 | 1 |
| Yuerlita 2017[6] | Obs. | Mixed | FGD; KII; survey; field reconnaissance surveys | Indonesia | rural | NR; at least 60 | 60 adult men and women; number of participants in FGDs and KIIs NR | P; CC; F; T; SC; K; | W | 4 | - |

| | | | | | | | | | | | |
|---|---|---|---|---|---|---|---|---|---|---|---|
| | | | | | | | | DM; CA; N; R | | | |
| Zolnikov 2016 | Int. | Qual | IDI | Kenya | unspecified. "semi-arid" is the only description given | 52 | 52 heads of household, primary water collectors, children (9 – 78 years) | BI; H; T; SC; K; N; R | W | 5 | 3 |

**Supplemental Table 4: Quality Appraisal of all Included Articles Using the Mixed Methods Appraisal Tool**

**Criteria from the Mixed Methods Appraisal Tool[1]**

KEY
Individual criteria scores can be either 0 (did not meet criteria) or 1 (met criteria); cells that are shaded in gray indicate that a criterion was not applicable to the study type

Qualitative, quantitative, mixed methods, and final scores ranged from 1-5, except in the case of RCTs, which had a maximum score of 4

† Indicates that the MMAT was deemed inappropriate for quality appraisal of the article.
* Indicates that an article was cross-checked by a second appraiser.

| Study | Final Score | Qualitative Score | Quantitative Score | Mixed Methods Score | 1.1 | 1.2 | 1.3 | 1.4 | 1.5 | 2.1 | 2.2 | 2.3 | 2.4 | 3.1 | 3.2 | 3.3 | 3.4 | 3.5 | 4.1 | 4.2 | 4.3 | 4.4 | 4.5 | 5.1 | 5.2 | 5.3 | 5.4 | 5.5 |
|---|---|---|---|---|---|---|---|---|---|---|---|---|---|---|---|---|---|---|---|---|---|---|---|---|---|---|---|---|
| Abrahams 2006 | 5 | 5 | | | 1 | 1 | 1 | 1 | 1 | | | | | | | | | | | | | | | | | | | |
| Abu 2019 | 5 | 5 | | | 1 | 1 | 1 | 1 | 1 | | | | | | | | | | | | | | | | | | | |
| Acey 2010* | 3 | 5 | 3 | 4 | 1 | 1 | 1 | 1 | 1 | | | | | | | | | | 1 | 0 | 1 | 0 | 1 | 0 | 1 | 1 | 1 | 1 |
| Adriaenssens 2018 | 5 | 5 | 5 | 5 | 1 | 1 | 1 | 1 | 1 | | | | | | | | | | 1 | 1 | 1 | 1 | 1 | 1 | 1 | 1 | 1 | 1 |
| Agesa 2019 | 4 | | 4 | | | | | | | | | | | | | | | | 1 | 1 | 1 | 0 | 1 | | | | | |
| Aguilar 2005† | | | | | | | | | | | | | | | | | | | | | | | | | | | | |
| Aihara 2015* | 4 | | 4 | | | | | | | | | | | 1 | 1 | 1 | 0 | 1 | | | | | | | | | | |
| Aihara 2016 | 5 | | 5 | | | | | | | | | | | | | | | | 1 | 1 | 1 | 1 | 1 | | | | | |
| Akolgo 2019 | 3 | 3 | 4 | 3 | 1 | 1 | 1 | 0 | 0 | | | | | | | | | | 1 | 0 | 1 | 1 | 1 | 1 | 1 | 0 | 0 | 1 |
| Aladuwaka 2010 | 5 | 5 | | | 1 | 1 | 1 | 1 | 1 | | | | | | | | | | | | | | | | | | | |
| Alam 2017 | 4 | | 4 | | | | | | | | | | | 1 | 0 | 1 | 1 | 1 | | | | | | | | | | |
| Ali 2013 | 5 | 5 | | | 1 | 1 | 1 | 1 | 1 | | | | | | | | | | | | | | | | | | | |
| Aluko 2019 | 4 | | 4 | | | | | | | | | | | | | | | | 1 | 1 | 1 | 0 | 1 | | | | | |
| Andajani-Sutjahjo 2015 | 5 | 5 | | | 1 | 1 | 1 | 1 | 1 | | | | | | | | | | | | | | | | | | | |
| Anderson 2013 | 5 | 5 | | | 1 | 1 | 1 | 1 | 1 | | | | | | | | | | | | | | | | | | | |

| | | | | | | | | | | | | | | | | | | | | | | | | | | | | |
|---|---|---|---|---|---|---|---|---|---|---|---|---|---|---|---|---|---|---|---|---|---|---|---|---|---|---|---|---|
| Anyarayor 2019 | 3 | 3 | 4 | 3 | 1 | 0 | 1 | 0 | 1 | | | | | | | | | | 1 | 1 | 1 | 0 | 1 | 1 | 0 | 0 | 1 | 1 |
| Arku 2010a | 5 | 5 | | | 1 | 1 | 1 | 1 | 1 | | | | | | | | | | | | | | | | | | | |
| Arku 2010b* | 5 | 5 | | | 1 | 1 | 1 | 1 | 1 | | | | | | | | | | | | | | | | | | | |
| Asaba 2013 | 4 | 5 | 4 | 5 | 1 | 1 | 1 | 1 | 1 | | | | | 1 | 1 | 1 | 0 | 1 | | | | | | 1 | 1 | 1 | 1 | 1 |
| Assad 2009 | 5 | 5 | | | 1 | 1 | 1 | 1 | 1 | | | | | | | | | | | | | | | | | | | |
| Azeez 2019 | 5 | 5 | | | 1 | 1 | 1 | 1 | 1 | | | | | | | | | | | | | | | | | | | |
| Baker 2017 | 5 | | 5 | | | | | | | | | | | 1 | 1 | 1 | 1 | 1 | | | | | | | | | | |
| Baluchova 2017 | 2 | 2 | | | 1 | 0 | 1 | 0 | 0 | | | | | | | | | | | | | | | | | | | |
| Bangdiwala 2004 | 4 | | 4 | | | | | | | | | | | | | | | | 1 | 1 | 1 | 0 | 1 | | | | | |
| Bapat 2003† | | | | | | | | | | | | | | | | | | | | | | | | | | | | |
| Barchi 2019 | 4 | | 4 | | | | | | | | | | | | | | | | 1 | 1 | 1 | 0 | 1 | | | | | |
| Bastidas 2005 | 2 | 5 | 2 | 5 | 1 | 1 | 1 | 1 | 1 | | | | | | | | | | 0 | 0 | 1 | 0 | 1 | 1 | 1 | 1 | 1 | 1 |
| Bastola 2015 | 2 | 2 | 3 | 3 | 1 | 1 | 0 | 0 | 0 | | | | | | | | | | 1 | 0 | 1 | 0 | 1 | 1 | 0 | 1 | 0 | 1 |
| BeBe 2015 | 4 | | 4 | | | | | | | | | | | | | | | | 1 | 1 | 1 | 0 | 1 | | | | | |
| Belur 2017 | 4 | 5 | 4 | 5 | 1 | 1 | 1 | 1 | 1 | | | | | | | | | | 1 | 1 | 1 | 0 | 1 | 1 | 1 | 1 | 1 | 1 |
| Bhandari 2009 | 2 | 5 | 2 | 5 | 1 | 1 | 1 | 1 | 1 | | | | | | | | | | 0 | 0 | 1 | 0 | 1 | 1 | 1 | 1 | 1 | 1 |
| Bhatt 2019 | 5 | 5 | | | 1 | 1 | 1 | 1 | 1 | | | | | | | | | | | | | | | | | | | |
| Bisung 2014 | 5 | | 5 | | | | | | | | | | | | | | | | 1 | 1 | 1 | 1 | 1 | | | | | |
| Bisung 2015a | 5 | 5 | | | 1 | 1 | 1 | 1 | 1 | | | | | | | | | | | | | | | | | | | |
| Bisung 2015b | 5 | 5 | | | 1 | 1 | 1 | 1 | 1 | | | | | | | | | | | | | | | | | | | |
| Bisung 2016 | 5 | 5 | | | 1 | 1 | 1 | 1 | 1 | | | | | | | | | | | | | | | | | | | |
| Bisung 2018 | 5 | | 5 | | | | | | | | | | | 1 | 1 | 1 | 1 | 1 | | | | | | | | | | |
| Bisung 2019 | 5 | 5 | | | 1 | 1 | 1 | 1 | 1 | | | | | | | | | | | | | | | | | | | |
| Boateng 2013a | 4 | 4 | 4 | 4 | 1 | 1 | 1 | 1 | 0 | | | | | | | | | | 1 | 1 | 1 | 0 | 1 | 1 | 0 | 1 | 1 | 1 |
| Boateng 2013b | 4 | 5 | 4 | 5 | 1 | 1 | 1 | 1 | 1 | | | | | | | | | | 1 | 1 | 1 | 0 | 1 | 1 | 1 | 1 | 1 | 1 |
| Boateng 2018 | 5 | 5 | 5 | 5 | 1 | 1 | 1 | 1 | 1 | | | | | | | | | | 1 | 1 | 1 | 1 | 1 | 1 | 1 | 1 | 1 | 1 |
| Boosey 2014 | 5 | 5 | 5 | 5 | 1 | 1 | 1 | 1 | 1 | | | | | | | | | | 1 | 1 | 1 | 1 | 1 | 1 | 1 | 1 | 1 | 1 |
| Bora 2016 | 2 | 2 | 5 | 5 | 1 | 1 | 0 | 0 | 0 | | | | | | | | | | 1 | 1 | 1 | 1 | 1 | 1 | 1 | 1 | 1 | 1 |
| Brewis 2018 | 4 | | 4 | | | | | | | | | | | 1 | 1 | 0 | 1 | 1 | | | | | | | | | | |
| Buor 2003 | 4 | | 4 | | | | | | | | | | | 1 | 1 | 1 | 0 | 1 | | | | | | | | | | |
| Bustamente 2015 | | | | | | | | | | | | | | | | | | | | | | | | | | | | |
| Cairns 2017 | 3 | 5 | 3 | 4 | 1 | 1 | 1 | 1 | 1 | | | | | | | | | | 1 | 1 | 1 | 0 | 0 | 1 | 1 | 0 | 1 | 1 |

| | | | | | | | | | | | | | | | | | | | | | | | | | | | | |
|---|---|---|---|---|---|---|---|---|---|---|---|---|---|---|---|---|---|---|---|---|---|---|---|---|---|---|---|---|
| Camenga 2019 | 5 | 5 | | | 1 | 1 | 1 | 1 | 1 | | | | | | | | | | | | | | | | | | | |
| Carmi 2019 | 3 | 5 | 3 | 5 | 1 | 1 | 1 | 1 | 1 | | | | | | | | | | 1 | 0 | 1 | 0 | 1 | 1 | 1 | 1 | 1 | 1 |
| Carolini 2012 | 2 | 2 | | | 1 | 1 | 0 | 0 | 0 | | | | | | | | | | | | | | | | | | | |
| Caruso 2017a | 3 | 3 | 5 | 5 | 1 | 1 | 1 | 0 | 0 | | | | | 1 | 1 | 1 | 1 | 1 | | | | | | 1 | 1 | 1 | 1 | 1 |
| Caruso 2017b | 5 | 5 | | | 1 | 1 | 1 | 1 | 1 | | | | | | | | | | | | | | | | | | | |
| Caruso 2018 | 5 | | 5 | | | | | | | | | | | 1 | 1 | 1 | 1 | 1 | | | | | | | | | | |
| Cheng 2012 | 5 | | 5 | | | | | | | | | | | | | | | | 1 | 1 | 1 | 1 | 1 | | | | | |
| Chew 2019 | 5 | 5 | | | 1 | 1 | 1 | 1 | 1 | | | | | | | | | | | | | | | | | | | |
| Chipeta 2019 | 4 | 4 | | | 1 | 0 | 1 | 1 | 1 | | | | | | | | | | | | | | | | | | | |
| Clement 2018 | 5 | 5 | | | 1 | 1 | 1 | 1 | 1 | | | | | | | | | | | | | | | | | | | |
| Coles 2009 | | | | | | | | | | | | | | | | | | | | | | | | | | | | |
| Collins 2018 | 5 | 5 | | | 1 | 1 | 1 | 1 | 1 | | | | | | | | | | | | | | | | | | | |
| Connolly 2013 | 5 | 5 | | | 1 | 1 | 1 | 1 | 1 | | | | | | | | | | | | | | | | | | | |
| Cooper-Vince 2017 | 5 | | 5 | | | | | | | | | | | 1 | 1 | 1 | 1 | 1 | | | | | | | | | | |
| Cooper-Vince 2018 | 5 | | 5 | | | | | | | | | | | 1 | 1 | 1 | 1 | 1 | | | | | | | | | | |
| Corburn 2015 | 3 | 5 | 3 | 4 | 1 | 1 | 1 | 1 | 1 | | | | | | | | | | 1 | 0 | 1 | 0 | 1 | 0 | 1 | 1 | 1 | 1 |
| Corburn 2016 | 3 | 5 | 3 | 4 | 1 | 1 | 1 | 1 | 1 | | | | | | | | | | 1 | 0 | 1 | 0 | 1 | 0 | 1 | 1 | 1 | 1 |
| Coswosk 2019 | 5 | 5 | | | 1 | 1 | 1 | 1 | 1 | | | | | | | | | | | | | | | | | | | |
| Coulter 2018 | 5 | 5 | | | 1 | 1 | 1 | 1 | 1 | | | | | | | | | | | | | | | | | | | |
| Crow 2002 | | | | | | | | | | | | | | | | | | | | | | | | | | | | |
| Crow 2010 | 4 | 5 | 5 | 4 | 1 | 1 | 1 | 1 | 1 | | | | | | | | | | 1 | 1 | 1 | 1 | 1 | 0 | 1 | 1 | 1 | 1 |
| Crow 2012 | 5 | 5 | | | 1 | 1 | 1 | 1 | 1 | | | | | | | | | | | | | | | | | | | |
| Czerniewska 2019 | 5 | 5 | | | 1 | 1 | 1 | 1 | 1 | | | | | | | | | | | | | | | | | | | |
| Daniel 2019 | 3 | | 3 | | | | | | | | | | | | | | | | 1 | 0 | 1 | 0 | 1 | | | | | |
| Das 2014 | 5 | 5 | | | 1 | 1 | 1 | 1 | 1 | | | | | | | | | | | | | | | | | | | |
| Das 2015 | 5 | | 5 | | | | | | | | | | | 1 | 1 | 1 | 1 | 1 | | | | | | | | | | |
| Datta 2020 | 5 | 5 | | | 1 | 1 | 1 | 1 | 1 | | | | | | | | | | | | | | | | | | | |
| de Moraes 2013 | 5 | 5 | | | 1 | 1 | 1 | 1 | 1 | | | | | | | | | | | | | | | | | | | |
| Delgado 2007 | | | | | | | | | | | | | | | | | | | | | | | | | | | | |
| Devasia 1998a | 4 | 4 | | | 1 | 1 | 1 | 0 | 1 | | | | | | | | | | | | | | | | | | | |

| | | | | | | | | | | | | | | | | | | | | | | | | | | | | |
|---|---|---|---|---|---|---|---|---|---|---|---|---|---|---|---|---|---|---|---|---|---|---|---|---|---|---|---|---|
| Devasia 1998b | 4 | 4 | | | 1 | 1 | 1 | 0 | 1 | | | | | | | | | | | | | | | | | | | |
| DeVries 2015 | 5 | 5 | | | 1 | 1 | 1 | 1 | 1 | | | | | | | | | | | | | | | | | | | |
| Dreibelbis 2013 | 4 | 5 | 4 | 4 | 1 | 1 | 1 | 1 | 1 | | | | | | | | | | 1 | 1 | 1 | 0 | 1 | 0 | 1 | 1 | 1 | 1 |
| Dudeja 2016 | 4 | | 4 | | | | | | | | | | | 1 | 1 | 1 | 0 | 1 | | | | | | | | | | |
| Dwipayanti 2019 | 5 | 5 | | | 1 | 1 | 1 | 1 | 1 | | | | | | | | | | | | | | | | | | | |
| El Katsha 1989 | 3 | 5 | 3 | 3 | 1 | 1 | 1 | 1 | 1 | | | | | | | | | | 1 | 0 | 1 | 0 | 1 | 1 | 1 | 0 | 0 | 1 |
| Elledge 2020 | 3 | 5 | 4 | 3 | 1 | 1 | 1 | 1 | 1 | | | | | | | | | | 1 | 1 | 0 | 1 | 1 | 0 | 1 | 1 | 0 | 1 |
| Ellis 2016 | 5 | 5 | | | 1 | 1 | 1 | 1 | 1 | | | | | | | | | | | | | | | | | | | |
| Enabor 2010 | 3 | | 3 | | | | | | | | | | | | | | | | 1 | 0 | 1 | 0 | 1 | | | | | |
| Ennis-McMillan 2001 | 5 | 5 | | | 1 | 1 | 1 | 1 | 1 | | | | | | | | | | | | | | | | | | | |
| Ennis-McMillan 2005 | 5 | 5 | | | 1 | 1 | 1 | 1 | 1 | | | | | | | | | | | | | | | | | | | |
| Faisal 2005 | 2 | 2 | 4 | 2 | 1 | 1 | 0 | 0 | 0 | | | | | | | | | | 1 | 1 | 1 | 0 | 1 | 0 | 1 | 0 | 0 | 1 |
| Fiasorgbor 2013 | 5 | 5 | | | 1 | 1 | 1 | 1 | 1 | | | | | | | | | | | | | | | | | | | |
| Fonjong 2014 | 4 | 5 | 4 | 5 | 1 | 1 | 1 | 1 | 1 | | | | | | | | | | 1 | 1 | 1 | 0 | 1 | 1 | 1 | 1 | 1 | 1 |
| Gabrielsson 2013 | 4 | 5 | 4 | 5 | 1 | 1 | 1 | 1 | 1 | | | | | | | | | | 1 | 1 | 1 | 0 | 1 | 1 | 1 | 1 | 1 | 1 |
| Gate 2001 | 5 | 5 | | | 1 | 1 | 1 | 1 | 1 | | | | | | | | | | | | | | | | | | | |
| Ge 2011 | 5 | 5 | | | 1 | 1 | 1 | 1 | 1 | | | | | | | | | | | | | | | | | | | |
| Geere 2018 | 3 | | 3 | | | | | | | | | | | | | | | | 1 | 0 | 1 | 0 | 1 | | | | | |
| Girod 2017 | 5 | 5 | | | 1 | 1 | 1 | 1 | 1 | | | | | | | | | | | | | | | | | | | |
| Gonsalves 2015 | | | | | | | | | | | | | | | | | | | | | | | | | | | | |
| Graham 2016 | 4 | | 4 | | | | | | | | | | | | | | | | 1 | 1 | 1 | 0 | 1 | | | | | |
| Grant 2019 | 5 | 5 | | | 1 | 1 | 1 | 1 | 1 | | | | | | | | | | | | | | | | | | | |
| Hall 2018 | 4 | 4 | | | 1 | 0 | 1 | 1 | 1 | | | | | | | | | | | | | | | | | | | |
| Halverson 2004 | 5 | 5 | | | 1 | 1 | 1 | 1 | 1 | | | | | | | | | | | | | | | | | | | |
| Hanrahan 2018 | 5 | 5 | | | 1 | 1 | 1 | 1 | 1 | | | | | | | | | | | | | | | | | | | |
| Harris 2017 | 4 | | 4 | | | | | | | | | | | | | | | | 1 | 1 | 1 | 0 | 1 | | | | | |

| | | | | | | | | | | | | | | | | | | | | | | | | | | | | |
|---|---|---|---|---|---|---|---|---|---|---|---|---|---|---|---|---|---|---|---|---|---|---|---|---|---|---|---|---|
| Hassan 2010 | 4 | | 4 | | | | | | | | | | | 1 | 1 | 1 | 0 | 1 | | | | | | | | | | |
| Hennegan 2018 | 5 | | 5 | | | | | | | | | | | 1 | 1 | 1 | 1 | 1 | | | | | | | | | | |
| Hirai 2016 | 5 | | 5 | | | | | | | | | | | | | | | | 1 | 1 | 1 | 1 | 1 | | | | | |
| Hirve 2015 | 5 | 5 | 5 | 5 | 1 | 1 | 1 | 1 | 1 | | | | | | | | | | 1 | 1 | 1 | 1 | 1 | 1 | 1 | 1 | 1 | 1 |
| Hoque 1994 | 1 | 4 | 1 | 4 | 1 | 0 | 1 | 1 | 1 | | | | | | | | | | 0 | 0 | 0 | 0 | 1 | 1 | 1 | 0 | 1 | 1 |
| Hulland, 2015 | 5 | 5 | | | 1 | 1 | 1 | 1 | 1 | | | | | | | | | | | | | | | | | | | |
| Ilahi 2000 | | | | | | | | | | | | | | | | | | | | | | | | | | | | |
| Indarti 2019 | 5 | 5 | | | 1 | 1 | 1 | 1 | 1 | | | | | | | | | | | | | | | | | | | |
| Irianti, 2019 | 4 | | 4 | | | | | | | | | | | | | | | | 1 | 1 | 1 | 0 | 1 | | | | | |
| Jadhav, 2018 | 4 | | 4 | | | | | | | | | | | | | | | | 1 | 1 | 1 | 0 | 1 | | | | | |
| James 2002 | 5 | 5 | | | 1 | 1 | 1 | 1 | 1 | | | | | | | | | | | | | | | | | | | |
| Janmohamed 2016 | 5 | | 5 | | | | | | | | | | | | | | | | 1 | 1 | 1 | 1 | 1 | | | | | |
| Jewitt 2014 | 5 | 5 | | | 1 | 1 | 1 | 1 | 1 | | | | | | | | | | | | | | | | | | | |
| Jha 2012 | 4 | 4 | | | 1 | 1 | 0 | 1 | 1 | | | | | | | | | | | | | | | | | | | |
| Joshi 2017 | 5 | 5 | | | 1 | 1 | 1 | 1 | 1 | | | | | | | | | | | | | | | | | | | |
| Joshi, 2012 | | | | | | | | | | | | | | | | | | | | | | | | | | | | |
| Joshy, 2019 | 3 | 5 | 3 | 4 | 1 | 1 | 1 | 1 | 1 | | | | | | | | | | 1 | 1 | 0 | 0 | 1 | 0 | 1 | 1 | 1 | 1 |
| Karim, 2012 | 4 | 5 | 4 | 5 | 1 | 1 | 1 | 1 | 1 | | | | | | | | | | 1 | 1 | 1 | 0 | 1 | 1 | 1 | 1 | 1 | 1 |
| Karin 2020 | 3 | 5 | 3 | 5 | 1 | 1 | 1 | 1 | 1 | | | | | | | | | | 1 | 0 | 1 | 0 | 1 | 1 | 1 | 1 | 1 | 1 |
| Kernecker 2017 | 5 | 5 | | | 1 | 1 | 1 | 1 | 1 | | | | | | | | | | | | | | | | | | | |
| Khanna 2016 | 5 | 5 | | | 1 | 1 | 1 | 1 | 1 | | | | | | | | | | | | | | | | | | | |
| Kher, 2015 | 4 | 5 | 4 | 4 | 1 | 1 | 1 | 1 | 1 | | | | | | | | | | 1 | 1 | 1 | 0 | 1 | 0 | 1 | 1 | 1 | 1 |
| Klugman, 2019 | 4 | | 4 | | | | | | | | | | | | | | | | 1 | 1 | 1 | 0 | 1 | | | | | |
| Kodjebacheva 2018 | 5 | 5 | | | 1 | 1 | 1 | 1 | 1 | | | | | | | | | | | | | | | | | | | |
| Kookana, 2016 | 3 | | 3 | | | | | | | | | | | | | | | | 1 | 0 | 1 | 0 | 1 | | | | | |
| Krumdieck 2016 | 3 | | 3 | | | | | | | | | | | 0 | 1 | 1 | 0 | 1 | | | | | | | | | | |
| Krusz 2019 | 5 | 5 | | | 1 | 1 | 1 | 1 | 1 | | | | | | | | | | | | | | | | | | | |
| Kulkarni 2017 | 5 | 5 | | | 1 | 1 | 1 | 1 | 1 | | | | | | | | | | | | | | | | | | | |
| Kwiringira 2014 | 5 | 5 | | | 1 | 1 | 1 | 1 | 1 | | | | | | | | | | | | | | | | | | | |

| | | | | | | | | | | | | | | | | | | | | | | | | | | | | |
|---|---|---|---|---|---|---|---|---|---|---|---|---|---|---|---|---|---|---|---|---|---|---|---|---|---|---|---|---|
| Leahy 2017 | 4 | 5 | 4 | 4 | 1 | 1 | 1 | 1 | 1 | | | | | | | | | | 1 | 1 | 1 | 0 | 1 | 0 | 1 | 1 | 1 | 1 |
| Lebel 2015 | 4 | 5 | 4 | 4 | 1 | 1 | 1 | 1 | 1 | | | | | | | | | | 1 | 1 | 0 | 1 | 1 | 0 | 1 | 1 | 1 | 1 |
| Leder 2017 | 5 | 5 | | | 1 | 1 | 1 | 1 | 1 | | | | | | | | | | | | | | | | | | | |
| Lee 2017 | 4 | | 4 | | | | | | | | | | | | | | | | 1 | 1 | 1 | 0 | 1 | | | | | |
| Leventhal 2016 | 3 | | 3 | | | | | | | 1 | 1 | 1 | 0 | | | | | | | | | | | | | | | |
| MacRae 2019 | 5 | 5 | | | 1 | 1 | 1 | 1 | 1 | | | | | | | | | | | | | | | | | | | |
| Makoni 2004 | 2 | 2 | 3 | 2 | 1 | 1 | 0 | 0 | 0 | | | | | | | | | | 1 | 1 | 0 | 0 | 1 | 0 | 1 | 0 | 0 | 1 |
| Malhotra 2016 | 3 | | 3 | | | | | | | | | | | | | | | | 1 | 0 | 1 | 0 | 1 | | | | | |
| Mandara 2013 | 3 | 5 | 3 | 4 | 1 | 1 | 1 | 1 | 1 | | | | | | | | | | 1 | 0 | 1 | 0 | 1 | 0 | 1 | 1 | 1 | 1 |
| Mandara 2017 | 2 | 5 | 2 | 5 | 1 | 1 | 1 | 1 | 1 | | | | | | | | | | 1 | 0 | 0 | 0 | 1 | 1 | 1 | 1 | 1 | 1 |
| Mannan 2018 | 2 | 2 | | | 1 | 1 | 0 | 0 | 0 | | | | | | | | | | | | | | | | | | | |
| Mason 2012 | 5 | 5 | | | 1 | 1 | 1 | 1 | 1 | | | | | | | | | | | | | | | | | | | |
| Mbatha 2011 | 5 | 5 | | | 1 | 1 | 1 | 1 | 1 | | | | | | | | | | | | | | | | | | | |
| McCammon 2020 | 5 | 5 | | | 1 | 1 | 1 | 1 | 1 | | | | | | | | | | | | | | | | | | | |
| McLean 2019 | 5 | 5 | | | 1 | 1 | 1 | 1 | 1 | | | | | | | | | | | | | | | | | | | |
| McMahon 2011 | 5 | 5 | | | 1 | 1 | 1 | 1 | 1 | | | | | | | | | | | | | | | | | | | |
| Mehretu 1992 | 3 | | 3 | | | | | | | | | | | | | | | | 1 | 0 | 1 | 0 | 1 | | | | | |
| Mehta 2015 | 5 | 5 | | | 1 | 1 | 1 | 1 | 1 | | | | | | | | | | | | | | | | | | | |
| Mmbengwa 2014 | 2 | 2 | 3 | 2 | 1 | 1 | 0 | 0 | 0 | | | | | | | | | | 1 | 0 | 1 | 0 | 1 | 1 | 0 | 0 | 0 | 1 |
| Mohankumar 2017 | 5 | 5 | | | 1 | 1 | 1 | 1 | 1 | | | | | | | | | | | | | | | | | | | |
| Mommen 2017 | 4 | | 4 | | | | | | | | | | | | | | | | 1 | 1 | 1 | 0 | 1 | | | | | |
| Mushavi 2020 | 5 | 5 | 5 | 5 | 1 | 1 | 1 | 1 | 1 | | | | | | | | | | 1 | 1 | 1 | 1 | 1 | 1 | 1 | 1 | 1 | 1 |
| Nagpal 2019 | 2 | | 2 | | | | | | | | | | | | | | | | 1 | 0 | 0 | 0 | 1 | | | | | |
| Naiga, 2017 | 5 | 5 | 5 | 5 | 1 | 1 | 1 | 1 | 1 | | | | | | | | | | 1 | 1 | 1 | 1 | 1 | 1 | 1 | 1 | 1 | 1 |
| Nallari 2015 | 5 | 5 | | | 1 | 1 | 1 | 1 | 1 | | | | | | | | | | | | | | | | | | | |
| Nalugya 2020 | 5 | 5 | 5 | 5 | 1 | 1 | 1 | 1 | 1 | | | | | | | | | | 1 | 1 | 1 | 1 | 1 | 1 | 1 | 1 | 1 | 1 |
| Nankinga 2019 | 4 | | 4 | | | | | | | | | | | | 1 | 1 | 0 | 1 | 1 | | | | | | | | | |
| Narain 2014 | 5 | 5 | | | 1 | 1 | 1 | 1 | 1 | | | | | | | | | | | | | | | | | | | |

| | | | | | | | | | | | | | | | | | | | | | | | | | | | | |
|---|---|---|---|---|---|---|---|---|---|---|---|---|---|---|---|---|---|---|---|---|---|---|---|---|---|---|---|---|
| Nerkar 2013 | 5 | 5 | | | 1 | 1 | 1 | 1 | 1 | | | | | | | | | | | | | | | | | | | |
| Ngila 2014 | 4 | | 4 | | | | | | | | | | | 1 | 1 | 1 | 0 | 1 | | | | | | | | | | |
| Norling 2016 | 5 | 5 | | | 1 | 1 | 1 | 1 | 1 | | | | | | | | | | | | | | | | | | | |
| Oluyemo 2012 | 2 | 2 | 3 | 5 | 1 | 1 | 0 | 0 | 0 | | | | | | | | | | 1 | 1 | 0 | 0 | 1 | 1 | 1 | 1 | 1 | 1 |
| O'Reilly 2006 | 5 | 5 | | | 1 | 1 | 1 | 1 | 1 | | | | | | | | | | | | | | | | | | | |
| O'Reilly 2010 | | | | | | | | | | | | | | | | | | | | | | | | | | | | |
| O'Reilly 2014 | 3 | 5 | 3 | 3 | 1 | 1 | 1 | 1 | 1 | | | | | | | | | | 1 | 0 | 1 | 0 | 1 | 1 | 1 | 0 | 0 | 1 |
| Padmaja 2020 | 3 | 4 | 3 | 5 | 1 | 1 | 1 | 1 | 0 | | | | | | | | | | 1 | 1 | 0 | 0 | 1 | 1 | 1 | 1 | 1 | 1 |
| Panda 2012 | 5 | 5 | | | 1 | 1 | 1 | 1 | 1 | | | | | | | | | | | | | | | | | | | |
| Pardeshi 2008 | 3 | 5 | 3 | 3 | 1 | 1 | 1 | 1 | 1 | | | | | | | | | | 1 | 1 | 0 | 0 | 1 | 0 | 1 | 0 | 1 | 1 |
| Pardeshi 2009 | 2 | 5 | 3 | 2 | 1 | 1 | 1 | 1 | 1 | | | | | | | | | | 1 | 1 | 0 | 0 | 1 | 0 | 1 | 0 | 0 | 1 |
| Pommells 2018 | 5 | 5 | | | 1 | 1 | 1 | 1 | 1 | | | | | | | | | | | | | | | | | | | |
| Porter, 2011 | 2 | 5 | 2 | 5 | 1 | 1 | 1 | 1 | 1 | | | | | | | | | | 1 | 0 | 0 | 0 | 1 | 1 | 1 | 1 | 1 | 1 |
| Prasad 2015 | 5 | 5 | | | 1 | 1 | 1 | 1 | 1 | | | | | | | | | | | | | | | | | | | |
| Prasad 2018 | | | | | | | | | | | | | | | | | | | | | | | | | | | | |
| Privott 2019 | 5 | 5 | | | 1 | 1 | 1 | 1 | 1 | | | | | | | | | | | | | | | | | | | |
| Prokopy 2004 | 3 | 3 | 4 | 4 | 1 | 1 | 1 | 0 | 0 | | | | | | | | | | 1 | 1 | 1 | 0 | 1 | 0 | 1 | 1 | 1 | 1 |
| Rajagopal 2017 | 3 | 5 | 3 | 4 | 1 | 1 | 1 | 1 | 1 | | | | | | | | | | 1 | 1 | 0 | 0 | 1 | 0 | 1 | 1 | 1 | 1 |
| Rajaraman 2013 | 5 | 5 | | | 1 | 1 | 1 | 1 | 1 | | | | | | | | | | | | | | | | | | | |
| Rajbangshi 2020 | 5 | 5 | | | 1 | 1 | 1 | 1 | 1 | | | | | | | | | | | | | | | | | | | |
| Ramanaik 2018 | 5 | 5 | | | 1 | 1 | 1 | 1 | 1 | | | | | | | | | | | | | | | | | | | |
| Rautanen 2005 | 3 | 3 | 3 | 4 | 1 | 1 | 1 | 0 | 0 | | | | | | | | | | 0 | 1 | 1 | 0 | 1 | 0 | 1 | 1 | 1 | 1 |
| Reddy 2008 | 3 | 5 | 3 | 5 | 1 | 1 | 1 | 1 | 1 | | | | | | | | | | 1 | 1 | 1 | 0 | 0 | 1 | 1 | 1 | 1 | 1 |
| Reddy 2011 | 4 | 5 | 4 | 5 | 1 | 1 | 1 | 1 | 1 | | | | | | | | | | 1 | 1 | 1 | 0 | 1 | 1 | 1 | 1 | 1 | 1 |
| Reddy 2019 | 5 | 5 | | | 1 | 1 | 1 | 1 | 1 | | | | | | | | | | | | | | | | | | | |
| Remigios 2011 | 5 | 5 | | | 1 | 1 | 1 | 1 | 1 | | | | | | | | | | | | | | | | | | | |
| Rheinlander 2018 | 5 | 5 | | | 1 | 1 | 1 | 1 | 1 | | | | | | | | | | | | | | | | | | | |
| Routray 2015 | 5 | 5 | | | 1 | 1 | 1 | 1 | 1 | | | | | | | | | | | | | | | | | | | |
| Routray 2017a | 5 | 5 | | | 1 | 1 | 1 | 1 | 1 | | | | | | | | | | | | | | | | | | | |

| | | | | | | | | | | | | | | | | | | | | | | | | | | | | |
|---|---|---|---|---|---|---|---|---|---|---|---|---|---|---|---|---|---|---|---|---|---|---|---|---|---|---|---|---|
| Routray 2017b | 4 | 5 | 4 | 5 | 1 | 1 | 1 | 1 | 1 | | | | | | | | | | 1 | 1 | 1 | 0 | 1 | 1 | 1 | 1 | 1 | 1 |
| Sahoo 2015 | 5 | 5 | | | 1 | 1 | 1 | 1 | 1 | | | | | | | | | | | | | | | | | | | |
| Sam 2020 | 5 | 5 | | | 1 | 1 | 1 | 1 | 1 | | | | | | | | | | | | | | | | | | | |
| Schmitt 2017 | 5 | 5 | | | 1 | 1 | 1 | 1 | 1 | | | | | | | | | | | | | | | | | | | |
| Scorgie 2015 | 5 | 5 | | | 1 | 1 | 1 | 1 | 1 | | | | | | | | | | | | | | | | | | | |
| Scott 2017a | 5 | 5 | | | 1 | 1 | 1 | 1 | 1 | | | | | | | | | | | | | | | | | | | |
| Scott 2017b | 5 | 5 | | | 1 | 1 | 1 | 1 | 1 | | | | | | | | | | | | | | | | | | | |
| Senior 2014 | 3 | 3 | 4 | 4 | 1 | 1 | 0 | 0 | 1 | | | | | | | | | | 1 | 1 | 0 | 1 | 1 | 0 | 1 | 1 | 1 | 1 |
| Shahid 2015 | 4 | 4 | | | 1 | 1 | 1 | 0 | 1 | | | | | | | | | | | | | | | | | | | |
| Shiras 2018 | 5 | 5 | | | 1 | 1 | 1 | 1 | 1 | | | | | | | | | | | | | | | | | | | |
| Siddiqui 2003 | 3 | | 3 | | | | | | | | | | | | | | | | 1 | 0 | 1 | 0 | 1 | | | | | |
| Sijbesma 2009 | 3 | 3 | 3 | 4 | 1 | 1 | 0 | 0 | 1 | | | | | | | | | | 1 | 0 | 1 | 0 | 1 | 0 | 1 | 1 | 1 | 1 |
| Sijbesma 2012 | 5 | 5 | | | 1 | 1 | 1 | 1 | 1 | | | | | | | | | | | | | | | | | | | |
| Silva 2020 | 5 | 5 | | | 1 | 1 | 1 | 1 | 1 | | | | | | | | | | | | | | | | | | | |
| Simiyu 2017 | 3 | 3 | | | 1 | 1 | 1 | 0 | 0 | | | | | | | | | | | | | | | | | | | |
| Singh 2005 | 5 | 5 | | | 1 | 1 | 1 | 1 | 1 | | | | | | | | | | | | | | | | | | | |
| Singh 2006a | 5 | 5 | | | 1 | 1 | 1 | 1 | 1 | | | | | | | | | | | | | | | | | | | |
| Singh 2006b | 5 | 5 | | | 1 | 1 | 1 | 1 | 1 | | | | | | | | | | | | | | | | | | | |
| Singh 2018 | 2 | 3 | 2 | 2 | 1 | 1 | 1 | 0 | 0 | | | | | | | | | | 1 | 0 | 0 | 0 | 1 | 1 | 0 | 0 | 0 | 1 |
| Singh 2019 | 4 | 4 | | | 1 | 1 | 0 | 1 | 1 | | | | | | | | | | | | | | | | | | | |
| Smith 2004 | 5 | 5 | | | 1 | 1 | 1 | 1 | 1 | | | | | | | | | | | | | | | | | | | |
| Sommer 2010 | 5 | 5 | | | 1 | 1 | 1 | 1 | 1 | | | | | | | | | | | | | | | | | | | |
| Sommer 2015a | 5 | 5 | | | 1 | 1 | 1 | 1 | 1 | | | | | | | | | | | | | | | | | | | |
| Sommer 2015b | 4 | | 4 | | | | | | | | | | | | | | | | 1 | 1 | 1 | 0 | 1 | | | | | |
| Sommer 2018 | 5 | 5 | | | 1 | 1 | 1 | 1 | 1 | | | | | | | | | | | | | | | | | | | |
| Sorenson 2011 | 5 | | 5 | | | | | | | | | | | | | | | | 1 | 1 | 1 | 1 | 1 | | | | | |
| Stevenson 2012 | 4 | 5 | 4 | 5 | 1 | 1 | 1 | 1 | 1 | | | | | | | | | | 1 | 1 | 1 | 0 | 1 | 1 | 1 | 1 | 1 | 1 |
| Stevenson 2016 | 4 | | 4 | | | | | | | | | | | 1 | 1 | 1 | 0 | 1 | | | | | | | | | | |
| Sultana 2009b | 5 | 5 | | | 1 | 1 | 1 | 1 | 1 | | | | | | | | | | | | | | | | | | | |
| Sultana 2009a | 5 | 5 | | | 1 | 1 | 1 | 1 | 1 | | | | | | | | | | | | | | | | | | | |

| | | | | | | | | | | | | | | | | | | | | | | | | | | | | |
|---|---|---|---|---|---|---|---|---|---|---|---|---|---|---|---|---|---|---|---|---|---|---|---|---|---|---|---|---|
| Tam 2012 | 5 | 5 | | | 1 | 1 | 1 | 1 | 1 | | | | | | | | | | | | | | | | | | | |
| Tegegne 2014 | 5 | 5 | 5 | 5 | 1 | 1 | 1 | 1 | 1 | | | | | | | | | | 1 | 1 | 1 | 1 | 1 | 1 | 1 | 1 | 1 | 1 |
| Thai 2019 | 4 | 4 | | | 1 | 1 | 0 | 1 | 1 | | | | | | | | | | | | | | | | | | | |
| Thompson 2017 | 5 | 5 | | | 1 | 1 | 1 | 1 | 1 | | | | | | | | | | | | | | | | | | | |
| Thuita 2017 | 5 | 5 | | | 1 | 1 | 1 | 1 | 1 | | | | | | | | | | | | | | | | | | | |
| Torri 2010 | 5 | 5 | | | 1 | 1 | 1 | 1 | 1 | | | | | | | | | | | | | | | | | | | |
| Tortajada 2003 | 5 | 5 | | | 1 | 1 | 1 | 1 | 1 | | | | | | | | | | | | | | | | | | | |
| Trinies, 2011 | 5 | 5 | | | 1 | 1 | 1 | 1 | 1 | | | | | | | | | | | | | | | | | | | |
| Trinies 2015 | 5 | 5 | | | 1 | 1 | 1 | 1 | 1 | | | | | | | | | | | | | | | | | | | |
| Tsai 2016 | 5 | | 5 | | | | | | | | | | | | | | | | 1 | 1 | 1 | 1 | 1 | | | | | |
| van Houweling 2012 | 3 | 3 | 3 | 4 | 1 | 1 | 1 | 0 | 0 | | | | | | | | | | 1 | 0 | 1 | 0 | 1 | 0 | 1 | 1 | 1 | 1 |
| Van Houweling 2015 | 5 | 5 | | | 1 | 1 | 1 | 1 | 1 | | | | | | | | | | | | | | | | | | | |
| Van Houweling 2016 | 5 | 5 | | | 1 | 1 | 1 | 1 | 1 | | | | | | | | | | | | | | | | | | | |
| Varickanickal 2019 | 5 | 5 | | | 1 | 1 | 1 | 1 | 1 | | | | | | | | | | | | | | | | | | | |
| Varua, 2018 | 2 | 2 | 4 | 4 | 1 | 1 | 0 | 0 | 0 | | | | | | | | | | 1 | 0 | 1 | 1 | 1 | 0 | 1 | 1 | 1 | 1 |
| Von Medeazza 2015 | 5 | 5 | | | 1 | 1 | 1 | 1 | 1 | | | | | | | | | | | | | | | | | | | |
| Wall 2018 | 5 | 5 | | | 1 | 1 | 1 | 1 | 1 | | | | | | | | | | | | | | | | | | | |
| Waterkeyn 2005 | 3 | 3 | 3 | 5 | 1 | 1 | 1 | 0 | 0 | | | | | 0 | 1 | 1 | 0 | 1 | | | | | | 1 | 1 | 1 | 1 | 1 |
| Whale 2018 | 5 | 5 | | | 1 | 1 | 1 | 1 | 1 | | | | | | | | | | | | | | | | | | | |
| White, 2016 | 5 | 5 | | | 1 | 1 | 1 | 1 | 1 | | | | | | | | | | | | | | | | | | | |
| Willetts 2010 | 5 | 5 | | | 1 | 1 | 1 | 1 | 1 | | | | | | | | | | | | | | | | | | | |
| Winter 2015 | 5 | | 5 | | | | | | | | | | | | | | | | 1 | 1 | 1 | 1 | 1 | | | | | |
| Winter 2018 | 5 | 5 | | | 1 | 1 | 1 | 1 | 1 | | | | | | | | | | | | | | | | | | | |
| Winter 2019a | 3 | 5 | 3 | 4 | 1 | 1 | 1 | 1 | 1 | | | | | | | | | | 1 | 0 | 1 | 0 | 1 | 0 | 1 | 1 | 1 | 1 |
| Winter 2019b | 5 | 5 | | | 1 | 1 | 1 | 1 | 1 | | | | | | | | | | | | | | | | | | | |
| Winter 2019c | 4 | 5 | 4 | 5 | 1 | 1 | 1 | 1 | 1 | | | | | | | | | | 1 | 1 | 1 | 0 | 1 | 1 | 1 | 1 | 1 | 1 |
| Winter 2019d | 5 | | 5 | | | | | | | | | | | 1 | 1 | 1 | 1 | 1 | | | | | | | | | | |

| | | | | | | | | | | | | | | | | | | | | | | | | | | | | |
|---|---|---|---|---|---|---|---|---|---|---|---|---|---|---|---|---|---|---|---|---|---|---|---|---|---|---|---|---|
| Winter 2019e | 4 | | 4 | | | | | | | | | | | | | | | | 1 | 1 | 1 | 0 | 1 | | | | | |
| Winter 2019f | 3 | | 3 | | | | | | | | | | | | | | | | 1 | 0 | 1 | 0 | 1 | | | | | |
| Winter, 2020 | 4 | | 4 | | | | | | | | | | | | | | | | 1 | 1 | 1 | 0 | 1 | | | | | |
| Wood, S., , 2012 | 5 | 5 | | | 1 | 1 | 1 | 1 | 1 | | | | | | | | | | | | | | | | | | | |
| Wutich 2008 | 4 | 5 | 4 | 5 | 1 | 1 | 1 | 1 | 1 | | | | | | | | | | 1 | 1 | 1 | 0 | 1 | 1 | 1 | 1 | 1 | 1 |
| Wutich 2009 | 4 | 5 | 4 | 5 | 1 | 1 | 1 | 1 | 1 | | | | | | | | | | 1 | 1 | 1 | 0 | 1 | 1 | 1 | 1 | 1 | 1 |
| Wutich 2012 | 3 | 3 | | | 1 | 1 | 1 | 0 | 0 | | | | | | | | | | | | | | | | | | | |
| Yerian 2014 | 5 | 5 | | | 1 | 1 | 1 | 1 | 1 | | | | | | | | | | | | | | | | | | | |
| You 2020 | 1 | 5 | 1 | 5 | 1 | 1 | 1 | 1 | 1 | | | | | | | | | | 0 | 0 | 0 | 0 | 1 | 1 | 1 | 1 | 1 | 1 |
| Yuerlita 2017 | 3 | 3 | 4 | 3 | 1 | 1 | 1 | 0 | 0 | | | | | | | | | | 1 | 1 | 1 | 0 | 1 | 1 | 1 | 0 | 0 | 1 |
| Zolnikov 2016 | 5 | 5 | | | 1 | 1 | 1 | 1 | 1 | | | | | | | | | | | | | | | | | | | |

[1]Hong, Q.N., Pluye, P., et al. Mixed Methods Appraisal Tool (MMAT) Version 2018 User Guide. McGill Department of Family Medicine. 2018.

**Criteria from the MMAT:**

1.1 Is the qualitative approach appropriate to answer the research question?
1.2 Are the qualitative data collection methods adequate to address the research question?
1.3 Are the findings adequately derived from the data?
1.4 Is the interpretation of results sufficiently substantiated by data?
1.5 Is there coherence between qualitative data sources, collection, analysis and interpretation?
2.1 Is randomization appropriately performed?
2.2 Are the groups comparable at baseline?
2.3 Are there complete outcome data?
2.4 Are outcome assessors blinded to the intervention provided?
3.1 Are the participants representative of the target population?
3.2 Are measurements appropriate regarding both the outcome and intervention (or exposure)?
3.3 Are there complete outcome data?
3.4 Are the confounders accounted for in the design and analysis?
3.5 During the study period, is the intervention administered (or exposure occurred) as intended?
4.1 Is the sampling strategy relevant to address the research question?
4.2 Is the sample representative of the target population?
4.3 Are the measurements appropriate?
4.4 Is the risk of nonresponse bias low?
4.5 Is the statistical analysis appropriate to answer the research question?
5.1 Is there an adequate rationale for using a mixed methods design to address the research question?
5.2 Are the different components of the study effectively integrated to answer the research question?
5.3 Are the outputs of the integration of qualitative and quantitative components adequately interpreted?
5.4 Are divergences and inconsistencies between quantitative and qualitative results adequately addressed?
5.5 Do the different components of the study adhere to the quality criteria of each tradition of the methods involved?

**Supplemental Table 5: Articles that Define or Explore Definitions of Women's Empowerment**

| Author (year) | Definition of women's empowerment used in paper |
|---|---|
| **Aguilar 2005** | Defines empowerment as "the perspective of making the community the decision maker." |
| **Aladuwaka 2010** | Draws on Kabeer's (1994)[1] definition, including access to tangible and intangible resources, and focuses on the potential for a water project to challenge unequal power relations, meeting women's strategic needs for time as well as building women's sense of "power within" and building consciousness of the challenges they face and how to address them |
| **Bisung 2019** | Provides a review of multiple definitions in the literature but does not utilize one specific definition; the goal of the research activity is for local actors and stakeholders to generate their own definitions of empowerment |
| **Clement 2018** | Draws on Eyben's (2008)[2] definition as "a process whereby 'individuals and organized groups are able to imagine their world differently and to realize that vision by changing the relations of power that have been keeping them in poverty'" and expands Kabeer's (1994)[1] framework to focus on four dimensions of women's empowerment: critical consciousness, access to resources, agency, and achievements |
| **Gabrielsson 2013** | Draws on Kabeer's (1999)[3] definition of empowerment as a process, "whereby people gain the ability to make choices they were previously denied," focusing on resources, agency, and achievements |
| **Grant 2019** | Incorporates several empowerment frameworks, including Longwe's framework (1991)[4], which consists of five stages of empowerment: (1) Welfare; (2) Access; (3) Conscientisation; (4) Participation; and (5) Control; and the four types of power, explained by Rowlands (1995)[5] and elaborated on over time by other scholars. |
| **Indarti 2019** | Draws on Cornwall and Rivas (2015)[6] and Rao and Kelleher (2005)[7] to define empowerment as "a process of transforming power relations in favor of women's rights and social justice and the transformation of economic, social, and political structures;" a transformation which, per Rao and Kelleher, "requires equal access to and control over resources (human, physical, intellectual), control over ideology (beliefs, values, attitudes), and changes in the institutions and structures that support unequal power relations." |
| **James et al., 2002** | Draws on Self-Employed Women's Association's (SEWA) (1999)[8] gender approach of helping poor women to become economically independent, providing assistance "to organize themselves, change their self-concept, and take their own development in hand." SEWA's women's empowerment approach focuses on ensuring "that women have the opportunity to earn and to benefit from their earnings." strives to ensure that women have the opportunity to earn and to benefit from their earnings" (p. 208) |
| **Leahy 2017** | Draws on Kabeer's (1999)[9] model, which "conceptualises the provision of material resources to women as the starting point for a process whereby they are able to put their choices and aims into action, ultimately achieving empowerment" (p. 287). |
| **Leder 2017** | Draws on Kabeer's (1999)[3] definition of empowerment that focuses on resources, agency, and achievement. States "Women's empowerment is a holistic concept that has social and political, as well as economic, dimensions. It requires change to gendered structures of power, but also to structures of power relating to other aspects of identity including class, race, and caste" (p. 237). "In addition to support extending their access and control over |

| | |
|---|---|
| | resources, women need support extending their agency. Agency could be described as the ability to make strategic decisions about what one values" (p. 238). |
| **Mmbengwa 2014** | Provides a brief review of multiple definitions of empowerment from a variety of sources, but does not commit to any one specific definition. The study's aim is "to investigate the empowerment of women in the water boards of South Africa by examining their proportional representation in the boards. It is assumed that the representation of women in the structures of governance shows the readiness to provide women with the opportunity to help shape the direction of the services of the water sector in South Africa." |
| **Oluyemo 2012** | Defines women's empowerment as "the ability to build the capacity of women in various endeavor in order for them to make informed and decisions about their lives and that of society. It is moving beyond the status quo, moving beyond cultural beliefs and practices, economic, social, religious, and political restrictions that constraint women in the society [sic]" |
| **O'Reilly 2006** | Provides a review of how programs and projects have defined empowerment in past projects and linked it to modernity, and how village women have rejected or embraced individual components; does not utilize one particular definition of empowerment for the purpose of research |
| **O'Reilly 2010** | Draws on Rowlands' (1997)[10] definition: "Empowerment is a process through which self-confidence, agency, and dignity increase" including control of resources and opportunities (p. 49). |
| **Tam 2012** | Draws on Longwe's (1997)[11] framework, which includes five levels of empowerment, includingt: (1) Welfare; (2) Access; (3) Conscientisation; (4) Participation; and (5) Control |
| **Varua 2018** | Defines empowerment as the "creation of an environment for women where they can make decisions of their own for their personal benefits as well as for the society." Draws on the Food and Agriculture Organization's (FAO) Policy on Gender Equality[12] to identify domains, which include: (i) equal participation of women and men as decision-makers in rural institutions and in shaping laws, policies and programmes; (ii) equal access for women and men to productive resources, assets, decent employment opportunities, and income; (iii) equal access for women and men to goods and services for agricultural development and to markets"; (iv) reduction of women's work burden through the provision of improved technologies, services and infrastructure; and (v) an increase share of total agricultural aid committed to projects that target women and promote gender equality" (p. 97). |

[1]Kabeer, N. *Reversed realities: Gender hierarchies in development thought.* Verso.1994.
[2]Eyben, R., Kabeer, N., Cornwall A. Conceptualizing empowerment and the implications for pro poor growth: a paper for the DAC poverty network; Institute of Development Studies (IIDS) Brighton. 1994.
[3]Kabeer, N. Resources, agency, achievements: Reflections on the measurement of women's empowerment. *Development and Change* 1999; *30(3)* 435-464.
[4]Wallace, T., March, C. Changing Perceptions: Writings on Gender and Development; Oxfam, GB: Oxford, UK, 1991.
[5]Rowlands, J. Empowerment examined. *Development in Practice*. 1995; 5, 101-107.
[6]Cornwall, A., Rivas, A.-M. From 'Gender Equality' and 'Women's Empowerment' to Global Justice: Reclaiming a Transformative Agenda for Gender and Development. *Third World Quarterly* 2015; 36(2): 396-415.
[7]Rao, A., Kelleher, D. Is there Life after Gender Mainstreaming? *Gender and Development* 2005; 13(2): 57-69
[8]Self-Employed Women's Association (SEWA). *Learning the Process of Development* Banaskantha DWACRA Mahila SEWA Association (BDSMA), Ahmedabad. 1999
[9]Kabeer, N. *The Conditions and Consequences of Choice: Reflections on the Measurement of Women's Empowerment*. UNRISD Discussion Paper No. 108, United Nations Research Institute for Social Development; Geneva. 1999.
[10]Rowlands, J. *Questioning Empowerment: Working with women in Honduras*. Oxfam: Oxford. 1997
[11]CCGD (Collaborative Centre for Gender and Development) Gender Training of Trainer – An Introduction. CCGD 1997
[12] Full citation not provided in paper

**Supplemental Table 6: Articles that Engage Menstrual Hygiene Management by Domain and Sub-Domain**

| **Empowerment Domains & Sub-Domains** | **Articles that Engage the Domain/Sub-Domain (N=198)** | | |
|---|---|---|---|
| | **Water** | **Water *and* Sanitation** | **Sanitation** |
| **AGENCY (N = 2)** | (*n* = 1) MacRae 2019 | (*n* = 0) | (*n* = 1) Winter 2019b |
| **Decision-making (N = 1)** | (*n* = 0) | (*n* = 0) | (*n* = 1) Winter 2019b |
| **Leadership (N = 1)** | (*n* = 0) | (*n* = 0) | (*n* = 1) Winter 2019b |
| **Collective Action (N = 0)** | (*n* = 0) | (*n* = 0) | (*n* = 0) |
| **Freedom of Movement (N = 1)** | (*n* = 1) MacRae 2019 | (*n* = 0) | (*n* = 0) |
| **RESOURCES (N = 34)** | (*n* = 2) Leventhal 2016; Mushavi 2020 | (*n* = 16) Caruso 2017b; Connolly 2013; Girod 2017; Hulland 2015; Jewitt 2014; MacRae 2019; McMahon 2011; Nallari 2015; Nalugya 2020; Reddy 2011; Schmitt 2017; Silva 2020; Sommer 2010; Tegegne 2014; Trinies 2015; Wall 2018 | (*n* = 16) Alam 2017; Belur 2017; Bhandari 2009; Corburn 2015; Corburn 2016; Ellis 2016; Hennegan 2018; Joshi 2017; O'Reilly 2010; Pardeshi 2009; Rajaraman 2013; Rheinlander 2018; Routray 2015; Scorgie 2015; Thuita 2017; Winter 2019b |
| **Bodily Integrity (N = 22)** | (*n* = 2) Mushavi 2020; Wall 2018 | (*n* = 10) Connolly 2013; Jewitt 2014; MacRae 2019; McMahon 2011; Nalugya 2020; Reddy 2011; Schmitt 2017; Silva 2020; Sommer 2010; Trinies 2015 | (*n* = 10) Alam 2017; Belur 2017; Corburn 2015; Corburn 2016; Girod 2017; Ellis 2016; O'Reilly 2010; Pardeshi 2009; Rajaraman 2013; Rheinlander 2018 |
| *Health (N = 8)* | (*n* = 1) McMahon 2011 | (*n* = 4) Hulland 2015; MacRae 2019; Nallari 2015; Reddy 2011 | (*n* = 3) Ellis 2016; Joshi 2017; Rajaraman 2013 |
| *Privacy (N = 18)* | (*n* = 2) Bhandari 2009; Girod 2017 | (*n* = 4) Connolly 2013; MacRae 2019; Nallari 2015; Reddy 2011 | (*n* = 12) Corburn 2016; Hennegan 2018; Jewitt 2014; McMahon 2011; Nalugya 2020; Rheinlander 2018; Routray 2015; Schmitt 2017; Scorgie 2015; Tegegne 2014; Trinies 2015; Wall 2018 |
| *Safety and Security* | (*n* = 0) | (*n* = 0) | (*n* = 4) |

| | | | |
|---|---|---|---|
| *(N = 4)* | | | Girod 2017; Hennegan 2018; Nalugya 2020; Pardeshi 2009 |
| **Critical Consciousness (N = 1)** | (*n* = 0) | (*n* = 0) | (*n* = 1) Winter 2019b |
| **Assets (N = 11)** | (*n* = 3) Leventhal 2016; MacRae 2019; McMahon 2011 | (*n* = 2) Caruso 2017b; Tegegne 2014 | (*n* = 6) Ellis 2016; Girod 2017; Rajaraman 2013; Schmitt 2017; Thuita 2017; Winter 2019b |
| *Financial and Productive Assets (N = 2)* | (*n* = 0) | (*n* = 0) | (*n* = 2) Rajaraman 2013; Winter 2019b |
| *Knowledge & Skills (N = 7)* | (*n* = 2) Leventhal 2016; McMahon 2011 | (*n* = 1) Tegegne 2014 | (*n* = 4) Ellis 2016; Girod 2017; Schmitt 2017; Thuita 2017 |
| *Social Capital (N = 1)* | (*n* = 1) MacRae 2019 | (*n* = 0) | (*n* = 0) |
| *Time (N = 4)* | (*n* = 2) MacRae 2019; McMahon 2011 | (*n* = 1) Caruso 2017b | (*n* = 1) Schmitt 2017 |
| **INSTITUTIONAL STRUCTURES (N = 8)** | (*n* = 2) MacRae 2019; Wall 2018 | (*n* = 4) Belur 2017; Caruso 2017b; El Katsha 1989; Schmitt 2017 | (*n* = 2) Ellis 2016; Nalugya 2020 |
| **Formal Laws & Policies (N = 0)** | (*n* = 0) | (*n* = 0) | ( *n* = 0) |
| **Norms (N = 7)** | (*n* = 2) MacRae 2019; Wall 2018 | (*n* = 3) Caruso 2017b; El Katsha 1989; Schmitt 2017 | (*n* = 2) Ellis 2016; Nalugya 2020 |
| **Relations (N = 2)** | ( *n* = 0) | (*n* = 1) Belur 2017 | (*n* = 1) Nalugya 2020 |

**Supplemental Figure 1: Number of Articles Addressing Each Domain and Sub-Domain of Empowerment, by Water, Sanitation, or Water and Sanitation Focus**

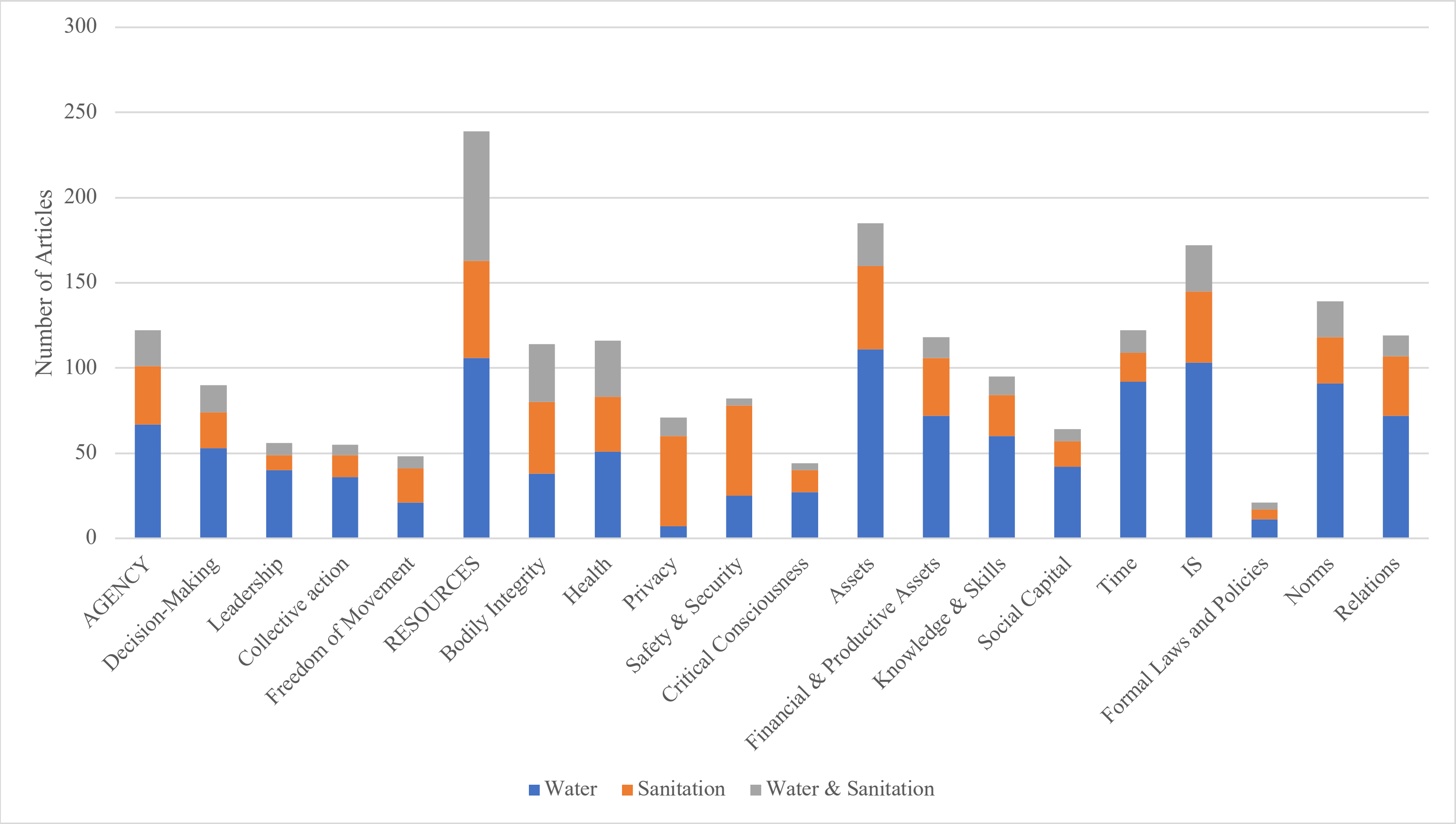

**Full Citations of Included Articles**

65. Coswosk ÉD, Neves-Silva P, Modena CM, Heller L. Having a toilet is not enough: the limitations in fulfilling the human rights to water and sanitation in a municipal school in Bahia, Brazil. *BMC Public Health* 2019; **19**(1): 137.
66. Coulter JE, Witinok-Huber RA, Bruyere BL, Dorothy Nyingi W. Giving women a voice on decision-making about water: barriers and opportunities in Laikipia, Kenya. *Gender, Place & Culture* 2018.
67. Crow B, Sultana F. Gender, class, and access to water: Three cases in a poor and crowded delta. *Society & Natural Resources* 2002; **15**(8): 709-24.
68. Crow B, Odaba E. Access to water in a Nairobi slum: Women's work and institutional learning. *Water International* 2010; **35**(6): 733-47.
69. Crow B, Swallow B, Asamba I. Community organized household water increases not only rural incomes, but also men’s work. *World Development* 2012; **40**(3): 528-41.
70. Czerniewska A, Muangi WC, Aunger R, Massa K, Curtis V. Theory-driven formative research to inform the design of a national sanitation campaign in Tanzania. *PloS one* 2019; **14**(8): e0221445.
71. Daniel D, Diener A, Pande S, et al. Understanding the effect of socio-economic characteristics and psychosocial factors on household water treatment practices in rural Nepal using Bayesian Belief Networks. *International Journal of Hygiene and Environmental Health* 2019; **222**(5): 847-55.
72. Das P. Women’s participation in community-level water governance in urban India: The gap between motivation and ability. *World Development* 2014; **64**: 206-18.
73. Das P, Baker KK, Dutta A, et al. Menstrual hygiene practices, WASH access and the risk of urogenital infection in women from Odisha, India. *PloS One* 2015; **10**(6): e0130777.
74. Datta A, Ahmed N. Intimate infrastructures: The rubrics of gendered safety and urban violence in Kerala, India. *Geoforum* 2020; **110**: 67-76.
75. de Moraes AFJ, Rocha C. Gendered waters: the participation of women in the ‘One Million Cisterns’ rainwater harvesting program in the Brazilian Semi-Arid region. *Journal of Cleaner Production* 2013; **60**: 163-9.
76. Delgado JV, Zwarteveen M. The public and private domain of the everyday politics of water: the constructions of gender and water power in the Andes of Perú. *International Feminist Journal of Politics* 2007; **9**(4): 503-11.
77. Devasia L. Safe drinking water and its acquisition: Rural women's participation in water management in Maharashtra, India. *International Journal of Water Resources Development* 1998; **14**(4): 537-46.
78. Devasia VV. Tribal women in sustainable development through watershed programmes in Vidarbha. *International Journal of Water Resources Development* 1998; **14**(4): 527-35.
79. DeVries K, Rizo A. Empowerment in action: Savings groups improving community water, sanitation, and hygiene services. *Enterprise Development & Microfinance* 2015; **26**(1): 34-44.
80. Dreibelbis R, Greene LE, Freeman MC, Saboori S, Chase RP, Rheingans R. Water, sanitation, and primary school attendance: A multi-level assessment of determinants of household-reported absence in Kenya. *International Journal of Educational Development* 2013; **33**(5): 457-65.

225. Torri MC. Power, structure, gender relations and community-based conservation: The Cawswe Study of the Sariska Region, Rajasthan, India. *Journal of International Women's Studies* 2010; **11**(4): 1-18.
226. Tortajada C. Professional women and water management: Case study from Morocco: A Water Forum contribution. *Water International* 2003; **28**(4): 532-9.
227. Trinies V, Freeman MC, Hennink M, Clasen T. The role of social networks on the uptake of household water filters by women in self-help groups in rural India. *Journal of Water, Sanitation and Hygiene for Development* 2011; **1**(4): 224-32.
228. Trinies V, Caruso BA, Sogoré A, Toubkiss J, Freeman MC. Uncovering the challenges to menstrual hygiene management in schools in Mali. *Waterlines* 2015; **34**(1): 31-40.
229. Tsai AC, Kakuhikire B, Mushavi R, et al. Population-based study of intra-household gender differences in water insecurity: Reliability and validity of a survey instrument for use in rural Uganda. *Journal of Water and Health* 2016; **14**(2): 280-92.
230. Van Houweling E, Hall R, Diop AS, Davis J, Seiss M. The role of productive water use in women's livelihoods. Evidence from rural Senegal. *Water Alternatives* 2012; **5**(3): 658.
231. Van Houweling E. Gendered water spaces: A study of the transition from wells to handpumps in Mozambique. *Gender, Place & Culture* 2015; **22**(10): 1391-407.
232. Van Houweling E. "A good wife brings her husband bath water": Gender roles and water practices in Nampula, Mozambique. *Society & Natural Resources* 2016; **29**(9): 1065-78.
233. Varickanickal J, Bisung E, Elliott SJ. Water risk perceptions across the life-course of women in Kenya. *Health Promotion International* 2019.
234. Varua ME, Ward J, Maheshwari B, Dave S, Kookana R. Groundwater management and gender inequalities: The case of two watersheds in rural India. *Groundwater for Sustainable Development* 2018; **6**: 93-100.
235. von Medeazza G, Jain M, Tiwari A, Shukla JP, Kumar N. Women-led total sanitation: Saving lives and dignity. In: Cronin AA, Mehta PK, Prakash A, eds. Gender Issues in Water and Sanitation Programmes: Lessons from India: SAGE Publications Pvt. Ltd.; 2015: 231-49.
236. Wall LL, Teklay K, Desta A, Belay S. Tending the 'monthly flower:'a qualitative study of menstrual beliefs in Tigray, Ethiopia. *BMC Women's Health* 2018; **18**(1): 183.
237. Waterkeyn J, Cairncross S. Creating demand for sanitation and hygiene through Community Health Clubs: A cost-effective intervention in two districts in Zimbabwe. *Social Science & Medicine* 2005; **61**(9): 1958-70.
238. Whale K, Cramer H, Joinson C. Left behind and left out: The impact of the school environment on young people with continence problems. *British Journal of Health Psychology* 2018; **23**(2): 253-77.
239. White S, Kuper H, Itimu-Phiri A, Holm R, Biran A. A qualitative study of barriers to accessing water, sanitation and hygiene for disabled people in Malawi. *PloS One* 2016; **11**(5): e0155043.
240. Willetts J, Halcrow G, Carrard N, Rowland C, Crawford J. Addressing two critical MDGs together: Gender in water, sanitation and hygiene initiative. *Pacific Economic Bulletin* 2010; **25**(1): 162-76.
